# Multiscale characterization of cortical signatures in positive and negative schizotypy: A worldwide ENIGMA study

**DOI:** 10.1101/2024.05.03.24306736

**Authors:** Matthias Kirschner, Benazir Hodzic-Santor, Leda Kennedy, Justine Y. Hansen, Mathilde Antoniades, Igor Nenadić, Tilo Kircher, Axel Krug, Tina Meller, Udo Dannlowski, Dominik Grotegerd, Kira Flinkenflügel, Susanne Meinert, Tiana Borgers, Janik Goltermann, Tim Hahn, Joscha Böhnlein, Elisabeth J. Leehr, Carlotta Barkhau, Alex Fornito, Aurina Arnatkeviciute, Mark A. Bellgrove, Jeggan Tiego, Pamela DeRosse, Melissa Green, Yann Quidé, Christos Pantelis, Raymond Chan, Yi Wang, Ulrich Ettinger, Martin Debbané, Melodie Derome, Christian Gaser, Bianca Besteher, Kelly Diederen, Tom J. Spencer, Josselin Houenou, Edith Pomarol-Clotet, Raymond Salvador, Wulf Rössler, Lukasz Smigielski, Veena Kumari, Preethi Premkumar, Haeme R. P. Park, Kristina Wiebels, Imke Lemmers-Jansen, James Gilleen, Paul Allen, Jan-Bernard Marsman, Irina Lebedeva, Alexander Tomyshev, Anne-Kathrin Fett, Iris Sommer, Sanne Koops, Phillip Grant, Indrit Bègue, Dennis Hernaus, Maria Jalbrzikowski, Casey Paquola, Sara Larivière, Boris Bernhardt, Sofie Valk, Bratislav Misic, Theo G. M. van Erp, Jessica A. Turner, Paul M. Thompson, Andre Aleman, Alain Dagher, Stefan Kaiser, Gemma Modinos

## Abstract

Positive and negative schizotypy reflect distinct patterns of subclinical traits in the general population associated with neurodevelopmental and schizophrenia-spectrum pathologies. Yet, a comprehensive characterization of the unique and shared neuroanatomical signatures of these schizotypy dimensions is lacking. Leveraging 3D brain MRI data from 2,730 unmedicated healthy individuals, we identified neuroanatomical profiles of positive and negative schizotypy and systematically compared them to disorder-specific, micro-architectural, connectome, and neurotransmitter-level measures. Positive and negative schizotypy were associated with thinner frontal and thicker paralimbic cortical areas, respectively, and were differentially linked to cortical patterns of schizophrenia-spectrum and neurodevelopmental conditions. Furthermore, these schizotypal cortical patterns mapped onto local attributes of gene expression, cortical myelination, D1 and histamine receptor distributions. Network models identified cortical hub vulnerability to schizotypy-related thickness reduction and epicenters in sensorimotor-to-association and paralimbic areas. This study yields insights into the complex cortical signatures of schizotypy and their relationship to diverse features of cortical organization.

## Introduction

Schizophrenia and related psychotic disorders exist on a spectrum, and the associated symptoms vary along a continuum from health to illness ^1–4^. Schizotypy describes a set of multidimensional traits in the general population associated with genetic, neurobiological, and behavioral liability to psychotic and related disorders ^1,5,6^. In this regard, two central dimensions of psychopathology are expressed in personality, schizotypy and schizophrenia symptoms^7,8^: One positive dimension (or psychoticism) –defined by mistrust, positive schizotypy, and positive symptoms– and one negative dimension (or detachment) spanning introversion (low extraversion), negative schizotypy, and negative symptoms. Both dimensions are linked to the schizophrenia spectrum and also intersect with neurodevelopmental disorders including autism spectrum disorder (ASD), attention-deficit/hyperactivity disorder (ADHD), and 22q11.2 deletion syndrome (22q11DS). Individuals with ASD, ADHD, and 22q11DS tend to show high schizotypal traits^9–13^, and ADHD in childhood/adolescence is predictive of higher schizotypal traits in adulthood^14^. In addition, more specific associations between negative schizotypy and ASD^11,13,15^ and positive schizotypy and 22q11DS^16,17^ have been reported. Collectively, these findings suggest common but also divergent associations of positive and negative schizotypy with schizophrenia spectrum disorders (SSD) as well as neurodevelopmental conditions. Thus, characterising the brain’s morphometric correlates of positive and negative schizotypy may inform both neurobiological pathways and protective mechanisms related to schizophrenia-spectrum and neurodevelopmental conditions, without confounding by antipsychotic medications or comorbidities with other mental health disorders.

While subtle structural and functional alterations have been associated with schizotypy and non-clinical psychotic symptoms in the general population^18–29^, prior research has been limited by methodological heterogeneity and small sample sizes in individual studies^30^. Recognizing these challenges, the ENIGMA Schizotypy Working Group recently coordinated an international analysis of morphometric profiles associated with overall schizotypy (n=3,004 individuals), using standardized image analyses, quality assurance and statistical analyses pipelines across cohorts^31^. We identified a distinct neuroanatomical profile characterized by thicker medial orbitofrontal/ventromedial prefrontal cortex (mOFC/vmPFC) associated with higher total schizotypy scores^31^. Furthermore, brain-wide cortical thickness patterns in schizotypy^31^ were significantly associated with patterns of cortical thinning observed in schizophrenia^32^, suggesting a neurobiological continuum within the extended psychosis phenotype. Parallel research has expanded this perspective, showing that genetic risk, clinical high-risk and neuropsychiatric conditions, including schizophrenia, exhibit spatial similarity of cortical alterations^33–35^. These shared patterns may be driven by a complex interplay of molecular and connectomic vulnerabilities^36^ as well as neurotransmitter profiles^37^, which manifest as coordinated alterations in cortical thickness^38,39^. In this context, two network mechanisms are proposed to guide such coordinated processes of morphometric alterations across risk stages and pathologies. First, it has been demonstrated that both structural and functional cortical hubs are more susceptible to alterations in neuropsychiatric conditions^36,38,40–42^. Second, it has been shown that disease-specific and cross-disorder alteration patterns are constrained by the normative connectivity profiles of distinct brain regions. This suggests that cortical alterations propagate in a network-like manner from one or more epicenters to the most closely connected brain regions^36,38,40,42–46^. However, the extent to which these mechanisms occur on a continuum and may reflect subtle neuroanatomical variations associated with schizotypal traits in the general population remains unknown.

Here we address these questions by conducting a multiscale analysis of the neuroanatomical variations related to positive and negative schizotypy. We first provide a large-scale meta-analysis of neuroanatomical signatures of positive and negative schizotypy, drawing on comprehensive data of 2,730 unmedicated healthy individuals from the ENIGMA-Schizotypy working group. We then systematically compare the cortical thickness profiles associated with positive and negative schizotypy to multiple existing resources of brain data spanning disease-related cortical abnormality maps, micro-architecture, and global connectomics^36,37^. Specifically, we contrast the cortical alteration patterns of positive and negative schizotypy with meta-analytic cortical alteration maps of schizophrenia spectrum conditions, including those found in groups of people at clinical high-risk for psychosis^47^ and with established schizophrenia^32^. To uncover shared and distinct associations with neurodevelopmental disorders, we also compare the schizotypy-related cortical patterns to cortical effect size maps of ASD^48^, ADHD^49^, and 22q11DS^50^. Further, we compare the cortical profiles of positive and negative schizotypy to detailed brain maps of multiple micro-architectural measures, including gene expression, metabolism, and myelination, alongside global connectomic measures such as number of connections, centrality, and diversity, and delineate the role of specific neurotransmitters. To further examine the association between schizotypal cortical maps and normative brain architecture, two network susceptibility models are applied: hub vulnerability, which compares regional network centrality with the magnitude of disease-specific atrophy; and epicenter mapping, which identifies regions whose typical connectivity profiles most closely reflect the spatial pattern of disease-specific alterations. Overall, we uncover how positive and negative schizotypy map onto multiple scales of biological and cortical organization, offering new insights into the neurobiological foundations of these complex schizophrenia-related traits in the general population.

## Results

We studied 2,730 unmedicated healthy individuals (12-68 years, 47% male) with varying levels of self-reported positive and negative schizotypy derived from 31 sites of the worldwide ENIGMA Schizotypy Working Group (Tables S1-4). By applying standardized protocols to 3D brain MRI scans, cortical thickness (CT) and surface area (SA) were measured of 68 gray matter brain regions, based on the Desikan-Killiany-Tourville (DKT) anatomical atlas^51^, and subcortical volume (SV) of 16 subcortical regions using FreeSurfer^52,53^ (http://surfer.nmr.mgh.harvard.edu) and standardized ENIGMA protocols (http://enigma.ini.usc.edu/protocols/imaging-protocols/). Following previous ENIGMA work^31,32^, we examined the associations between regional CT, SA, SV values and either positive or negative schizotypy scores using partial correlations including age and sex (and intracranial volume (ICV) for SV) as covariates for each site separately. We generated separate meta-analytic cortical effect size maps for positive and negative schizotypy including the Pearson’s *r* effect sizes from each site using inverse variance-weighted random effects models, which take into account different sample sizes across sites. We corrected for multiple comparisons using the false discovery rate (FDR; p_FDR_<0.05)^54^.

### Morphometric profiles of positive and negative schizotypy

Assessing the relationship between both schizotypy dimensions and regional CT, we found that higher positive schizotypy was significantly associated with thinner left inferior frontal gyrus *pars opercularis* (*r*=-0.07, *p*_FDR_=0.018, 95% CI [-0.109, -0.034]) and *pars orbitalis* (*r*=-.06, *p*_FDR_=0.043, 95% CI [-0.101, -0.026]), and at a trend-level with thinner right postcentral gyrus (*r*=-0.07, *p*_FDR_=0.08, 95% CI [-0.137, -0.012]) (Figures 1-2, Table S5). In contrast, higher negative schizotypy was significantly associated with thicker right mOFC/vmPFC (right, *r*=0.07, *p*_FDR_ =0.01, 95% CI [0.033, 0.112]; left, *r*=0.05, *p*_FDR_ =0.25, 95% CI [0.008, 0.090]) and bilateral rostral anterior cingulate cortex (right, *r*=.06, *p*_FDR_ =0.03, 95% CI [0.023, 0.099]; left, *r*=.06, *p*_FDR_ =0.03, 95% CI [0.022, 0.098]) (Figs.1-2, Table S6). Moderator analyses showed that schizotypy questionnaire (for details see Methods “Assessment of positive and negative schizotypy"), FreeSurfer version and scanner field strength did not significantly affect these associations (all *p*_FDR_>.05) (Tables S7-S12). Meta-analysis of SA and SV did not reveal any significant associations with positive or negative schizotypy and were not considered in further multiscale analyses (for details see Tables S13-S16). Together, these findings suggest opposing patterns of lower regional CT in individuals with higher positive schizotypy and higher regional CT in those with higher negative schizotypy (Figures 1-2).

**Fig. 1.**
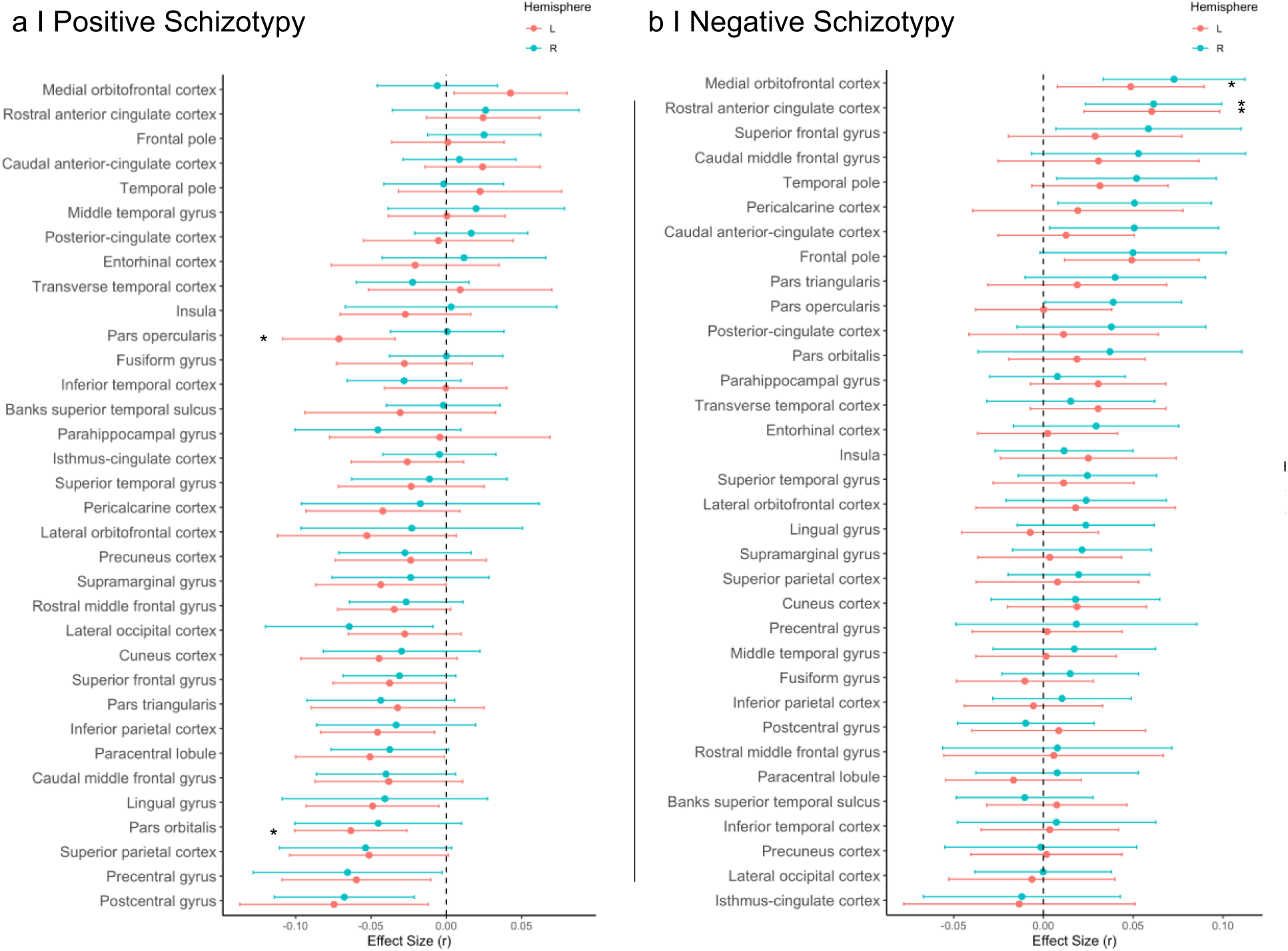
Effect sizes of partial correlation (*r*) between cortical thickness and schizotypy dimensions. Partial correlations (corrected for age and sex) between regional cortical thickness and **(a)** positive schizotypy, and **(b)** negative schizotypy. *regions with *p_FDR_* < 0.05.

**Fig. 2.**
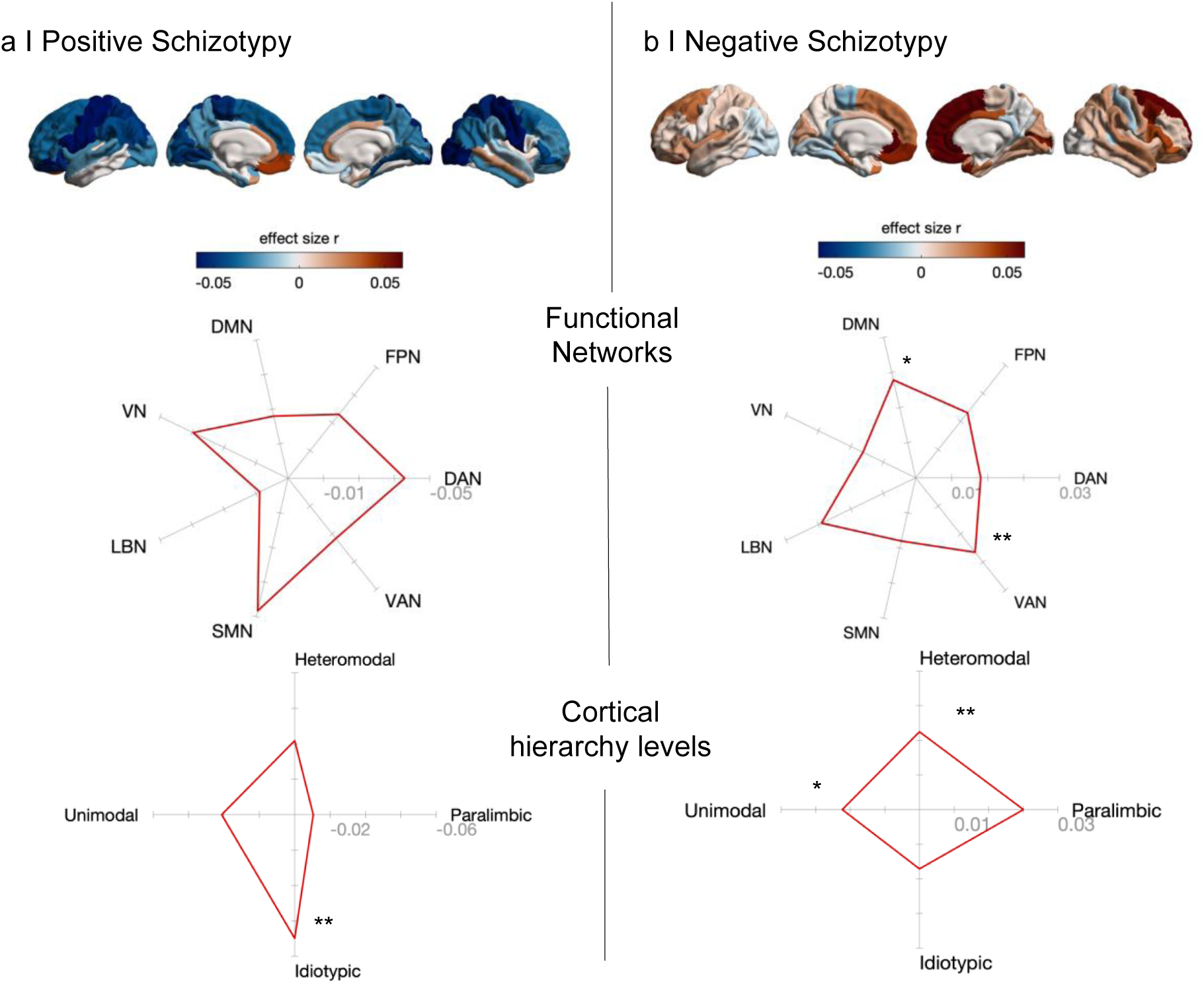
Cortical effects in positive and negative schizotypy aggregate within functional networks and cortical hierarchy levels. Cortical surfaces show the mean effect sizes (partial correlation r) for (a) positive and (b) negative schizotypy. Spider/Radar plots stratify the mean effect size values for positive and negative schizotypy according to functional networks^55^ and cortical hierarchy levels^56^. Significance was assessed with a one sample *t*-test against 1000 null models using spin permutation testing to account for spatial autocorrelation. In positive schizotypy, idiotypic-type regions exhibit significantly lower *t* statistics relative to spin permutation null models. In negative schizotypy, VAN, DMN functional networks, as well as heteromodal-, and unimodal-type regions show significantly higher t statistics relative to spin permutation models. DAN, Dorsal Attention Network; DMN, Default Mode Network; FPN, Fronto-Parietal Network; LBN, Limbic Network; SMN, Somato-Sensory Network; VAN, Ventral Attention Network; VN, Visual Network. *<0.05 p_spin_**<0.01 p_spin_

### Stratification of cortical thickness profiles of schizotypy to functional networks and cortical organization

To further contextualize the distinct schizotypy-dimension spatial patterning, we stratified the effect size maps of cortical thickness associations with positive and negative schizotypy according to intrinsic functional brain networks^55^ and levels of cortical hierarchy^56^. In positive schizotypy, thickness reduction was predominantly observed in the somatomotor (SMN)/visual (VN) and dorsal attention networks (DAN), but these effects were not significant in comparison to null models preserving spatial autocorrelation (all *p*_spin_>0.05). With respect to cortical hierarchy levels, we observed a significant aggregation of the association between positive schizotypy and reduced cortical thickness in idiotypic regions (mean=-0.05, *t*=-183.52, *p*_spin_=0.008). The association between negative schizotypy and greater cortical thickness was significantly stronger than null models in the visual attention network (VAN) (mean=0.019, *t*=52.28, *p*_spin_=0.005) and the default mode network (DMN) (mean=0.019, *t*=52.55, *p*_spin_=0.026). With respect to cortical hierarchy levels, the association between negative schizotypy and thickness increase was significantly more pronounced in heteromodal (mean=0.016, *t*=70.04, *p*_spin_=0.009) and unimodal regions (mean=0.014, *t*=56.97, *p*_spin_=0.038) (Fig. 2). Together, these observations suggest that the spatial patterns of associations between CT and higher positive and negative schizotypy are concentrated in different functional networks (SMN/VN/DAN vs. DMN/VAN) and levels of cortical organization (Idiotypic vs Unimodal/Heteromodal).

### Cortical pattern similarity of schizotypy dimensions with neurodevelopmental disorders

We tested whether the spatial patterns of cortical effect size maps of positive and negative schizotypy (partial correlation, *r*, Fig. 2) are related to case-control cortical effect size patterns (Cohen’s *d*) of schizophrenia-spectrum and neurodevelopmental disorders. To this end, we systematically correlated the cortical effect size maps of both schizotypy dimensions with the case-control effect size maps of each disorder derived from recently published ENIGMA studies^32,48–50^. Specifically, we included meta- and mega-analytic case-control cortical maps of individuals at clinical high-risk (CHR) for psychosis who subsequently developed psychosis (CHR-converters)^47^, schizophrenia^32^, 22q11DS individuals with psychosis (22q11DS-psychosis)^50^, ADHD^49^, and ASD^48^. Both positive and negative schizotypy patterns of cortical differences showed significant positive correlations with the cortical alteration pattern in schizophrenia (positive: *r*=0.221, *p*_spin_=0.041; negative: *r*=0.334, *p*_spin_=0.008). In contrast, only the negative schizotypy pattern was related to the cortical alteration pattern of CHR-converters (positive: *r*=-0.001, *p*_spin_>0.05; negative: *r*=0.24, *p*_spin_=0.03) (Fig. 3). Similar results were found for the cortical profiles of all CHR individuals (including those who did not develop psychosis) compared to healthy controls (Fig. S1).

**Figure 3.**
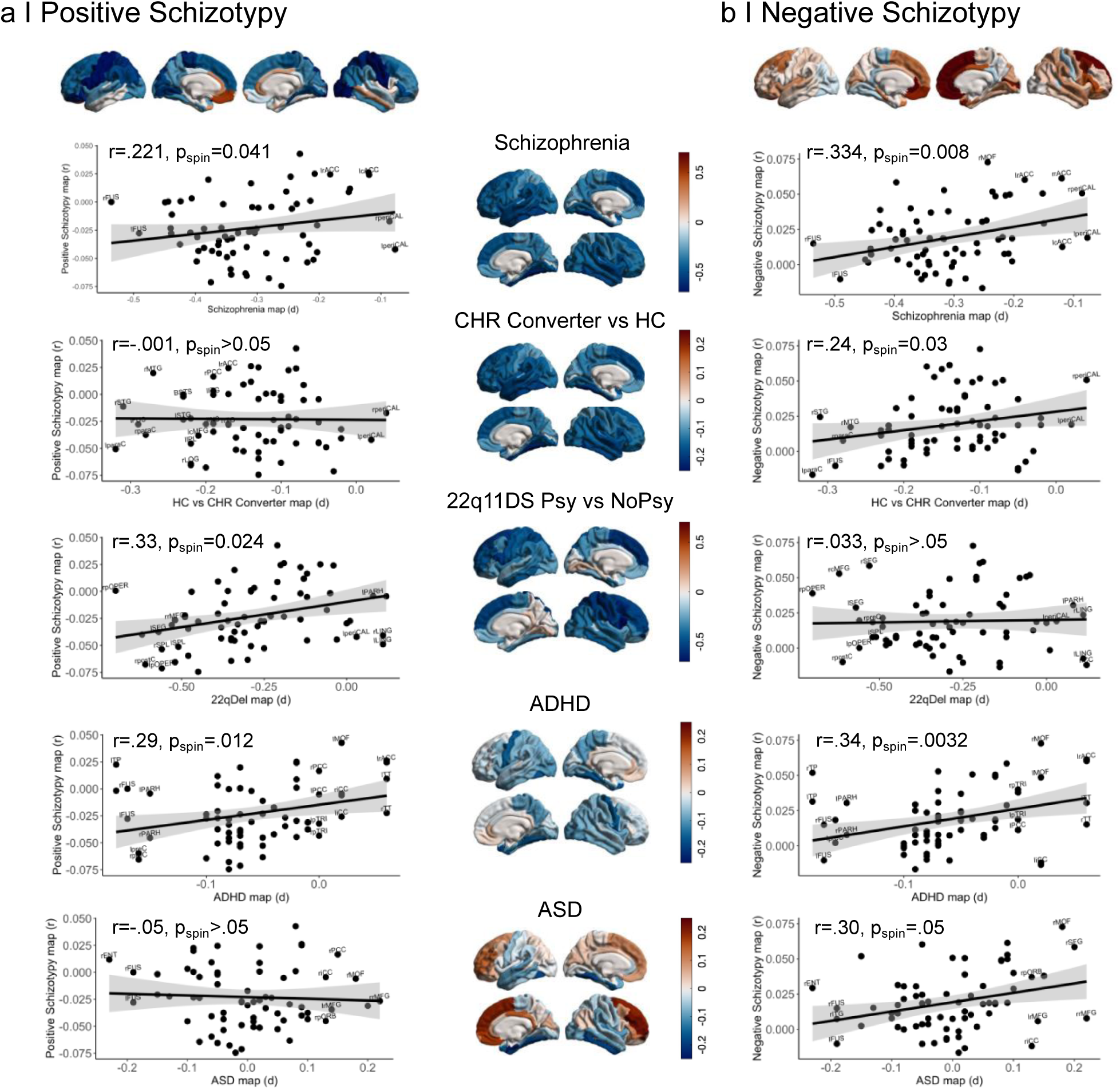
Cortical pattern similarity of schizotypy dimensions with schizophrenia and neurodevelopmental disorders. The cortical surfaces show the partial correlation effect sizes of positive and negative schizotypy, and the case-control effect sizes (Cohen’s D) of schizophrenia, CHR individuals who converted to psychosis, 22q11DS-psychosis vs non-psychosis individuals, children with ADHD (age 4-14 years), and ASD on cortical thickness from ENIGMA meta-/mega-analyses of each disorder^32,47–50^. Significance was assessed using spin permutation (1000 repetitions) to account for spatial autocorrelation between two brain maps^57^. Scatterplots show the correlation of each disorder map with **(a)** cortical effects of positive schizotypy and **(b)** cortical effects of negative schizotypy.

With respect to neurodevelopmental disorders, the cortical pattern of positive schizotypy showed significant positive correlations with the case-control cortical alteration patterns of 22q11DS-psychosis (*r*=0.33, *p*_spin_=0.024) and ADHD (*r*=0.29, *p*_spin_=0.012), but not ASD (*r*=-0.05, *p*_spin_>0.05) (Fig. 3). The spatial pattern of cortical differences in negative schizotypy showed significant positive correlations with the cortical alteration patterns in ADHD (*r*=0.34, *p*_spin_=0.003) and ASD (*r*=0.30, *p*_spin_=0.05), but not in 22q11DS-psychosis (*r*=0.033, *p*_spin_>0.05) (Fig. 3). These findings suggest that both schizotypy dimensions significantly overlap with the cortical alteration patterns seen in established schizophrenia, but only negative schizotypy is associated with the cortical profile seen in early psychosis. Furthermore, the results indicate overlapping associations with the cortical alteration pattern of ADHD, and non-overlapping associations with the cortical profiles in 22q11DS-psychosis and ASD.

### Molecular, connectivity and neurotransmitter contributions to cortical morphology of schizotypy dimensions

We next examined how cortical differences in positive and negative schizotypy map onto specific molecular gradients, structural network properties, and neurotransmitter receptors/transporter distributions. To address these questions, we used brain maps derived from recent publications by Hansen and colleagues^36,37^. These include molecular, structural connectivity and neurotransmitter profiles that are parcellated according to the same 68 regions of the DKT atlas, allowing for direct comparison with the cortical morphology pattern of positive and negative schizotypy (for details see^36,37^ and Materials and Methods). Following previous work^36,37^, we fitted multiple linear models using three sets of predictors – molecular, structural connectivity, and neurotransmitter features – against cortical morphology maps for positive and negative schizotypy separately, resulting in six distinct model fits (Fig. 4a). We found that cortical morphology of positive schizotypy was significantly predicted by each set of features (adj *R²*>.2, *p*<0.005) (Fig. 4a). The cortical morphology of negative schizotypy was significantly predicted by molecular and neurotransmitter features (both adj *R²*>.2, *p*<0.005), while the model fit including connectivity features did not reach significance (adj *R²*=.1, *p*=.06). These findings suggest that cortical morphology patterns of positive and negative schizotypy are both associated with molecular and neurotransmitter features.

**Fig 4.**
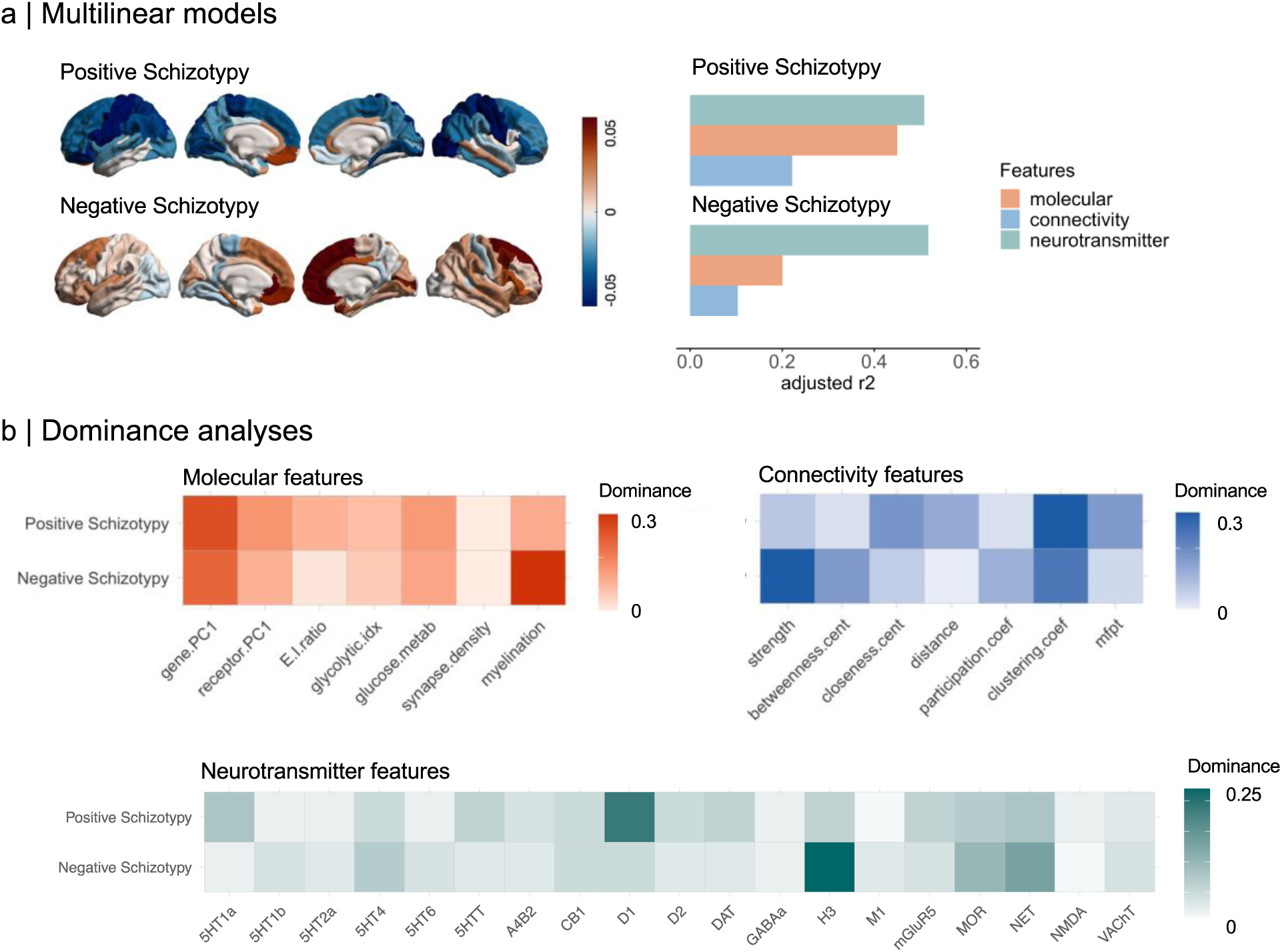
Molecular, connectivity, and neurotransmitter contributions to cortical morphology of schizotypy dimensions. **(a)** In total, six multilinear models were fit between molecular, structural connectivity, and neurotransmitter predictors to cortical morphology maps of positive and negative schizotypy. Adjusted *R2* is shown in the bar plot (*orange*: molecular; *blue*: connectivity; *green*: neurotransmitter). (**b)** Dominance analysis was applied to assess the contribution of each input variable to the fit of the model. This was done separately for the molecular (*orange*), connectivity (*blue*), and neurotransmitter (*green*) predictors. Molecular predictors: gene PC1 = first component of 11,560 genes’ expression; receptor PC1 = first component of 18 PET-derived receptor/transporter density; E:I ratio = excitatory:inhibitory receptor density ratio; glycolytic index = amount of aerobic glycolysis; glucose metabolism = [^18^F]-labelled fluorodeoxyglucose (FDG) PET image; synapse density = synaptic vesicle glycoprotein 2A (SV2A)-binding [^11^C]UCB-J PET tracer; myelination = T1w/T2w ratio. Connectivity predictors: strength = sum of weighted connections; betweenness = fraction of all shortest paths traversing region *i*; closeness = mean shortest path length between region *i* and all other regions; Euclidean distance = mean Euclidean distance between region *i* and all other regions; participation coefficient = diversity of connections from region *i* to the seven Yeo-Krienen resting-state networks^46^; clustering = fraction of triangles including region *i*; mean first passage time = average time for a random walker to travel from region i to any other region. Neurotransmitter predictors: Neurotransmitter predictors used in the multilinear models included 18 cortical receptor/transporter maps derived from PET tracer studies. These include dopamine (D1, D2, DAT), norepinephrine (NET), serotonin (5-HT1A, 5-HT1B, 5-HT2A, 5-HT4, 5-HT6, 5-HTT), acetylcholine (α4β2, M1, VAChT), glutamate (mGluR5), GABA (GABA_A_), histamine (H3), cannabinoid (CB1), and opioid (MOR). Data collection and processing of all PET maps into 68 cortical DKT regions is detailed in Hansen et al^37^.

To further examine the contribution (“dominance”) that each input variable has on the cortical morphology patterns of both schizotypy dimensions, we applied dominance analyses for each multilinear model separately (Fig 4b). Dominance analysis quantifies the relative importance or "dominance" of each input variable in contributing to the overall fit (adjusted *R²*) of the multiple linear regression model. In addition to the results above, dominance analyses revealed that the importance of predictors varied between schizotypy dimensions (Fig. 4b). For CT pattern related to positive schizotypy, the first principal gene expression gradient from Allen Human Brain Atlas (a potential proxy for cell-type distribution^58–61^), and dopamine D1 receptor density were the most dominant predictors. In contrast, for the CT pattern related to negative schizotypy, myelination and histamine H3 receptor density demonstrated the greatest dominance in the multilinear models (Fig. 4b). Other predictors such as synapse density or NMDA receptor density were consistently less dominant in predicting cortical morphology of both schizotypy dimensions. Collectively, these findings point to divergent biological mechanisms underlying cortical morphology profiles of positive and negative schizotypy, with distinct molecular and neurotransmitter predictors demonstrating variable influence across these dimensions.

### Hub vulnerability and epicenters of schizotypy dimensions

Subsequently, we tested whether the hub vulnerability hypothesis –that is, that regions with higher network centrality are more susceptible to pathological and maladaptive processes in neuropsychiatric disorders than non-hub regions^40–42,45,62^– extends to positive and negative schizotypy. To this end, we examined whether cortical hub regions (defined by higher normative network centrality derived from an independent healthy human connectome project (HCP) sample^63^, n=207) display stronger associations (partial correlation, *r*) with positive or negative schizotypy compared to non-hub regions. We compared the spatial patterns of normative functional and structural nodal degree centrality (Figs. S2-S3) and the cortical thickness patterns of positive and negative schizotypy (Fig. 2). In positive schizotypy, we found a significant correlation between functional and structural centrality and CT reduction in association with positive schizotypy (functional: *r*=-.54, *p*_spin_= .003; structural *r*=- .25, *p*_spin_= .036, Fig. 5a). Conversely, regions with relative CT increase in relation to positive schizotypy showed relative lower function and structural centrality. In negative schizotypy, the same association was observed for functional but not structural cortical hub regions (functional: *r*=-.48, *p*_spin_= .018; structural *r*=-.13, *p*_spin_= .197, Fig. 5a). In other words, cortical hub regions were more likely to display lower CT in relation to either higher positive or higher negative schizotypy scores, while regions with lower centrality in the network were more likely to exhibit greater CT in association to either positive or negative schizotypy.

**Fig 5.**
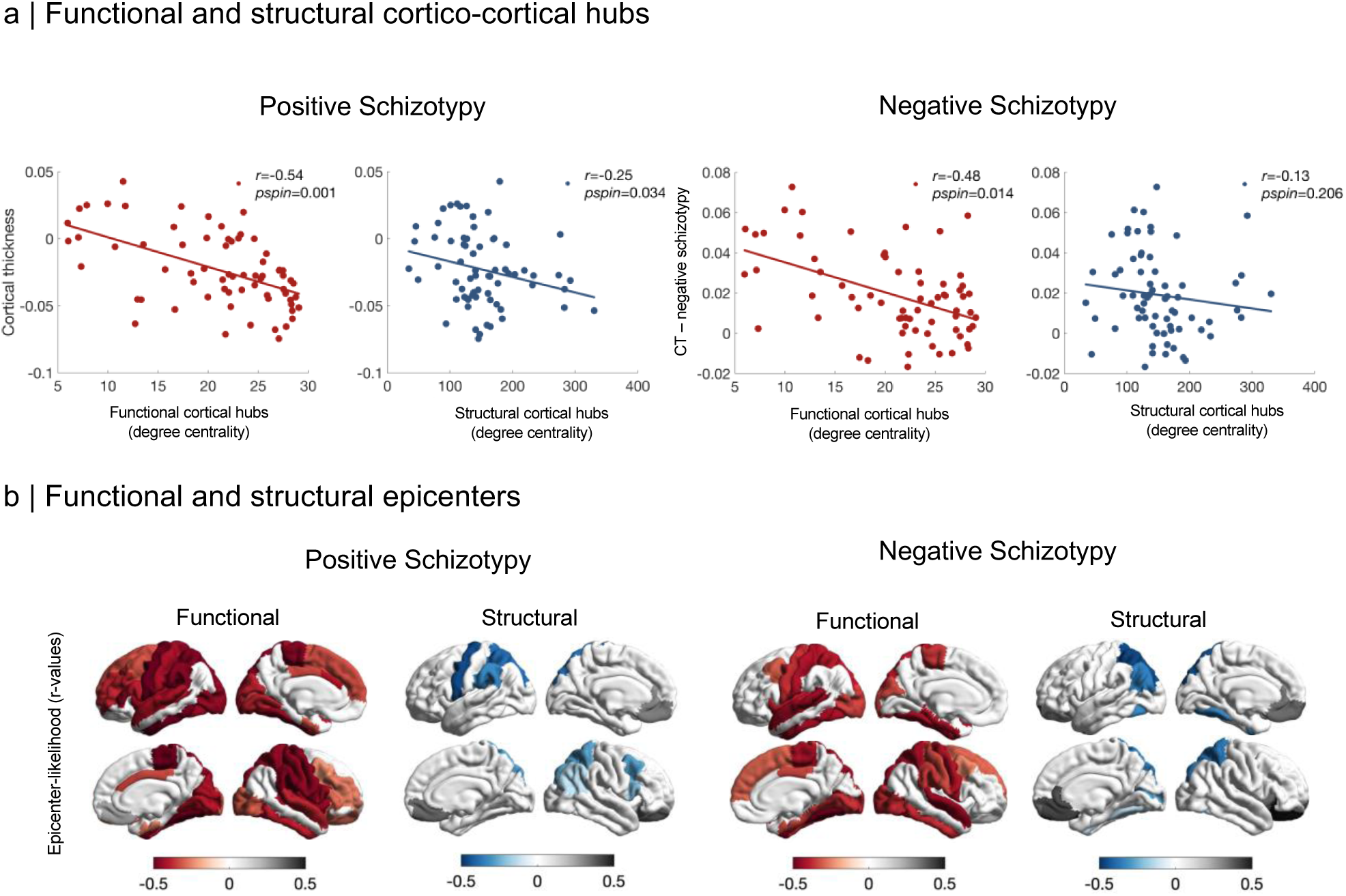
Hub and epicenter models of cortical morphology patterns in positive and negative schizotypy. **(a)** Correlation between cortical morphology patterns of positive and negative schizotypy with normative node-level functional (*red*) and structural (*blue*) degree centrality maps (derived from the healthy adult HCP dataset). In positive schizotypy, regions with high functional or structural centrality are significantly more likely to display reduced cortical thickness and regions with low functional or structural centrality are more likely to display cortical thickness increase. The same association was found for negative schizotypy and functional cortical hub regions but not structural cortical hubs. (**b)** Epicenter maps depicting the strength of associations (correlation coefficients) between the normative region-based functional (*red*) and structural (*blue*) connectivity and cortical morphology maps of positive and negative schizotypy. Significant epicenters with negative *r*-values are regions that are more strongly connected to regions with more reduced cortical thickness in association with positive and negative schizotypy (indicated by red and blue color range respectively). Inversely, significant epicenters with positive r-values are more strongly connected to regions with greater cortical thickness in association with positive and negative schizotypy (indicated by gray color range). Only significant epicenters with *p*_spin_<0.05, after spin permutation test (1000 repetitions) are displayed.

Finally, we aimed to identify putative epicenters of the cortical thickness patterns of positive and negative schizotypy. Epicenters are defined as regions whose normative connectivity profile (derived from the independent healthy adult HCP dataset^63^ mentioned above, Figs. S2-S3) most closely resembles disease-related morphological alteration patterns^43,45,64,65^. That is, epicenters are connected to brain regions with the strongest cortical thickness differences. Significant negative epicenter-likelihood (R-values) indicates that the connectivity profile of a region is more strongly associated with lower CT (meta-analytic negative partial R-values). Positive epicenter-likelihood reflects that the connectivity profile of a region is more strongly related to greater CT (meta-analytic positive partial R-values). For lower CT associated with positive or negative schizotypy, we observed several functional epicenters distributed across sensorimotor to association regions, and structural epicenters more circumscribed within sensorimotor and parietal cortices (Fig. 5b, Table S17-S18). In contrast, epicenters of greater CT in both positive and negative schizotypy were exclusively localized in the mOFC/vmPFC, lateral OFC, and ACC (Fig. 5b, Table S19-S20). In summary, epicenters of schizotypy-related CT reduction were more widely distributed across sensorimotor-associative regions, and epicenters of schizotypy-related CT increase were more circumscribed in the mOFC/vmPFC and adjacent regions.

## Discussion

Capitalizing on large-scale neuroimaging data from the ENIGMA Schizotypy Working Group, the key finding of our study is the identification of overlapping and distinct neuroanatomical signatures of positive and negative self-reported schizotypy in the general population. More specifically, higher positive schizotypy scores were associated with a cortical profile of predominantly CT reduction, with strongest associations in the inferior frontal gyrus. Higher negative schizotypy was associated with a pattern of mostly CT increase, showing strongest associations in the mOFC/vmPFC and rostral ACC. The spatial pattern of both CT profiles was constrained to different functional networks and regions with distinct levels of cortical hierarchy. Associations between CT and positive schizotypy were predominantly found in idiotypic regions, whereas significant associations with negative schizotypy were mainly localized in the VAN and DMN as well as in heteromodal and unimodal regions. We further observed that the cortical profiles of both schizotypy dimensions were spatially correlated with those in schizophrenia and ADHD. In addition, distinct associations emerged, with positive schizotypy-related cortical alterations correlating with those in 22q11DS-psychosis, and negative schizotypy-related cortical alterations with those in CHR-converters and ASD. These findings suggest overlapping but also divergent neurodevelopmental mechanisms related to positive and negative schizotypy. Linking the cortical profiles of both schizotypy dimensions to microarchitectural measures, distinct molecular and neurotransmitter features emerged as predictors of cortical variations in schizotypy, suggesting specific biological mechanisms involved. Finally, comparing the cortical profiles of both schizotypy dimensions with normative functional and structural connectivity, we found that cortical hubs were more vulnerable to the CT associations with positive and negative schizotypy. In addition, sensori-motor and prefrontal areas emerged as putative epicenters of CT reduction and increase, respectively. Collectively, this work advances our understanding of the neuroanatomy of positive and negative schizotypy in healthy individuals, without confounding by medication or comorbidities, reveals links to schizophrenia-spectrum and related neurodevelopmental conditions, and uncovers relationships with cortical organization, network characteristics, and biological mechanisms.

Our analyses revealed that higher positive schizotypy predominantly correlated with lower CT, while higher negative schizotypy predominantly correlated with higher CT. Interestingly, greater OFC/vmPFC thickness emerged as a signature common to both dimensions (Fig. 1), albeit only to a significant degree in negative schizotypy, mirroring our prior meta-analytic results using total schizotypy scores^31^. These findings suggest that neuroanatomical variations observed in positive and negative schizotypy may reflect distinct neurodevelopmental mechanisms of accelerated versus delayed cortical maturation. Consistent with this notion, the positive and negative schizotypy-related cortical profiles spatially overlapped with those of several psychiatric and neurodevelopmental conditions, including schizophrenia-spectrum conditions, ADHD, 22q11DS, and ASD. We found that the cortical alteration patterns of both schizotypy dimensions were spatially related to those seen in schizophrenia, confirming previous findings of a neuroanatomical continuum across the extended psychosis phenotype^31,47,50^. Furthermore, the cortical profile of negative schizotypy, but not positive schizotypy, was spatially associated with cortical alterations seen in CHR individuals who subsequently transitioned to psychosis^38^. Psychometric studies have repeatedly shown that both positive and negative schizotypy predict schizophrenia-spectrum psychopathology and disorders^1,5,66^. Our findings expand this body of work by suggesting that while behaviourally both schizotypy dimensions have been associated with transition to psychosis in CHR individuals^1,5,66^, neuroanatomically the negative schizotypy dimension profile associates more strongly with the CHR-converter profile. It should be noted that despite the observed spatial associations, the distributed greater CT in negative schizotypy contrasts with the predominantly reduced CT patterns of schizophrenia and CHR converters. This increased CT in negative schizotypy may reflect pre-existing processes prior to accelerated cortical thinning around the onset of psychosis. Alternatively, the observed divergent pattern -greater CT in negative schizotypy and lower CT in SSD-may suggest the absence of pathological or the presence of compensatory mechanisms in healthy individuals with negative schizotypy. In this context, advances in translating meta-analytic findings into individual vulnerability scores^67–69^ could be used to longitudinally assess whether higher similarity to the cortical profile of negative schizotypy improves the prediction of psychosis in CHR individuals.

The coordinated processes of cortical maturation during childhood and adolescence not only reflect typical development but also highlight potential vulnerabilities to neurodevelopmental conditions^35,70,71^. Comparing the CT profiles of neurodevelopmental conditions with those of both schizotypy dimensions, we observed overlapping associations of both schizotypy dimensions and ADHD. In addition, unique associations emerged between positive schizotypy and 22q11DS-psychosis, and between negative schizotypy and ASD. These findings are consistent with psychometric studies showing common but also distinct relationships of positive and negative schizotypy with subclinical and clinical symptoms of neurodevelopmental conditions^9,10,14–17^. An ADHD diagnosis during adolescence was found to predict higher total and negative schizotypy later in life^14^, and associations between higher ADHD features and positive schizotypy have been observed in relatives of individuals with schizophrenia^72,73^ and in healthy individuals^74,75^. Studies of 22q11DS individuals consistently reported elevated positive schizotypy scores^9,16,76^, mirroring the well-established increased risk of developing schizophrenia in this population^77^. In addition, 22q11DS individuals who later developed psychosis exhibited neuroanatomical alterations similar to those seen in schizophrenia^50^. For ASD, diametric associations of higher negative schizotypy with higher autistic traits and an inverse relationship between positive schizotypy and autistic traits were previously observed^11^. This supports our findings of a specific positive correlation between the CT profiles of negative schizotypy and ASD. Collectively, these findings suggest that schizotypal traits are related to complex variations of cortical development that may manifest in different psychotic and neurodevelopmental psychopathologies.

In this regard, a growing literature on cross-disorder effects revealed shared vulnerabilities that affect multiple scales of cortical organization including transcriptomics, neurotransmitter systems, and microstructural characteristics^34,36,38,78^. Local molecular features of neuroreceptor density, gene expression, and microstructure have been found to represent important predictors of disease-specific and cross-disorder cortical alteration patterns^34,36^. Our findings reveal that the cortical profiles of positive and negative schizotypy are not randomly distributed but can be explained by regional variations of molecular attributes and specific neurotransmitters. The cortical profile of positive schizotypy was significantly explained by a regional distribution of gene expression while the cortical morphology associated with negative schizotypy was significantly predicted by differences in T1w/T2w ratios, an MRI proxy for intracortical myelin content. Both gradients of gene expression and cortical myelination follow a sensorimotor to transmodal axis of cortical organization^58,61,70,79,80^ that has been linked to common neuroanatomical effects across neurodevelopmental and psychiatric disorders^34,36,70^. Here, we found that cortical profiles of positive and negative schizotypy relate to similar principles of cortical hierarchies suggesting common vulnerability during cortical development. With respect to neurotransmitter systems, prior work found that brain-wide receptor distributions were associated with cortical maps of major psychiatric disorders but not with the cortical profile of total schizotypy^37^. In the present study, however, decomposing total schizotypy into positive and negative schizotypy revealed significant associations with two different neurotransmitter systems. The cortical differences seen in positive schizotypy were explained by the distribution of cortical D1 receptor density, mirroring the above-mentioned dorsolateral to ventromedial cortical hierarchy^37^. In contrast, the negative schizotypy-related cortical differences were predicted by cortical H3 receptor distribution, which can be attributed to its high density in frontal regions, including the ACC and vmPFC. Cortical histamine modulates multiple cognitive processes^81,82^ and altered H3 receptor function is increasingly recognized as a potential mechanism underlying cognitive impairments in schizophrenia^83,84^. Therefore, although speculative, the associations of negative schizotypy-related cortical difference and H3 receptor distribution may reflect either compensatory mechanisms that preserve cognitive function or pre-existing factors underlying cognitive deficits of schizophrenia. Future work is needed to further decipher the putative pathological or compensatory nature of these newly identified spatial associations. Collectively, our findings indicate that the cortical patterns of both positive and negative schizotypy follow unifying principles of cortical transcriptomic and microstructural differentiation with additional contributions of distinct neurotransmitter systems. This might embed shared vulnerability to neurodevelopmental and neuropsychiatric conditions, but may also suggest resilience mechanisms. The differential contribution of gene expression and myelination may be further explained by subtle local variations in the cortical profiles of positive and negative schizotypy.

Finally, we contextualized the contribution of normative brain network architecture to the spatial pattern of cortical differences in positive and negative schizotypy. We provide evidence that regions with higher network centrality are more susceptible to cortical differences associated with both schizotypy dimensions. Within the topology of brain networks, centrally located neuronal populations are especially vulnerable to pathophysiological perturbations due to their higher neuronal activity and concomitant metabolic demands^41,42,85^. In addition, the most central hub regions are concentrated in heteromodal association cortices that, due to its prolonged window of developmental plasticity, have been shown to be especially vulnerable to insults during cortical development^70^. Such increased vulnerability of cortical hub regions to pathological processes have been consistently found in neurodegenerative disorders^86–89^, and more recently in neurodevelopmental and psychiatric conditions^40,41,90^. Several biological mechanisms could lead to a marked susceptibility of hub regions. Extending this literature, our findings demonstrate that hub vulnerability may already manifest itself in cortical variations associated with vulnerability traits such as schizotypy.

Spatial patterns of disease-specific and cross-disorder alterations can be explained, to some extent, by connectivity profiles of specific regions - or epicenters - that most closely resemble the corresponding morphometric alteration profile^36,38,40^. Pathological processes in brain disorders may spread in a network-like manner along the underlying normative connectome architecture^42,46^. For both positive and negative schizotypy, we observed epicenters of CT reduction in sensorimotor-to-association areas and epicenters of CT increase in the vmPFC/mOFC and ACC. The distributed sensorimotor-to association epicenters of CT reduction align well with recently reported epicenters of cross-disorder cortical abnormalities and co-alteration of neurodevelopmental, neurological and psychiatric disorders^36,38^. In a complementary way, the circumscribed paralimbic epicenters of CT increase closely mirror recent findings across different stages of schizophrenia^40,43,44^. Together, these associations with the connectome architecture may reflect how schizotypy-related cortical processes could contribute to both neurodevelopment in general and specific neuroanatomical processes related to psychosis.

The present work has some methodological limitations. First, significant associations of positive and negative schizotypy were only observed with regional CT and so even larger sample sizes might be needed to identify associations with subcortical volumes. Second, the molecular, and neurotransmitter maps are coming from different individuals/datasets, are all derived from healthy individuals, and are available only for cortical regions. Third, the present work considered neuroanatomical signatures of the positive and negative schizotypy based on the large body of research for these dimensions and the available data across sites. However, future efforts of the ENIGMA Schizotypy Working Group will aim to pool the necessary data for the disorganized schizotypy dimension^91^. Fourth, the current findings build on comparisons of meta-analytic cortical profiles of schizotypy dimensions with cortical effect size maps from disorder and at-risk populations, as well as group-level brain maps of molecular, connectivity, and neurotransmitter features derived from separate samples of healthy participants. As such, these findings do not capture individual variability of multiscale differentiation or allow for direct comparisons of different populations across the life-span. However, it should be noted that large-scale multiscale investigations using multiple PET scans and histology at the level of the individual are not possible and future work will continue to rely on data aggregation from different resources and cohorts. These are however powerful hypothesis-generating approaches that offer candidate targets for direct experimentation in future studies. Future efforts of the world-wide neuroscience community including the ENIGMA consortium^92–95^, will be needed to aggregate the necessary large-scale individual multimodal neuroimaging, genetic, and behavioral data across multiple neurodevelopmental and psychiatric disorders to address these questions at the level of the individual.

In summary, we identified cortical signatures of positive and negative schizotypy that are embedded along multiple scales of cortical organization and neuropsychiatric pathologies. More broadly, our work yields novel insights into how neurobiology and brain architecture may guide neuroanatomical vulnerability to general psychopathology and psychosis in the general population.

## Methods

### Study sample

This study included 2,730 unmedicated healthy individuals with varying levels of positive and negative schizotypy (below) from the worldwide ENIGMA Schizotypy Working Group. Sample-size average of mean (range) age across samples for this meta-analysis was 30.1 (12–68) years and samples were on average 46.5% male (27%-100%) (Table S1-S2). All participants were unmedicated healthy individuals with no history of psychiatric or neurological disorders. Study samples were collected with participants’ informed consent and approved by local ethics review boards.

### Assessment of positive and negative schizotypy

Across all 31 cohorts, schizotypy was assessed with well-validated self-report instruments, including the Chapman scales^96,97^, the Community Assessment of Psychotic Experiences (CAPE)^98^, the Schizotypal Personality Questionnaire (SPQ)^99^, the Oxford-Liverpool Inventory of Feelings and Experiences (O-LIFE)^100^, and the Rust Inventory of Schizotypal Cognitions (RISC)^101^. Positive schizotypy was assessed with the SPQ (or SPQ-B) cognitive/perceptual subscore, the CAPE positive dimension, the Chapman perceptual aberration score, the O-LIFE unusual experience score, and the RISC total score. Total negative schizotypy was assessed with the SPQ (SPQ-B) interpersonal subscore, the CAPE negative dimension, the Chapman physical and social anhedonia, and the O-LIFE introvertive anhedonia subscale (for details of schizotypy measures, see Table S4). In the case of two scores per site (London1a and Melbourne), the mean score of both scales was used.

### Image acquisition and processing

Following published ENIGMA procedures^31,32,102^, all sites processed T1-weighted structural scans using FreeSurfer^52,53^ (http://surfer.nmr.mgh.harvard.edu) and extracted CT, SA for 68 Desikan-Killiany (DKT) atlas regions^51^ (34 regions per hemisphere). In addition, subcortical volumes of 16 brain structures including left and right lateral ventricle, thalamus, caudate, putamen, pallidum, accumbens, hippocampus and amygdala, and intracranial volume (ICV) were extracted. Number of scanners, vendor, strength, sequence, acquisition parameters, and FreeSurfer versions are provided in Table S3. QC followed standard ENIGMA protocols at each site before analysis. For subcortical data, all regions of interest (ROIs) with a volume deviating from the mean by more than 1.5 times from the interquartile range were identified and only included after additional visual inspection. For cortical data, ENIGMA’s quality assurance protocol was performed (http://enigma.usc.edu/protocols/imaging-protocols) including visual inspection of the cortical segmentation and region-by-region removal of values from incorrect segmentations.

### Statistical meta-analyses

#### Model fit cortical measures

To examine the relationship between positive or negative schizotypy scores and CT (or SA) we fitted continuous models in each sample separately. We used partial correlation analysis (pcor.test, R version 3.6.0, R Foundation for Statistical Computing, Vienna, Austria)^103^ to assess the association between the CT (or SA) of left and right DKT atlas regions with positive or negative schizotypy scores including age, sex as covariates. For multiscanner studies (n=3), binary dummy covariates (n-1 scanners) were included within each local site model prior to meta-analysis to account for potential differences that may emerge across different scanners, consistent with the standard meta-analysis approach of previous ENIGMA studies^32,102^.

#### Model fit subcortical volumes

Similar to the cortical analyses, we applied continuous models to examine the relationship between schizotypy scores and subcortical volume for each ROI in each sample. To this end, partial correlation analysis was used to test correlations between the left and right subcortical volumes with positive and negative schizotypy scores including age, sex, and ICV as covariates. For multi-scanner studies (n = 3), binary dummy covariates were included to account for differences that may emerge across scanners, as described above.

#### Meta-analyses

Pearson’s r effect sizes from the partial correlations using schizotypy scores as a continuous predictor were meta-analyzed in separate random effects models to account for between study differences (rma function, metafor package for R 3.6.0)^104^. The false discovery rate (FDR) procedure (*p*_FDR_ < 0.05) was used to control for multiple comparisons^54^. Meta-analyses were adjusted for sample sizes across different sites and results were weighted for sample sizes. Possible confounding effects of schizotypy questionnaire type, FreeSurfer version, number of scanners, and scanner field strength were examined using moderator analyses (Tables S7-S12).

### Aggregation of the cortical profiles of positive and negative schizotypy within functional networks and cortical hierarchy levels

We stratified the regional effect sizes of cortical thickness associations with positive and negative schizotypy according to intrinsic functional brain networks^55^ and levels of cortical hierarchy^56^. Following previous work each DKT region was annotated to one of the seven functional network and four cortical hierarchy levels^34^. Mean cortical effect sizes were calculated separately for each network and for each level of the cortical hierarchy, based on the annotated individual DKT regions. We then calculated the mean t-values of the association between cortical thickness and positive or negative schizotypy for each functional network or cortical hierarchy. Significance of mean t-values in each functional network or cortical hierarchy was assessed with one sample t-test against 1000 null models using spin permutation testing to account for spatial autocorrelation^57,105^.

### Cortical pattern similarity of schizotypy dimensions with neurodevelopmental disorders

We examined how the spatial pattern of cortical differences in positive and negative schizotypy relate to the cortical alteration patterns observed in, CHR, schizophrenia, 22q11DS psychosis vs non-psychosis individuals, ADHD, and ASD. Similar to prior work^31,33,35,36,38,78^, we spatially correlated the cortical alteration patterns of positive and negative schizotypy (correlation coefficients) with the recently published ENIGMA meta-/mega-analyses of cortical abnormalities maps (Cohen’s *d*) of CHR^47^, schizophrenia^32^, 22q11S psychosis^50^, paediatric ADHD^49^, ASD^50^. Specifically, we systematically compared the spatial pattern similarity of cortical effect size maps for positive and negative schizotypy with each of the other disorder-specific cortical effect size maps, using Pearson correlations. Statistical significance of all cortical pattern correlations was assessed using spin permutation tests (1000 repetitions) correcting for spatial autocorrelation^57,106^ implemented in the ENIGMA toolbox^105^. This spatial autocorrelation approach–and the adequate control of false positives–has been comprehensively validated for parcellated ROI data, including resolutions of 68 parcellations as in our study^106^.

### Molecular, connectivity and neurotransmitter contributions to cortical morphology of schizotypy dimensions

We examined how cortical profiles in positive and negative schizotypy are informed by three different sets of molecular (or microarchitectural), normative structural connectivity, and specific neurotransmitter measures. All molecular, structural connectivity, and neurotransmitter maps provide regional values for each of the 68 DKT regions and were derived from two recent publication^36,37^. Details on the aggregation and processing of each brain map can be found in Hansen and colleagues for molecular and connectivity data^36^, and Hansen and colleagues for neurotransmitter maps^37^. Data can be obtained here: https://github.com/netneurolab/hansen_crossdisorder_vulnerability.git, https://github.com/netneurolab/hansen_receptors.git.

#### Molecular predictors

Following Hansen and colleagues^36^ we used a total of seven local molecular predictors in the multilinear models to examine the influence that local molecular attributes have on cortical morphology of positive and negative schizotypy.

#### Gene expression gradient

The first principal component of gene expression (“gene gradient”) was used to represent the variation in gene expression levels across the left cortex. Gene expression data of in total 11,560 genes derived from the Allen Human Brain Atlas as described in Hawrylycz^60^ and processed by *abagen*, an open-source Python toolbox^107^.

#### Receptor gradient

The first principal component of receptor density (“receptor gradient”) was used to represent the variation in receptor densities across the cortex. Receptor densities were estimated using positron emission tomography (PET) tracer studies in healthy volunteers, for a total of 18 receptors and transporters, across 9 neurotransmitter systems. For details on the included receptors and transporters see^37^.

#### Glycolytic index

This brain map provides glycolytic index values defined as the residual after fitting glucose metabolism to oxygen metabolism in a linear regression model. Larger values indicate more aerobic glycolysis. Original data were collected, calculated, and made available by Vaishnavi et al^108^.

#### Glucose metabolism

The glucose metabolism cortical map is based on [^18^F] -labelled fluorodeoxyglucose (FDG) PET scans in 33 healthy adults (19 female, mean age 25.4 ± 2.6 years) as described in detail in Vaishnavi et al.^108^.

#### Synapse density

The synapse density cortical maps is based on PET scans in 76 healthy adults (45 males, 48.9 ± 18.4 years of age) using [^11^C]UCB-J, a PET tracer that binds to the synaptic vesicle glycoprotein 2A (SV2A).

#### Myelination

The cortical myelination map is based on T1w/T2w ratios from healthy individuals (n= 417, age range 22–37 years, 193 males) of the Human Connectome Project (HCP, S1200 release)^109,110^.

#### Connectivity predictors

Following^36^, a total of nine global connectome predictors computed on weighted structural connectome were used in the multilinear model to examine their influence on cortical morphology of positive and negative schizotypy. Structural group-level connectome data are based on a healthy cohort (16 females, 25.3 ± 4.9) collected at the Department of Radiology, University Hospital Center and University of Lausanne. Structural connectivity estimation and group-consensus structural network generation are described in detail in Hansen et al.^36^. All connectivity measures were computed using the Python-equivalent of the Brain Connectivity Toolbox, bctpy. Connectivity predictors included: strength = sum of weighted connections; betweenness = fraction of all shortest paths traversing region i; closeness = mean shortest path length between region i and all other regions; Euclidean distance = mean Euclidean distance between region i and all other regions; participation coefficient = diversity of connections from region i to the seven Yeo-Krienen resting-state networks^55^; clustering = fraction of triangles including region i; mean first passage time = average time for a random walker to travel from region i to any other region.

#### Neurotransmitter predictors

Neurotransmitter predictors used in the multilinear models included 18 cortical receptor/transporter maps derived from PET tracer studies^37^. These include dopamine (D1, D2, DAT), norepinephrine (NET), serotonin (5-HT1A, 5-HT1B, 5-HT2A, 5-HT4, 5-HT6, 5-HTT), acetylcholine (α4β2, M1, VAChT), glutamate (mGluR5), GABA (GABA_A_), histamine (H3), cannabinoid (CB1), and opioid (MOR). Data collection and processing of all PET maps into 68 cortical DKT regions is described in detail in Hansen et al^37^.

### Dominance analysis

Dominance analysis is a technique designed to assess the individual significance of each predictor within the multiple linear regression framework in terms of their contribution to the model’s overall goodness of fit, as indicated by the adjusted R² (see https://github.com/dominance-analysis/dominance-analysis and^111,112^ for detailed methodology). In this approach the same regression model is fitted on every combination of input variables (2p − 1 submodels for a model with p input variables). Total dominance is quantified as the mean of the relative increase in R² observed when a particular predictor is incorporated into a submodel, averaged over all possible submodel combinations, which amounts to 2^p - 1 variations for p predictors. The sum of the dominance of all input variables is equal to the total adjusted R2 of the complete model, making total dominance an intuitive measure of contribution.

### Hub vulnerability model

Following previous work^38,40,64^, we tested the hub vulnerability hypothesis, *i.e.,* that nodes with higher normative network centrality derived from an independent sample of healthy individuals of the HCP would display higher levels of cortical differences related to positive or negative schizotypy. Using the analyses pipeline of the ENIGMA toolbox^105^, we assessed spatial correlations between the cortical morphology profiles of positive and negative schizotypy and normative weighted degree centrality maps (Fig. S1-S2). Weighted degree centrality was defined by the sum of all weighted cortico-cortical connections for every region and used to identify structural and functional hub regions. Regions with a higher weighted degree centrality are denoted as hubs, compared to regions with relatively lower weighted degree centrality. To mitigate potential bias from selecting an arbitrary threshold and inadvertently excluding valuable information, the analyses were conducted on unthresholded connectivity matrices. We corrected for spatial autocorrelation using the spin permutation procedure (1000 repetitions) implemented in the ENIGMA toolbox^105^ (see above).

### Mapping disease epicenters

Similar to the hub vulnerability model, we followed previous work^36,38,40,43,44,64^ and aimed to identify epicenters of cortical morphology patterns related to positive or negative schizotypy. Epicenters are defined as regions whose normative connectivity profile most closely resembles disease-related morphological alteration patterns^36,38,40,44,45,64^. In other words, disease epicenters are connected to those brain regions with the strongest disease-related morphological alterations. We spatially correlated every region’s healthy functional and structural cortical connectivity profile (Fig. S1-S2) to the whole-brain patterns of cortical differences in positive and negative schizotypy (Fig. 2). We repeated this approach systematically for each parcellated region and for functional and structural cortical connectivity separately. Regions were ranked in descending order based on the strength of their correlation coefficients, with the highest-ranked regions being considered the most significant disease epicenters. A region can be considered an epicenter regardless of its cortical difference magnitude (correlation coefficient of the partial correlations with either positive or negative schizotypy), as long as it is strongly connected to other regions with high cortical differences (higher *r*-values) and weakly connected to regions with low cortical alteration (*r*-values). Statistical significance of spatial similarity between an individual brain region’s functional and structural connectivity profile and schizophrenia-related cortical alterations was determined using the spin permutation procedure (1000 repetitions) implemented in the ENIGMA toolbox^105^ (see above).

## Data Availability

All data produced in the present study are available upon reasonable request to the authors

## References

1. Debbané, M. et al. Developing psychosis and its risk states through the lens of schizotypy. Schizophr Bull 41 Suppl 2, S396–407 (2015).

2. van Os, J. & Reininghaus, U. Psychosis as a transdiagnostic and extended phenotype in the general population. World Psychiatry 15, 118–124 (2016).

3. van Os, J., Linscott, R. J., Myin-Germeys, I., Delespaul, P. & Krabbendam, L. A systematic review and meta-analysis of the psychosis continuum: evidence for a psychosis proneness-persistence-impairment model of psychotic disorder. Psychol Med 39, 179–95 (2009).

4. Jonas, K. G. et al. Psychosis superspectrum I: Nosology, etiology, and lifespan development. Mol Psychiatry 1–15 (2024) doi:10.1038/s41380-023-02388-2.

5. Kwapil, T. R. & Barrantes-Vidal, N. Schizotypy: Looking Back and Moving Forward. Schizophr Bull 41, S366–S373 (2015).

6. Kwapil, T. R., Gross, G. M., Silvia, P. J. & Barrantes-Vidal, N. Prediction of psychopathology and functional impairment by positive and negative schizotypy in the Chapmans’ ten-year longitudinal study. J Abnorm Psychol 122, 807–815 (2013).

7. Kotov, R. et al. Validity and utility of Hierarchical Taxonomy of Psychopathology (HiTOP): I. Psychosis superspectrum. World Psychiatry 19, 151–172 (2020).

8. Cicero, D. C., Jonas, K. G., Li, K., Perlman, G. & Kotov, R. Common Taxonomy of Traits and Symptoms: Linking Schizophrenia Symptoms, Schizotypy, and Normal Personality. Schizophr Bull 45, 1336–1348 (2019).

9. Fonseca-Pedrero, E., Debbané, M., Schneider, M., Badoud, D. & Eliez, S. Schizotypal traits in adolescents with 22q11.2 deletion syndrome: validity, reliability and risk for psychosis. Psychological Medicine 46, 1005–1013 (2016).

10. Barneveld, P. S. et al. Overlap of autistic and schizotypal traits in adolescents with Autism Spectrum Disorders. Schizophr. Res. 126, 231–236 (2011).

11. Zhou, H.-Y. et al. Revisiting the overlap between autistic and schizotypal traits in the non-clinical population using meta-analysis and network analysis. Schizophr. Res. 212, 6–14 (2019).

12. Gong, J.-B., Wang, Y., Lui, S. S. Y., Cheung, E. F. C. & Chan, R. C. K. Childhood trauma is not a confounder of the overlap between autistic and schizotypal traits: A study in a non-clinical adult sample. Psychiatry Res 257, 111–117 (2017).

13. Nenadić, I. et al. Subclinical schizotypal vs. autistic traits show overlapping and diametrically opposed facets in a non-clinical population. Schizophr Res 231, 32–41 (2021).

14. Fagel, S. S. A. A. et al. Development of schizotypal symptoms following psychiatric disorders in childhood or adolescence. Eur Child Adolesc Psychiatry 22, 683–692 (2013).

15. Crespi, B., Dinsdale, N., Read, S. & Hurd, P. Spirituality, dimensional autism, and schizotypal traits: The search for meaning. PLOS ONE 14, e0213456 (2019).

16. Debbané, M., Glaser, B., David, M. K., Feinstein, C. & Eliez, S. Psychotic symptoms in children and adolescents with 22q11.2 deletion syndrome: Neuropsychological and behavioral implications. Schizophrenia Research 84, 187–193 (2006).

17. Stoddard, J., Niendam, T., Hendren, R., Carter, C. & Simon, T. J. Attenuated positive symptoms of psychosis in adolescents with chromosome 22q11.2 deletion syndrome. Schizophrenia Research 118, 118– 121 (2010).

18. Modinos, G. et al. Neuroanatomical changes in people with high schizotypy: relationship to glutamate levels. Psychol Med 48, 1880–1889 (2018).

19. van Lutterveld, R. et al. Cortical thickness in individuals with non-clinical and clinical psychotic symptoms. Brain 137, 2664–2669 (2014).

20. DeRosse, P. et al. Evidence from structural and diffusion tensor imaging for frontotemporal deficits in psychometric schizotypy. Schizophr Bull 41, 104–114 (2015).

21. Derome, M. et al. Developmental Trajectories of Cortical Thickness in Relation to Schizotypy During Adolescence. Schizophr Bull (2020) doi:10.1093/schbul/sbaa020.

22. Ettinger, U. et al. Association between brain structure and psychometric schizotypy in healthy individuals. World J. Biol. Psychiatry 13, 544–549 (2012).

23. Modinos, G. et al. Schizotypy and brain structure: a voxel-based morphometry study. Psychol Med 40, 1423–1431 (2010).

24. Nenadic, I. et al. Brain structural correlates of schizotypy and psychosis proneness in a non-clinical healthy volunteer sample. Schizophr. Res. 168, 37–43 (2015).

25. Meller, T. et al. Brain structural correlates of schizotypal signs and subclinical schizophrenia nuclear symptoms in healthy individuals. Psychol Med 1–10 (2020) doi:10.1017/S0033291720002044.

26. Kirschner, M. et al. Ventral Striatal Dysfunction and Symptom Expression in Individuals With Schizotypal Personality Traits and Early Psychosis. Schizophr Bull (2016) doi:10.1093/schbul/sbw142.

27. Kirschner, M. et al. Orbitofrontal-Striatal Structural Alterations Linked to Negative Symptoms at Different Stages of the Schizophrenia Spectrum. Schizophrenia Bulletin (2020) doi:10.1093/schbul/sbaa169.

28. Sabaroedin, K. et al. Functional Connectivity of Corticostriatal Circuitry and Psychosis-like Experiences in the General Community. Biol. Psychiatry 86, 16–24 (2019).

29. Kirschner, M. et al. Deficits in context-dependent adaptive coding in early psychosis and healthy individuals with schizotypal personality traits. Brain 141, 2806–2819 (2018).

30. Tonini, E., Quidé, Y., Kaur, M., Whitford, T. J. & Green, M. J. Structural and functional neural correlates of schizotypy: A systematic review. Psychological Bulletin 147, 828–866 (2021).

31. Kirschner, M. et al. Cortical and subcortical neuroanatomical signatures of schizotypy in 3004 individuals assessed in a worldwide ENIGMA study. Mol Psychiatry (2021) doi:10.1038/s41380-021-01359-9.

32. van Erp, T. G. M. et al. Cortical Brain Abnormalities in 4474 Individuals With Schizophrenia and 5098 Control Subjects via the Enhancing Neuro Imaging Genetics Through Meta Analysis (ENIGMA) Consortium. Biol. Psychiatry 84, 644–654 (2018).

33. Opel, N. et al. Cross-Disorder Analysis of Brain Structural Abnormalities in Six Major Psychiatric Disorders: A Secondary Analysis of Mega- and Meta-analytical Findings From the ENIGMA Consortium. Biological Psychiatry 0, (2020).

34. Park, B. et al. Multiscale neural gradients reflect transdiagnostic effects of major psychiatric conditions on cortical morphology. Commun Biol 5, 1–14 (2022).

35. Kirschner, M. et al. Schizophrenia Polygenic Risk During Typical Development Reflects Multiscale Cortical Organization. Biological Psychiatry: Global Open Science 0, (2022).

36. Hansen, J. Y. et al. Local molecular and global connectomic contributions to cross-disorder cortical abnormalities. Nat Commun 13, 4682 (2022).

37. Hansen, J. Y. et al. Mapping neurotransmitter systems to the structural and functional organization of the human neocortex. Nat Neurosci 25, 1569–1581 (2022).

38. Hettwer, M. D. et al. Coordinated cortical thickness alterations across six neurodevelopmental and psychiatric disorders. Nat Commun 13, 6851 (2022).

39. Wannan, C. M. J. et al. Evidence for Network-Based Cortical Thickness Reductions in Schizophrenia. Am J Psychiatry 176, 552–563 (2019).

40. Georgiadis, F. et al. Connectome architecture shapes large-scale cortical alterations in schizophrenia: a worldwide ENIGMA study. Mol Psychiatry 1–13 (2024) doi:10.1038/s41380-024-02442-7.

41. Crossley, N. A. et al. The hubs of the human connectome are generally implicated in the anatomy of brain disorders. Brain 137, 2382–2395 (2014).

42. Fornito, A., Zalesky, A. & Breakspear, M. The connectomics of brain disorders. Nature Reviews Neuroscience 16, 159–172 (2015).

43. Shafiei, G. et al. Spatial Patterning of Tissue Volume Loss in Schizophrenia Reflects Brain Network Architecture. Biological Psychiatry 87, 727–735 (2020).

44. Chopra, S. et al. Network-Based Spreading of Gray Matter Changes Across Different Stages of Psychosis. JAMA Psychiatry 80, 1246–1257 (2023).

45. Zhou, J., Gennatas, E. D., Kramer, J. H., Miller, B. L. & Seeley, W. W. Predicting Regional Neurodegeneration from the Healthy Brain Functional Connectome. Neuron 73, 1216–1227 (2012).

46. van den Heuvel, M. P. & Sporns, O. A cross-disorder connectome landscape of brain dysconnectivity. Nat Rev Neurosci 20, 435–446 (2019).

47. ENIGMA Clinical High Risk for Psychosis Working Group. Association of Structural Magnetic Resonance Imaging Measures With Psychosis Onset in Individuals at Clinical High Risk for Developing Psychosis: An ENIGMA Working Group Mega-analysis. JAMA Psychiatry (2021) doi:10.1001/jamapsychiatry.2021.0638.

48. van Rooij, D. et al. Cortical and Subcortical Brain Morphometry Differences Between Patients With Autism Spectrum Disorder and Healthy Individuals Across the Lifespan: Results From the ENIGMA ASD Working Group. Am J Psychiatry 175, 359–369 (2018).

49. Hoogman, M. et al. Brain Imaging of the Cortex in ADHD: A Coordinated Analysis of Large-Scale Clinical and Population-Based Samples. AJP 176, 531–542 (2019).

50. Sun, D. et al. Large-scale mapping of cortical alterations in 22q11.2 deletion syndrome: Convergence with idiopathic psychosis and effects of deletion size. Molecular Psychiatry 1–13 (2018) doi:10.1038/s41380-018-0078-5.

51. Desikan, R. S. et al. An automated labeling system for subdividing the human cerebral cortex on MRI scans into gyral based regions of interest. NeuroImage 31, 968–980 (2006).

52. Fischl, B. FreeSurfer. Neuroimage 62, 774–781 (2012).

53. Fischl, B. et al. Whole Brain Segmentation: Automated Labeling of Neuroanatomical Structures in the Human Brain. Neuron 33, 341–355 (2002).

54. Benjamini, Y. & Hochberg, Y. Controlling the False Discovery Rate: A Practical and Powerful Approach to Multiple Testing. Journal of the Royal Statistical Society. Series B (Methodological) 57, 289–300 (1995).

55. Yeo, B. T. T. et al. The organization of the human cerebral cortex estimated by intrinsic functional connectivity. J. Neurophysiol. 106, 1125–1165 (2011).

56. Mesulam, M. M. From sensation to cognition. Brain 121, 1013–1052 (1998).

57. Alexander-Bloch, A. F. et al. On testing for spatial correspondence between maps of human brain structure and function. Neuroimage 178, 540–551 (2018).

58. Burt, J. B. et al. Hierarchy of transcriptomic specialization across human cortex captured by structural neuroimaging topography. Nat. Neurosci. 21, 1251–1259 (2018).

59. Seidlitz, J. et al. Transcriptomic and cellular decoding of regional brain vulnerability to neurogenetic disorders. Nature Communications 11, 3358 (2020).

60. Hawrylycz, M. J. et al. An anatomically comprehensive atlas of the adult human brain transcriptome. Nature 489, 391–399 (2012).

61. Hansen, J. Y. et al. Mapping gene transcription and neurocognition across human neocortex. Nat Hum Behav 5, 1240–1250 (2021).

62. Avena-Koenigsberger, A., Misic, B. & Sporns, O. Communication dynamics in complex brain networks. Nat Rev Neurosci 19, 17–33 (2018).

63. Van Essen, D. C. et al. The Human Connectome Project: a data acquisition perspective. Neuroimage 62, 2222–2231 (2012).

64. Larivière, S. et al. Network-based atrophy modeling in the common epilepsies: A worldwide ENIGMA study. Science Advances 6, eabc6457 (2020).

65. Yau, Y. et al. Network connectivity determines cortical thinning in early Parkinson’s disease progression. Nature Communications 9, 12 (2018).

66. Flückiger, R. et al. Psychosis-predictive value of self-reported schizotypy in a clinical high-risk sample. Journal of Abnormal Psychology 125, 923–932 (2016).

67. Rodrigue, A. L. et al. Multimodal Neuroimaging Summary Scores as Neurobiological Markers of Psychosis. Schizophrenia Bulletin sbad149 (2023) doi:10.1093/schbul/sbad149.

68. Kochunov, P. et al. Translating ENIGMA schizophrenia findings using the regional vulnerability index: Association with cognition, symptoms, and disease trajectory. Hum Brain Mapp 43, 566–575 (2022).

69. Karcher, N. R., Modi, H., Kochunov, P., Gao, S. & Barch, D. M. Regional Vulnerability Indices in Youth With Persistent and Distressing Psychoticlike Experiences. JAMA Network Open 6, e2343081 (2023).

70. Sydnor, V. J. et al. Neurodevelopment of the association cortices: Patterns, mechanisms, and implications for psychopathology. Neuron 109, 2820–2846 (2021).

71. Hettwer, M. D. et al. Longitudinal trajectories of resilient psychosocial functioning link to ongoing cortical myelination and functional reorganization during adolescence. (2024) doi:10.31234/osf.io/2dv68.

72. Keshavan, M. et al. Psychopathology among offspring of parents with schizophrenia: Relationship to premorbid impairments. Schizophrenia Research 103, 114–120 (2008).

73. Keshavan, M. S., Sujata, M., Mehra, A., Montrose, D. M. & Sweeney, J. A. Psychosis proneness and ADHD in young relatives of schizophrenia patients. Schizophrenia Research 59, 85–92 (2003).

74. Ettinger, U., Joober, R., De Guzman, R. & O’driscoll, G. A. Schizotypy, attention deficit hyperactivity disorder, and dopamine genes. Psychiatry and Clinical Neurosciences 60, 764–767 (2006).

75. Louzolo, A. et al. Delusion-proneness displays comorbidity with traits of autistic-spectrum disorders and ADHD. PLOS ONE 12, e0177820 (2017).

76. Monks, S. et al. Further evidence for high rates of schizophrenia in 22q11.2 deletion syndrome. Schizophr Res 153, 231–236 (2014).

77. Bassett, A. S. & Chow, E. W. C. 22q11 Deletion Syndrome: A Genetic Subtype of Schizophrenia. Biol Psychiatry 46, 882–891 (1999).

78. Writing Committee for the Attention-Deficit/Hyperactivity Disorder et al. Virtual Histology of Cortical Thickness and Shared Neurobiology in 6 Psychiatric Disorders. JAMA Psychiatry 78, 47–63 (2021).

79. Margulies, D. S. et al. Situating the default-mode network along a principal gradient of macroscale cortical organization. Proceedings of the National Academy of Sciences 113, 12574–12579 (2016).

80. Paquola, C. et al. Microstructural and functional gradients are increasingly dissociated in transmodal cortices. PLOS Biology 17, e3000284 (2019).

81. Esbenshade, T. A. et al. The histamine H3 receptor: an attractive target for the treatment of cognitive disorders. Br J Pharmacol 154, 1166–1181 (2008).

82. Provensi, G., Costa, A., Izquierdo, I., Blandina, P. & Passani, M. B. Brain histamine modulates recognition memory: possible implications in major cognitive disorders. Br J Pharmacol 177, 539–556 (2020).

83. Jin, C. Y., Anichtchik, O. & Panula, P. Altered histamine H3 receptor radioligand binding in post-mortem brain samples from subjects with psychiatric diseases. Br J Pharmacol 157, 118–129 (2009).

84. Arumuham, A. et al. The histamine system and cognitive function: An in vivo H3 receptor PET imaging study in healthy volunteers and patients with schizophrenia. J Psychopharmacol 37, 1011–1022 (2023).

85. Saxena, S. & Caroni, P. Selective neuronal vulnerability in neurodegenerative diseases: from stressor thresholds to degeneration. Neuron 71, 35–48 (2011).

86. Zheng, Y.-Q. et al. Local vulnerability and global connectivity jointly shape neurodegenerative disease propagation. PLOS Biology 17, e3000495 (2019).

87. Buckner, R. L. et al. Cortical hubs revealed by intrinsic functional connectivity: mapping, assessment of stability, and relation to Alzheimer’s disease. J Neurosci 29, 1860–1873 (2009).

88. Haan, W. de, Mott, K., Straaten, E. C. W. van, Scheltens, P. & Stam, C. J. Activity Dependent Degeneration Explains Hub Vulnerability in Alzheimer’s Disease. PLOS Computational Biology 8, e1002582 (2012).

89. Seeley, W. W., Crawford, R. K., Zhou, J., Miller, B. L. & Greicius, M. D. Neurodegenerative diseases target large-scale human brain networks. Neuron 62, 42–52 (2009).

90. de Lange, S. C. et al. Shared vulnerability for connectome alterations across psychiatric and neurological brain disorders. Nat Hum Behav 3, 988–998 (2019).

91. Kemp, K. C., Bathery, A. J., Barrantes-Vidal, N. & Kwapil, T. R. Positive, Negative, and Disorganized Schizotypy Predict Differential Patterns of Interview-Rated Schizophrenia-Spectrum Symptoms and Impairment: Assessment (2020) doi:10.1177/1073191119900008.

92. Thompson, P. M. et al. ENIGMA and global neuroscience: A decade of large-scale studies of the brain in health and disease across more than 40 countries. Transl Psychiatry 10, 1–28 (2020).

93. Bethlehem, R. a. I., et al. Brain charts for the human lifespan. Nature 604, 525–533 (2022).

94. Elam, J. S. et al. The Human Connectome Project: A retrospective. Neuroimage 244, 118543 (2021).

95. Garavan, H. et al. Recruiting the ABCD sample: Design considerations and procedures. Developmental Cognitive Neuroscience 32, 16–22 (2018).

96. Chapman, L. J., Chapman, J. P. & Raulin, M. L. Scales for physical and social anhedonia. J Abnorm Psychol 85, 374–82 (1976).

97. Chapman, L. J., Edell, W. S. & Chapman, J. P. Physical anhedonia, perceptual aberration, and psychosis proneness. Schizophrenia Bulletin 6, 639–653 (1980).

98. Stefanis, N. C. et al. Evidence that three dimensions of psychosis have a distribution in the general population. Psychol Med 32, 347–58 (2002).

99. Raine, A. The SPQ: a scale for the assessment of schizotypal personality based on DSM-III-R criteria. Schizophr Bull 17, 555–564 (1991).

100. Mason, O., Claridge, G. & Jackson, M. New scales for the assessment of schizotypy. Personality and Individual Differences 18, 7–13 (1995).

101. Rust, J. The Rust Inventory of Schizoid Cognitions (RISC): a psychometric measure of psychoticism in the normal population. Br J Clin Psychol 26, 151–152 (1987).

102. van Erp, T. G. M. et al. Subcortical brain volume abnormalities in 2028 individuals with schizophrenia and 2540 healthy controls via the ENIGMA consortium. Molecular Psychiatry 21, 547–553 (2016).

103. Kim, S. ppcor: An R Package for a Fast Calculation to Semi-partial Correlation Coefficients. Commun Stat Appl Methods 22, 665–674 (2015).

104. Viechtbauer, W. Conducting Meta-Analyses in R with the metafor Package. Journal of Statistical Software 36, 1–48 (2010).

105. Larivière, S. et al. The ENIGMA Toolbox: multiscale neural contextualization of multisite neuroimaging datasets. Nat Methods 18, 698–700 (2021).

106. Markello, R. D. & Misic, B. Comparing spatial null models for brain maps. NeuroImage 236, 118052 (2021).

107. Markello, R., Shafiei, G., Zheng, Y.-Q. & Mišić, B. abagen: A toolbox for the Allen Brain Atlas genetics data. Zenodo 10.5281/zenodo.3726257 (2020).

108. Vaishnavi, S. N. et al. Regional aerobic glycolysis in the human brain. Proc Natl Acad Sci U S A 107, 17757–17762 (2010).

109. Van Essen, D. C. et al. The WU-Minn Human Connectome Project: An Overview. Neuroimage 80, 62–79 (2013).

110. Glasser, M. F. et al. The minimal preprocessing pipelines for the Human Connectome Project. Neuroimage 80, 105–124 (2013).

111. Budescu, D. V. Dominance analysis: A new approach to the problem of relative importance of predictors in multiple regression. Psychological Bulletin 114, 542–551 (1993).

112. Azen, R. & Budescu, D. V. The dominance analysis approach for comparing predictors in multiple regression. Psychol Methods 8, 129–148 (2003).

