## Supplement for "Multiscale characterization of cortical signatures in positive and negative schizotypy: A worldwide ENIGMA study"

### Table S1. Demographics sites included in positive schizotypy analysis

| Site | N | Scale 1 | Scale 2 | %Male | Mean Age | Age SD (years) | Score 1 Mean | Score 1 SD | Score 2 Mean | Score 2 SD |
| --- | --- | --- | --- | --- | --- | --- | --- | --- | --- | --- |
| ASRB | 191 | SPQ-CP |  | 47.64 | 39.69 | 13.77 | 3 | 3.9 |  |  |
| AUK | 49 | OLIFE-UE |  | 34.69 | 23.33 | 4.5 | 12.51 | 6.84 |  |  |
| BEIJ-ECNU | 70 | CHAPMAN-PA |  | 45.71 | 19.89 | 1.16 | 14.28 | 6.42 |  |  |
| BEIJ-GZ | 76 | CHAPMAN-PA |  | 48.68 | 19.28 | 0.9 | 15.62 | 9.61 |  |  |
| BONN1 | 126 | SPQ-CP |  | 51.97 | 27 | 7.84 | 3 | 2.5 |  |  |
| BONN2 | 51 | RISC |  | 64.71 | 27.2 | 5.36 | 22.37 | 7.71 |  |  |
| BONN3 | 31 | OLIFE-UE |  | 43.75 | 24 | 4.52 | 5 | 3.73 |  |  |
| CAM | 89 | SPQ-CP |  | 46.07 | 24.07 | 5.3 | 5.54 | 4.93 |  |  |
| NYC | 240 | CAPE-POS | SPQ-CP | 51.33 | 35.99 | 12.94 | 22.67 | 3.14 | 1.89 | 3.05 |
| FIDMAG | 64 | RISC |  | 42.19 | 37.89 | 10.29 | 22.5 | 9.91 |  |  |
| GENEVA | 114 | SPQ-CP |  | 54.63 | 16.43 | 2.63 | 8.49 | 6.87 |  |  |
| GRON1 | 58 | PANSS-POS |  | 73.13 | 34.23 | 10.99 | 14.35 | 4.29 |  |  |
| GRON2 | 43 | SPQ-CP |  | 62.79 | 22.67 | 2.26 | 7.59 | 5.91 |  |  |
| IGP | 63 | SPQ-CP |  | 55.56 | 36.43 | 11.05 | 4.05 | 5.03 |  |  |
| LOND1a | 46 | SPQ-CP | OLIFE-UE | 52.17 | 27.46 | 6.84 | 5.3 | 6.67 | 6.02 | 6.4 |
| LOND1b | 40 | CAPE-POS |  | 52.5 | 20.8 | 3.76 | 1.44 | 0.34 |  |  |
| LOND2 | 44 | OLIFE-UE |  | 27.27 | 29.41 | 9.2 | 3.27 | 3.44 |  |  |
| LOND3 | 40 | SPQ-CP |  | 60 | 30.97 | 10.87 | 2.45 | 2.07 |  |  |
| LOND4 | 26 | CAPE-POS |  | 65.38 | 38.62 | 7.34 | 28.15 | 6.27 |  |  |
| FOR-2107-MR | 446 | SPQ-CP |  | 37.44 | 34.67 | 12.85 | 0.83 | 0.89 |  |  |
| MELB | 376 | CAPE-POS | OLIFE-UE | 43.35 | 23.39 | 5.15 | 25.85 | 4.68 | 2.98 | 2.65 |
| MTL | 40 | CHAPMAN-PA |  | 28 | 19.97 | 1.25 | 13.6 | 9.28 |  |  |
| MOSC | 107 | SPQ-CP |  | 48.6 | 26.46 | 6.61 | 6.18 | 4.67 |  |  |
| MNC | 173 | SPQ-CP |  | 46.82 | 43.31 | 11.85 | 1.46 | 1.45 |  |  |
| FOR-2017-MS | 228 | SPQ-CP |  | 35.96 | 28.42 | 10.41 | 0.7 | 0.83 |  |  |
| PARIS | 52 | SPQ-CP |  | 42.31 | 35.85 | 11.24 | 1.32 | 2.23 |  |  |
| ZUR1 | 27 | SPQ-CP |  | 62.96 | 29.15 | 10.35 | 17.56 | 5.01 |  |  |
| ZUR2 | 62 | SPQ-CP |  | 100 | 27.74 | 4.74 | 8.76 | 6.45 |  |  |

### Table S2. Demographics, sites included in negative schizotypy analysis

| Site | N | Scale 1 | Scale 2 | % male | Mean age | Age SD | Score 1 Mean | Score 1 SD | Score 2 Mean | Score 2 SD |
| --- | --- | --- | --- | --- | --- | --- | --- | --- | --- | --- |
| AMS | 49 | CAPE |  | 53.06 | 21.36 | 3.08 | 45.51 | 9.90 |  |  |
| ASRB | 191 | SPQ |  | 52.36 | 39.69 | 13.77 | 5.23 | 5.46 |  |  |
| AUK | 49 | OLIFE |  | 34.69 | 23.33 | 4.50 | 7.73 | 5.66 |  |  |
| BEIJ | 146 | SPQ | CAP | 47.26 | 19.57 | 1.08 | 18.56 | 10.08 | 9.97 | 7.54 |
| BONN1 | 93 | SPQ |  | 50.54 | 28.08 | 8.02 | 2.81 | 2.93 |  |  |
| CAM | 88 | SPQ |  | 45.45 | 24.33 | 4.72 | 5.45 | 4.41 |  |  |
| FOR-2017-MS | 228 | SPQB |  | 35.96 | 28.42 | 10.41 | 1.49 | 1.65 |  |  |
| FOR-2107-MR | 446 | SPQB |  | 37.44 | 34.67 | 12.85 | 1.78 | 1.74 |  |  |
| GENEVA | 114 | SPQ |  | 52.63 | 15.94 | 2.64 | 6.04 | 4.31 |  |  |
| GRON1 | 35 | CAPE-Adj |  | 51.43 | 36.11 | 9.31 | 6.41 | 1.29 |  |  |
| GRON2 | 42 | SPQB |  | 61.9 | 22.67 | 2.26 | 5.55 | 4.43 |  |  |
| IGP | 63 | SPQB |  | 55.56 | 36.43 | 11.05 | 6.44 | 6.00 |  |  |
| LOND1a | 46 | SPQB | OLIFE | 52.17 | 27.48 | 6.83 | 6.18 | 5.41 | 13.09 | 3.02 |
| LOND3 | 40 | SPQB |  | 60 | 30.97 | 10.87 | 3.45 | 2.32 |  |  |
| LOND4 | 26 | CAPE |  | 65.38 | 38.62 | 7.34 | 23.88 | 5.35 |  |  |
| MELB | 397 | CAPE |  | 43.32 | 23.42 | 5.17 | 23.81 | 5.71 |  |  |
| MNC | 173 | SPQB |  | 46.82 | 43.31 | 11.85 | 3.34 | 3.08 |  |  |
| MOSC | 58 | SPQ |  | 51.72 | 25.66 | 5.13 | 6.93 | 6.35 |  |  |
| MTL | 40 | CHAP |  | 32.5 | 19.83 | 1.14 | 15.70 | 11.28 |  |  |
| NYC | 225 | CAPE-Adj |  | 51.11 | 35.92 | 12.85 | 1.25 | 0.26 |  |  |
| UTR | 91 | SPQ |  | 37.36 | 40.92 | 14.26 | 5.82 | 6.50 |  |  |
| ZUR1 | 27 | SPQ |  | 62.96 | 29.15 | 10.35 | 18.15 | 5.40 |  |  |
| ZUR2 | 62 | SPQ | CHAP | 100 | 27.74 | 4.74 | 7.03 | 4.95 | 10.90 | 5.78 |

### Table S3. Scanner Details

| **Sample** | **Number of Scanners** | **Scanner Vendor & Type** | **Imaging Protocols** | **Slice Orientation** | **FreeSurfer Version** | **Operating System** | **Number of subjects removed after QC** |
| --- | --- | --- | --- | --- | --- | --- | --- |
| AMS | 1 | 3T Phillips Achieva | 3D T1-weighted images (TR = 8.2, TE = 3.8, FA = 8°, FOV 240 × 188 mm, voxel size 1 × 1 × 1, 220 slices) | sequential ascending | 6.0.1 | virtual linux environment (sigularity on HPC) | 0 |
| ASRB | 5 | 1.5T Siemens Avanto | 3D MPRAGE; TR 1980ms, TE 4.3ms, field-of-view 250 x 250 mm2, data acquisition matrix 256 x 256, 176 contiguous 1mm slices; voxel size 0.98 x 0.98 x 1.0 mm3, flip angle 15° | sagittal | 5.1 | Mac OS X 10.9.5 | 0 |
| AUCK | 1 | 3T Siemens | 3D MPRAGE; FOV = 256 mm2; matrix size = 256x 256 mm; number of slices = 176; slice thickness = 1 mm; voxel-size = 1 x1 x1 mm3; TE/TR/flip angle = 2.07 ms/1900 ms/9 deg; GRAPPA acceleration factor = 2; TA = 4.26 min. | interleaved | 5.3.0 | linux | 0 |
| BEIJ | 2 | 3T Siemens | site 1: MPRAGE; TR 2530ms, TE 2.34ms, FOV 256mm, matrix 256x256, 192 slices, slice thickness 1mm, flip angle 7; site 2: MPRAGE; TR 2530ms, TE 2.34ms, FOV 256mm, matrix 256x256, 172 slices, slice thickness 1mm, flip angle 7; | sagittal | 6.0.0 | Linux | 0 |
| BONN_3 | 1 | 3T Siemens Trio | TR = 1570 ms; TE = 3.42 ms; inversion time (TI) = 800 ms; flip angle = 15°; FoV = 256 mm; matrix size = 256 x 256; 160 slices; slice thickness = 1 mm; voxel size = 1 x 1 x 1 | sequential | 6.0.0 |  | 0 |
| BONN_2 | 2 | 1.5 T GE Signa Advantage, 3T Siemens Magnetom Verio | GE Signa Advantage: 3D-SPGR voxel resolution: 1 x 1 x 1.5 mm; repetition time (TR): 18 ms; inversion time (TI): 450 ms; echo time (TE): 5.1 ms; bandwidth: 15.63 kHz |  | 6.0.0 |  | 0 |
| BONN1 | 1 | 3T Siemens Magnetom Verio | 3D MPRAGE sequence, repetition time of TR = 2400 ms, echo time TE = 3.06 ms, flip angle = 9 degrees with 160 slices, slice thickness = 1.0 mm, voxel size = 1.0 × 1.0 × 1.0 mm, field of view FOV = 256 mm |  | 6.0.0 |  | 0 |
| CAM | 1 | 3T Siemens Trio | MPRAGE; TR/TE 2.98/2300 ms, 1 × 1 voxels, slice thickness 1 mm, flip angle 9°, FOV 24 × 25.6 mm, 176 slices |  | 6.0.0 | Unix | 0 |
| FIDMAG | 1 | 1.5T GE Signa Excite | 3D TR=12.356 ms, TE=5.192 ms, inversion time=450 ms, acquisition matrix=256 x 224, voxel size=1*1*1 mm^3, flip angle=20°, 180 slices | axial | 7.1.1 | Linux | 0 |
| FOR2107-MR | 1 | 3T Siemens Magnetom TiroTim syngo | 3D T1-weighted magnetization prepared rapid acquisition gradient echo (MPRAGE); TR=1900ms, TE=2.26ms, TI=900ms, FA=9°, voxel size=1.0x1.0x1.0mm³, Acquisition Direction Sagittal, 176 slices, slice gap 0.5mm. | Sagittal | 5.3 | Red Hat Enterprise Linux Server release 5.11 (Tikanga) | 0 |
| FOR2107-MS | 1 | 3T Siemens PRISMA | 3D T1-weighted magnetization prepared rapid acquisition gradient echo (MPRAGE); TR=2130ms, TE=2.28ms, TI=900ms, FA=8°, voxel size=1.0x1.0x1.0mm³, Acquisition Direction Sagittal, 192 slices, no slice gap. | Sagittal | 5.3 | Red Hat Enterprise Linux Server release 5.11 (Tikanga) | 0 |
| GENEVA | 2 | 3T Siemens Trio | T1-Weighted images (192 slices, TR=2500ms, TE=3ms, flip angle= 8 degree, slice thickness=1.1mm, FOV=22cm, Acq matrix 256x256) | A >> P or descending | 6.0.0 | Mac OS SIERRA v 10.12.6 | 0 |
| GRON1 | 1 | 3T Phillips | a T1-weighted image was obtained (TR/TE = 9/3.5 ms) using fast-field echo and turbo-field echo: 170 axial slices; FOV (rl, ap, fh) = 232 × 170 × 256 mm; flip angle = 8°, voxel size = 1 × 1 × 1 mm, slice thickness = 1 mm. | descending | 6.0.1 | virtual linux environment (sigularity on HPC) | 0 |
| GRON2 | 1 | 3T Phillips | anatomic images were obtained using a sagittal 3-dimensional T1-weighted sequence (176 slices, repetition time 9 ms, echo time 3.5 ms, field of view 256 mm, voxel size 1 × 1 × 1 mm, slice thickness 1.0 mm) using an 8 channel head coil. | descending | 6.0.1 | virtual linux environment (sigularity on HPC) | 0 |
| IGP | 1 | 3T Phillips Achieva TX | 3D MPRAGE; TR 8.9ms, TE 4.1ms, field of view 240mm, matrix 268 x 268, 200 slices, slice thickness 0.9mm, no gap | sagittal | 5.3 | Mac OS X 10.9.5 | 0 |
| JENA1 | 1 | 3T Siemens Tim Trio | 3D T1-weighted MPRAGE, TR: 2300ms, TE: 3.03 ms, flip angle: 9°, 192 slices, field of view 256mm, voxel resolution 1x1x1mm, acquisition time: 5:21min | sagittal | 6.0.0 | Linux | 0 |
| LOND1a | 1 | 3T GE Discovery MR750 | 3D T1-weighted inversion recovery prepared gradient echo sequence (voxel size: 1.05 × 1.05 × 1.2 mm, field of view: 270 mm, 196 slices, TR: 7.3 ms, TE: 3.0 ms, inversion time: 400 ms, flip angle = 11°), based on the well-validated ADNI 2/ADNI GO protocols (see http://adni.loni.usc.edu/methods/documents/mri-protocols/). | sequential (top down) | 6.0.0 | Unix | 0 |
| LOND1b | 1 | 3T Phillips Intera | T1-weighted 3D fast-field echo (FFE) sequence [repetition time (TR)=25 ms, echo time (TE)=4.6 ms, field of view (FOV)=260 mm, matrix=256x256, 160 contiguous axial slices of 1-mm thickness, voxel size= 1x1x1 mm]. | interleaved | 6.0.0 | Unix | 0 |
| LOND2 | 2 | 1.5 T GE, 3T GE | 1.5 T GE: 3D, TR=11.1 ms, TE=4.9 ms, inversion time=300 ms, acquisition matrix=256 x 160, 150 locations, slice thickness=1.1 mm, in-plane resolution=1.094 mm, flip angle=18°; 3 T GE: TR: 7.3 ms, TE: 3.0 ms, inversion time: 400 ms, voxel size: 1.05 x1.05 x1.2mm, field of view: 270mm, 196 slices, flip angle = 11° |  | 6.0.0 | Unix | 0 |
| LOND3 | 1 | 3T Siemens | MPRAGE; TR 2000ms; 1mm x 1mm 1mm. 256 x 256 x 176 slices, flip angle 11, slice thickness 1mm, TE = 2.07ms | interleaved | 6.0.0 | Mac | 0 |
| LOND4 | 1 | 3T GE | T1-weighted image (196 slices; isotropic voxels of 1.2 mm; TR 7.312 ms; TE 3.016 ms;flip angle: 11°; FOV 270 mm) | sequential (top down) | 6.0.0 | Unix | 0 |
| LOND5 | 1 | 3T Siemens | MPRAGE; TR 1900ms; 1mm x 1mm 1mm voxel size; in plane resolution of 256 x 256 x 176 slices, flip angle 11, slice thickness 1mm, TE = 2.07ms | interleaved | 6.0.0 | Linux Ubuntu Bionic | 0 |
| MELB | 1 | 3T Siemens | 3D magnetic-prepared rapid gradient echo sequence. A total of 192 slices were acquired for each participant’s T1-weighted images using an ascending acquisition with the following parameters: TR of 2300 ms, TE of 2.07 ms, flip angle of 9°, FOV of 256mm, and voxel size of 1mm3. | sagittal | 5.3.0 | Linux | 34 |
| MNC | 1 | 3T scanner Gyroscan Intera, Philips Medical Systems | 3D fast gradient echo sequence (turbo field echo), TR = 7.4 msec, TE = 3.4 msec, Flip Angle = 9°, two signal averages, inversion prepulse every 814.5 msec, acquired over a FOV of 256 (feet-head [FH]) × 204 (anterior-posterior [AP]) × 160 (right-left [RL]) mm, phase encoding in AP and RL direction, reconstructed to cubic voxels of .5 × .5 × .5 mm | Sagittal | 5.3 | Red Hat Enterprise Linux Server release 5.11 (Tikanga) | 0 |
| MOSC | 1 | 3 T Phillips Ingenia | 3D T1-weighted TFE; TR=7.9 ms, TE=3.5 ms; flip angle 8; number of slices=170; voxel size=0.98 x 0.98 x 1.0, no gap | axial | 5.3 | LinuxCentOS 6.10 | 0 |
| MTL | 1 | 3T Siemens Trio | 3D T1-weighted images were acquired with a gradient echo T1-weighted sequence, voxel size 1x1x1 mm3. TR=1900ms; TE=4.9ms; FA=25; matrix 176x256x256 | sagittal | 6.0.0 | Unix | 0 |
| NYC (Hillside) | 1 | 3T GE | 3D-SPGR images using a 1mm thick slice acquisition with image parameters: TR = 7.5 ms, TE = 3 ms, matrix = 256x256, FOV = 240 mm, 216 contiguous images. | interleaved | 6.0.0 | Linux | 0 |
| PARIS | 1 | 3T Siemens Trio | 3D T1 TE, 2.98 ms; TR, 2300 ms; 160 sections; voxel size, 1.0 × 1.0 × 1.1 mm |  |  |  |  |
| UTR | 1 | 3T Phillips Achieva | T1-weighted images were acquired with a 3D T1 turbo field-echo sequence. The acquisition protocol parameters were: 160 slices; repetition time = 9.96 ms; echo time = 4.59 ms; flip angle 8°; 1 mm slice thickness with no inter-slice gap; matrix = 256 × 256 and field of view 224 mm, achieving a voxel size of 0.875 × 0.875 × 1 mm. Scan time was 8 min and 50 s | saggital | 5.1.0 | Unix | 0 |
| ZUR1 | 1 | 3T Phillips | 3D T1-weighted images were acquired with an ultrafast gradient echo T1-weighted sequence (TR=8.4ms, TE=3.8ms, flip angle=8°) in 160 sagittal plan slices (1mm slice thickness, no slice gap) of 240×240mm2 resulting in 1x1x1mm3 voxels. | sagittal | 6.0.0 | Unix | 0 |
| ZUR2 | 1 | 3T Phillips Achieva | 3D T1-weighted images; 160 slices; TR, 8.2 ms; TE, 3.8 ms; flip angle, 8°; spatial resolution, 1 × 1 × 1 mm3; FOV = 160 × 240 mm2 | sagittal | 6.0.0 | OS X | 0 |

### Table S4. Questionnaires/Rating Scales used across sites

| Questionnaires | Total number of items | Factors/Subscales |
| --- | --- | --- |
| CAPE | 42 | Depression, Negative, Positive Symptoms |
| CHAP | 196 | Perceptual Aberration; Magical Ideation; Physical Anhedonia; Social Anhedonia; Impulsive Nonconformity |
| OLIFE | 104 | Unusual Experiences; Cognitive Disorganization;  Introvertive Anhedonia; Impulsive Nonconformity |
| PANSS | 30 | Positive; Negative; General symptoms |
| RISC | 26 | Positive and Cognitive content of schizotypy |
| SPQ | 74 | Cognitive/Perceptual; Interpersonal; Disorganized |
| SPQ-B | 22 | Cognitive/Perceptual; Interpersonal; Disorganized |

### Table S5. Association between CT and positive schizotypy corrected for age, and sex

| DKT Regions | Partial R | Std. Err. | 95% CI, lower bound | 95% CI, upper bound | p-value | FDR-pvalue | Sample size per region |
| --- | --- | --- | --- | --- | --- | --- | --- |
| L_bankssts_thickavg | -0.031 | 0.032 | -0.094 | 0.033 | 0.344 | 0.517 | 2539 |
| L_caudalanteriorcingulate_thickavg | 0.024 | 0.020 | -0.014 | 0.062 | 0.216 | 0.401 | 2704 |
| L_caudalmiddlefrontal_thickavg | -0.038 | 0.025 | -0.087 | 0.011 | 0.126 | 0.316 | 2711 |
| L_cuneus_thickavg | -0.045 | 0.026 | -0.096 | 0.007 | 0.092 | 0.271 | 2587 |
| L_entorhinal_thickavg | -0.021 | 0.028 | -0.076 | 0.035 | 0.467 | 0.615 | 2558 |
| L_fusiform_thickavg | -0.028 | 0.023 | -0.073 | 0.017 | 0.227 | 0.405 | 2676 |
| L_inferiorparietal_thickavg | -0.046 | 0.019 | -0.084 | -0.008 | 0.018 | 0.197 | 2634 |
| L_inferiortemporal_thickavg | 0.000 | 0.021 | -0.041 | 0.040 | 0.989 | 0.999 | 2651 |
| L_isthmuscingulate_thickavg | -0.026 | 0.019 | -0.063 | 0.012 | 0.175 | 0.380 | 2721 |
| L_lateraloccipital_thickavg | -0.028 | 0.019 | -0.065 | 0.010 | 0.151 | 0.351 | 2688 |
| L_lateralorbitofrontal_thickavg | -0.053 | 0.030 | -0.112 | 0.007 | 0.082 | 0.259 | 2715 |
| L_lingual_thickavg | -0.049 | 0.022 | -0.093 | -0.005 | 0.029 | 0.224 | 2672 |
| L_medialorbitofrontal_thickavg | 0.043 | 0.019 | 0.005 | 0.080 | 0.025 | 0.212 | 2689 |
| L_middletemporal_thickavg | 0.000 | 0.020 | -0.038 | 0.039 | 0.984 | 0.999 | 2546 |
| L_parahippocampal_thickavg | -0.004 | 0.037 | -0.078 | 0.069 | 0.909 | 0.992 | 2706 |
| L_paracentral_thickavg | -0.051 | 0.025 | -0.100 | -0.001 | 0.044 | 0.238 | 2714 |
| L_parsopercularis_thickavg | -0.071 | 0.019 | -0.109 | -0.034 | 0.000 | 0.018 | 2692 |
| L_parsorbitalis_thickavg | -0.063 | 0.019 | -0.101 | -0.026 | 0.001 | 0.043 | 2699 |
| L_parstriangularis_thickavg | -0.032 | 0.029 | -0.090 | 0.025 | 0.271 | 0.452 | 2685 |
| L_pericalcarine_thickavg | -0.042 | 0.026 | -0.093 | 0.009 | 0.106 | 0.287 | 2643 |
| L_postcentral_thickavg | -0.074 | 0.032 | -0.137 | -0.012 | 0.020 | 0.197 | 2665 |
| L_posteriorcingulate_thickavg | -0.005 | 0.025 | -0.055 | 0.045 | 0.838 | 0.952 | 2720 |
| L_precentral_thickavg | -0.060 | 0.025 | -0.109 | -0.010 | 0.018 | 0.197 | 2686 |
| L_precuneus_thickavg | -0.024 | 0.026 | -0.074 | 0.027 | 0.356 | 0.518 | 2710 |
| L_rostralanteriorcingulate_thickavg | 0.024 | 0.019 | -0.013 | 0.062 | 0.201 | 0.401 | 2695 |
| L_rostralmiddlefrontal_thickavg | -0.035 | 0.019 | -0.072 | 0.003 | 0.071 | 0.238 | 2700 |
| L_superiorfrontal_thickavg | -0.038 | 0.019 | -0.075 | 0.000 | 0.050 | 0.238 | 2699 |
| L_superiorparietal_thickavg | -0.051 | 0.027 | -0.104 | 0.001 | 0.056 | 0.238 | 2665 |
| L_superiortemporal_thickavg | -0.023 | 0.025 | -0.072 | 0.025 | 0.347 | 0.517 | 2529 |
| L_supramarginal_thickavg | -0.043 | 0.022 | -0.087 | 0.000 | 0.049 | 0.238 | 2584 |
| L_frontalpole_thickavg | 0.001 | 0.019 | -0.036 | 0.038 | 0.955 | 0.995 | 2726 |
| L_temporalpole_thickavg | 0.022 | 0.028 | -0.032 | 0.077 | 0.417 | 0.571 | 2679 |
| L_transversetemporal_thickavg | 0.009 | 0.031 | -0.052 | 0.070 | 0.766 | 0.896 | 2726 |
| L_insula_thickavg | -0.027 | 0.022 | -0.071 | 0.016 | 0.220 | 0.401 | 2673 |
| R_bankssts_thickavg | -0.002 | 0.019 | -0.040 | 0.036 | 0.921 | 0.992 | 2658 |
| R_caudalanteriorcingulate_thickavg | 0.009 | 0.019 | -0.029 | 0.046 | 0.646 | 0.798 | 2716 |
| R_caudalmiddlefrontal_thickavg | -0.040 | 0.024 | -0.086 | 0.006 | 0.089 | 0.271 | 2700 |
| R_cuneus_thickavg | -0.030 | 0.027 | -0.082 | 0.022 | 0.265 | 0.449 | 2613 |
| R_entorhinal_thickavg | 0.012 | 0.028 | -0.043 | 0.066 | 0.671 | 0.801 | 2472 |
| R_fusiform_thickavg | 0.000 | 0.019 | -0.038 | 0.038 | 0.999 | 0.999 | 2682 |
| R_inferiorparietal_thickavg | -0.033 | 0.027 | -0.086 | 0.020 | 0.217 | 0.401 | 2628 |
| R_inferiortemporal_thickavg | -0.028 | 0.019 | -0.066 | 0.010 | 0.148 | 0.351 | 2659 |
| R_isthmuscingulate_thickavg | -0.005 | 0.019 | -0.042 | 0.033 | 0.813 | 0.935 | 2711 |
| R_lateraloccipital_thickavg | -0.064 | 0.028 | -0.120 | -0.009 | 0.024 | 0.212 | 2677 |
| R_lateralorbitofrontal_thickavg | -0.023 | 0.038 | -0.096 | 0.051 | 0.542 | 0.678 | 2694 |
| R_lingual_thickavg | -0.041 | 0.035 | -0.109 | 0.027 | 0.242 | 0.417 | 2671 |
| R_medialorbitofrontal_thickavg | -0.006 | 0.020 | -0.046 | 0.034 | 0.770 | 0.896 | 2667 |
| R_middletemporal_thickavg | 0.020 | 0.030 | -0.039 | 0.078 | 0.506 | 0.649 | 2637 |
| R_parahippocampal_thickavg | -0.045 | 0.028 | -0.101 | 0.010 | 0.107 | 0.287 | 2708 |
| R_paracentral_thickavg | -0.037 | 0.020 | -0.077 | 0.002 | 0.060 | 0.238 | 2713 |
| R_parsopercularis_thickavg | 0.001 | 0.019 | -0.037 | 0.038 | 0.973 | 0.999 | 2666 |
| R_parsorbitalis_thickavg | -0.045 | 0.028 | -0.101 | 0.010 | 0.110 | 0.287 | 2695 |
| R_parstriangularis_thickavg | -0.043 | 0.025 | -0.093 | 0.006 | 0.083 | 0.259 | 2661 |
| R_pericalcarine_thickavg | -0.017 | 0.040 | -0.096 | 0.062 | 0.669 | 0.801 | 2625 |
| R_postcentral_thickavg | -0.068 | 0.024 | -0.114 | -0.021 | 0.004 | 0.086 | 2665 |
| R_posteriorcingulate_thickavg | 0.017 | 0.019 | -0.021 | 0.054 | 0.387 | 0.545 | 2721 |
| R_precentral_thickavg | -0.066 | 0.032 | -0.128 | -0.003 | 0.041 | 0.238 | 2668 |
| R_precuneus_thickavg | -0.027 | 0.022 | -0.071 | 0.016 | 0.221 | 0.401 | 2709 |
| R_rostralanteriorcingulate_thickavg | 0.026 | 0.032 | -0.036 | 0.088 | 0.408 | 0.567 | 2662 |
| R_rostralmiddlefrontal_thickavg | -0.027 | 0.019 | -0.064 | 0.011 | 0.167 | 0.370 | 2678 |
| R_superiorfrontal_thickavg | -0.031 | 0.019 | -0.068 | 0.006 | 0.104 | 0.287 | 2699 |
| R_superiorparietal_thickavg | -0.054 | 0.029 | -0.111 | 0.004 | 0.066 | 0.238 | 2676 |
| R_superiortemporal_thickavg | -0.011 | 0.026 | -0.063 | 0.041 | 0.673 | 0.801 | 2611 |
| R_supramarginal_thickavg | -0.024 | 0.027 | -0.076 | 0.028 | 0.374 | 0.534 | 2601 |
| R_frontalpole_thickavg | 0.025 | 0.019 | -0.012 | 0.063 | 0.190 | 0.401 | 2716 |
| R_temporalpole_thickavg | -0.002 | 0.020 | -0.041 | 0.038 | 0.933 | 0.992 | 2612 |
| R_transversetemporal_thickavg | -0.022 | 0.019 | -0.060 | 0.015 | 0.241 | 0.417 | 2729 |
| R_insula_thickavg | 0.003 | 0.036 | -0.067 | 0.073 | 0.928 | 0.992 | 2640 |

### Table S6. Association between CT and negative schizotypy corrected for age, and sex

| DKT Regions | Partial R | Std. Err. | 95% CI, lower bound | 95% CI, upper bound | p-value | FDR-pvalue | Sample size per region |
| --- | --- | --- | --- | --- | --- | --- | --- |
| L_bankssts_thickavg | 0.007 | 0.020 | -0.032 | 0.046 | 0.709 | 0.973 | 2512 |
| L_caudalanteriorcingulate_thickavg | 0.013 | 0.019 | -0.025 | 0.050 | 0.514 | 0.971 | 2668 |
| L_caudalmiddlefrontal_thickavg | 0.031 | 0.029 | -0.025 | 0.087 | 0.282 | 0.784 | 2675 |
| L_cuneus_thickavg | 0.019 | 0.020 | -0.020 | 0.057 | 0.345 | 0.802 | 2538 |
| L_entorhinal_thickavg | 0.002 | 0.020 | -0.037 | 0.041 | 0.905 | 0.979 | 2510 |
| L_fusiform_thickavg | -0.010 | 0.019 | -0.048 | 0.028 | 0.595 | 0.971 | 2640 |
| L_inferiorparietal_thickavg | -0.006 | 0.020 | -0.044 | 0.033 | 0.776 | 0.973 | 2586 |
| L_inferiortemporal_thickavg | 0.004 | 0.020 | -0.035 | 0.042 | 0.855 | 0.979 | 2611 |
| L_isthmuscingulate_thickavg | -0.013 | 0.033 | -0.078 | 0.051 | 0.682 | 0.973 | 2686 |
| L_lateraloccipital_thickavg | -0.006 | 0.024 | -0.053 | 0.040 | 0.786 | 0.973 | 2644 |
| L_lateralorbitofrontal_thickavg | 0.018 | 0.028 | -0.038 | 0.073 | 0.526 | 0.971 | 2659 |
| L_lingual_thickavg | -0.007 | 0.019 | -0.045 | 0.031 | 0.704 | 0.973 | 2622 |
| L_medialorbitofrontal_thickavg | 0.049 | 0.021 | 0.008 | 0.090 | 0.020 | 0.247 | 2642 |
| L_middletemporal_thickavg | 0.001 | 0.020 | -0.038 | 0.041 | 0.940 | 0.979 | 2506 |
| L_parahippocampal_thickavg | 0.030 | 0.019 | -0.007 | 0.068 | 0.113 | 0.560 | 2671 |
| L_paracentral_thickavg | -0.017 | 0.019 | -0.054 | 0.021 | 0.387 | 0.861 | 2682 |
| L_parsopercularis_thickavg | 0.000 | 0.019 | -0.038 | 0.038 | 0.993 | 0.994 | 2658 |
| L_parsorbitalis_thickavg | 0.019 | 0.019 | -0.019 | 0.057 | 0.335 | 0.797 | 2664 |
| L_parstriangularis_thickavg | 0.019 | 0.025 | -0.031 | 0.069 | 0.460 | 0.925 | 2649 |
| L_pericalcarine_thickavg | 0.019 | 0.030 | -0.039 | 0.078 | 0.522 | 0.971 | 2592 |
| L_postcentral_thickavg | 0.009 | 0.025 | -0.040 | 0.057 | 0.729 | 0.973 | 2618 |
| L_posteriorcingulate_thickavg | 0.011 | 0.027 | -0.041 | 0.064 | 0.676 | 0.973 | 2685 |
| L_precentral_thickavg | 0.002 | 0.021 | -0.040 | 0.044 | 0.921 | 0.979 | 2638 |
| L_precuneus_thickavg | 0.002 | 0.021 | -0.040 | 0.044 | 0.935 | 0.979 | 2666 |
| L_rostralanteriorcingulate_thickavg | 0.060 | 0.019 | 0.022 | 0.098 | 0.002 | 0.036 | 2657 |
| L_rostralmiddlefrontal_thickavg | 0.006 | 0.031 | -0.055 | 0.067 | 0.855 | 0.979 | 2665 |
| L_superiorfrontal_thickavg | 0.029 | 0.025 | -0.019 | 0.077 | 0.242 | 0.780 | 2665 |
| L_superiorparietal_thickavg | 0.008 | 0.023 | -0.037 | 0.053 | 0.734 | 0.973 | 2623 |
| L_superiortemporal_thickavg | 0.011 | 0.020 | -0.028 | 0.050 | 0.573 | 0.971 | 2492 |
| L_supramarginal_thickavg | 0.004 | 0.020 | -0.036 | 0.044 | 0.862 | 0.979 | 2530 |
| L_frontalpole_thickavg | 0.049 | 0.019 | 0.012 | 0.087 | 0.010 | 0.168 | 2691 |
| L_temporalpole_thickavg | 0.031 | 0.019 | -0.007 | 0.069 | 0.105 | 0.560 | 2640 |
| L_transversetemporal_thickavg | 0.030 | 0.019 | -0.007 | 0.068 | 0.114 | 0.560 | 2691 |
| L_insula_thickavg | 0.025 | 0.025 | -0.024 | 0.074 | 0.316 | 0.797 | 2618 |
| R_bankssts_thickavg | -0.010 | 0.019 | -0.048 | 0.028 | 0.592 | 0.971 | 2620 |
| R_caudalanteriorcingulate_thickavg | 0.051 | 0.024 | 0.004 | 0.098 | 0.035 | 0.319 | 2683 |
| R_caudalmiddlefrontal_thickavg | 0.053 | 0.030 | -0.007 | 0.112 | 0.082 | 0.491 | 2667 |
| R_cuneus_thickavg | 0.018 | 0.024 | -0.029 | 0.065 | 0.456 | 0.925 | 2569 |
| R_entorhinal_thickavg | 0.029 | 0.023 | -0.017 | 0.075 | 0.211 | 0.763 | 2431 |
| R_fusiform_thickavg | 0.015 | 0.019 | -0.023 | 0.053 | 0.442 | 0.925 | 2647 |
| R_inferiorparietal_thickavg | 0.010 | 0.020 | -0.028 | 0.049 | 0.599 | 0.971 | 2584 |
| R_inferiortemporal_thickavg | 0.007 | 0.028 | -0.048 | 0.063 | 0.797 | 0.973 | 2621 |
| R_isthmuscingulate_thickavg | -0.012 | 0.028 | -0.067 | 0.043 | 0.669 | 0.973 | 2675 |
| R_lateraloccipital_thickavg | 0.000 | 0.019 | -0.038 | 0.038 | 0.994 | 0.994 | 2638 |
| R_lateralorbitofrontal_thickavg | 0.024 | 0.023 | -0.021 | 0.068 | 0.296 | 0.797 | 2636 |
| R_lingual_thickavg | 0.024 | 0.019 | -0.015 | 0.062 | 0.225 | 0.775 | 2623 |
| R_medialorbitofrontal_thickavg | 0.073 | 0.020 | 0.033 | 0.112 | 0.000 | 0.016 | 2622 |
| R_middletemporal_thickavg | 0.017 | 0.023 | -0.028 | 0.062 | 0.454 | 0.925 | 2593 |
| R_parahippocampal_thickavg | 0.008 | 0.019 | -0.030 | 0.046 | 0.686 | 0.973 | 2673 |
| R_paracentral_thickavg | 0.008 | 0.023 | -0.038 | 0.053 | 0.741 | 0.973 | 2679 |
| R_parsopercularis_thickavg | 0.039 | 0.019 | 0.001 | 0.077 | 0.044 | 0.370 | 2637 |
| R_parsorbitalis_thickavg | 0.037 | 0.037 | -0.036 | 0.111 | 0.323 | 0.797 | 2662 |
| R_parstriangularis_thickavg | 0.040 | 0.026 | -0.010 | 0.090 | 0.119 | 0.560 | 2628 |
| R_pericalcarine_thickavg | 0.051 | 0.022 | 0.008 | 0.093 | 0.020 | 0.247 | 2576 |
| R_postcentral_thickavg | -0.010 | 0.019 | -0.048 | 0.028 | 0.612 | 0.971 | 2632 |
| R_posteriorcingulate_thickavg | 0.038 | 0.027 | -0.015 | 0.090 | 0.157 | 0.685 | 2686 |
| R_precentral_thickavg | 0.018 | 0.034 | -0.049 | 0.085 | 0.592 | 0.971 | 2636 |
| R_precuneus_thickavg | -0.001 | 0.027 | -0.055 | 0.052 | 0.957 | 0.987 | 2670 |
| R_rostralanteriorcingulate_thickavg | 0.061 | 0.019 | 0.023 | 0.099 | 0.002 | 0.036 | 2633 |
| R_rostralmiddlefrontal_thickavg | 0.008 | 0.033 | -0.056 | 0.072 | 0.811 | 0.975 | 2643 |
| R_superiorfrontal_thickavg | 0.058 | 0.026 | 0.007 | 0.110 | 0.026 | 0.264 | 2664 |
| R_superiorparietal_thickavg | 0.020 | 0.020 | -0.020 | 0.059 | 0.328 | 0.797 | 2638 |
| R_superiortemporal_thickavg | 0.024 | 0.020 | -0.014 | 0.063 | 0.212 | 0.763 | 2569 |
| R_supramarginal_thickavg | 0.021 | 0.020 | -0.017 | 0.060 | 0.276 | 0.784 | 2559 |
| R_frontalpole_thickavg | 0.050 | 0.026 | -0.002 | 0.102 | 0.059 | 0.453 | 2682 |
| R_temporalpole_thickavg | 0.052 | 0.023 | 0.007 | 0.096 | 0.022 | 0.247 | 2577 |
| R_transversetemporal_thickavg | 0.015 | 0.024 | -0.032 | 0.062 | 0.523 | 0.971 | 2694 |
| R_insula_thickavg | 0.012 | 0.020 | -0.027 | 0.050 | 0.558 | 0.971 | 2585 |

Table S7. Positive schizotypy questionnaire moderator of association between CT and positive schizotypy

| DKT Regions | Q | p-value | FDR p-value |
| --- | --- | --- | --- |
| L_bankssts_thickavg | 14.274 | 0.075 | 0.259 |
| L_caudalanteriorcingulate_thickavg | 13.430 | 0.098 | 0.273 |
| L_caudalmiddlefrontal_thickavg | 17.828 | 0.023 | 0.125 |
| L_cuneus_thickavg | 9.784 | 0.281 | 0.481 |
| L_entorhinal_thickavg | 13.754 | 0.088 | 0.273 |
| L_frontalpole_thickavg | 11.621 | 0.169 | 0.392 |
| L_fusiform_thickavg | 10.545 | 0.229 | 0.438 |
| L_inferiorparietal_thickavg | 17.155 | 0.029 | 0.137 |
| L_inferiortemporal_thickavg | 10.348 | 0.241 | 0.438 |
| L_insula_thickavg | 11.514 | 0.174 | 0.392 |
| L_isthmuscingulate_thickavg | 13.411 | 0.098 | 0.273 |
| L_lateraloccipital_thickavg | 8.232 | 0.411 | 0.617 |
| L_lateralorbitofrontal_thickavg | 3.732 | 0.880 | 0.946 |
| L_lingual_thickavg | 10.051 | 0.261 | 0.459 |
| L_medialorbitofrontal_thickavg | 5.702 | 0.681 | 0.860 |
| L_middletemporal_thickavg | 11.804 | 0.160 | 0.384 |
| L_paracentral_thickavg | 5.004 | 0.757 | 0.879 |
| L_parahippocampal_thickavg | 2.348 | 0.968 | 0.982 |
| L_parsopercularis_thickavg | 15.693 | 0.047 | 0.188 |
| L_parsorbitalis_thickavg | 21.847 | 0.005 | 0.054 |
| L_parstriangularis_thickavg | 5.741 | 0.676 | 0.860 |
| L_pericalcarine_thickavg | 8.263 | 0.408 | 0.617 |
| L_postcentral_thickavg | 7.647 | 0.469 | 0.662 |
| L_posteriorcingulate_thickavg | 3.125 | 0.926 | 0.980 |
| L_precentral_thickavg | 23.180 | 0.003 | 0.040 |
| L_precuneus_thickavg | 31.255 | 0.000 | 0.005 |
| L_rostralanteriorcingulate_thickavg | 2.489 | 0.962 | 0.982 |
| L_rostralmiddlefrontal_thickavg | 14.245 | 0.076 | 0.259 |
| L_superiorfrontal_thickavg | 8.641 | 0.373 | 0.585 |
| L_superiorparietal_thickavg | 13.252 | 0.103 | 0.276 |
| L_superiortemporal_thickavg | 18.336 | 0.019 | 0.125 |
| L_supramarginal_thickavg | 10.316 | 0.244 | 0.438 |
| L_temporalpole_thickavg | 5.020 | 0.755 | 0.879 |
| L_transversetemporal_thickavg | 10.501 | 0.232 | 0.438 |
| R_bankssts_thickavg | 23.836 | 0.002 | 0.040 |
| R_caudalanteriorcingulate_thickavg | 18.075 | 0.021 | 0.125 |
| R_caudalmiddlefrontal_thickavg | 11.180 | 0.192 | 0.406 |
| R_cuneus_thickavg | 23.021 | 0.003 | 0.040 |
| R_entorhinal_thickavg | 1.727 | 0.988 | 0.988 |
| R_frontalpole_thickavg | 8.109 | 0.423 | 0.621 |
| R_fusiform_thickavg | 15.310 | 0.053 | 0.202 |
| R_inferiorparietal_thickavg | 15.995 | 0.042 | 0.188 |
| R_inferiortemporal_thickavg | 4.836 | 0.775 | 0.886 |
| R_insula_thickavg | 8.022 | 0.431 | 0.621 |
| R_isthmuscingulate_thickavg | 7.150 | 0.521 | 0.707 |
| R_lateraloccipital_thickavg | 4.379 | 0.821 | 0.924 |
| R_lateralorbitofrontal_thickavg | 5.085 | 0.748 | 0.879 |
| R_lingual_thickavg | 5.913 | 0.657 | 0.860 |
| R_medialorbitofrontal_thickavg | 10.658 | 0.222 | 0.438 |
| R_middletemporal_thickavg | 15.739 | 0.046 | 0.188 |
| R_paracentral_thickavg | 17.958 | 0.022 | 0.125 |
| R_parahippocampal_thickavg | 2.921 | 0.939 | 0.980 |
| R_parsopercularis_thickavg | 13.451 | 0.097 | 0.273 |
| R_parsorbitalis_thickavg | 6.323 | 0.611 | 0.815 |
| R_parstriangularis_thickavg | 4.241 | 0.835 | 0.925 |
| R_pericalcarine_thickavg | 10.713 | 0.218 | 0.438 |
| R_postcentral_thickavg | 5.304 | 0.725 | 0.879 |
| R_posteriorcingulate_thickavg | 7.336 | 0.501 | 0.693 |
| R_precentral_thickavg | 21.009 | 0.007 | 0.064 |
| R_precuneus_thickavg | 12.548 | 0.128 | 0.330 |
| R_rostralanteriorcingulate_thickavg | 5.348 | 0.720 | 0.879 |
| R_rostralmiddlefrontal_thickavg | 9.030 | 0.340 | 0.546 |
| R_superiorfrontal_thickavg | 27.783 | 0.001 | 0.012 |
| R_superiorparietal_thickavg | 20.318 | 0.009 | 0.074 |
| R_superiortemporal_thickavg | 32.057 | 0.000 | 0.005 |
| R_supramarginal_thickavg | 9.570 | 0.296 | 0.496 |
| R_temporalpole_thickavg | 9.012 | 0.341 | 0.546 |
| R_transversetemporal_thickavg | 3.763 | 0.878 | 0.946 |

### Table S8. Freesurfer version moderator of association with CT and positive schizotypy

| DKT regions | Q | p-value | FDR p-value |
| --- | --- | --- | --- |
| L_bankssts_thickavg | 0.902 | 0.970 | 0.953 |
| L_caudalanteriorcingulate_thickavg | 0.233 | 0.999 | 0.995 |
| L_caudalmiddlefrontal_thickavg | 2.346 | 0.800 | 0.953 |
| L_cuneus_thickavg | 4.581 | 0.469 | 0.953 |
| L_entorhinal_thickavg | 1.007 | 0.962 | 0.953 |
| L_fusiform_thickavg | 3.572 | 0.613 | 0.995 |
| L_inferiorparietal_thickavg | 3.096 | 0.685 | 0.953 |
| L_inferiortemporal_thickavg | 2.554 | 0.768 | 0.953 |
| L_isthmuscingulate_thickavg | 4.024 | 0.546 | 0.953 |
| L_lateraloccipital_thickavg | 2.991 | 0.701 | 0.953 |
| L_lateralorbitofrontal_thickavg | 4.232 | 0.516 | 0.953 |
| L_lingual_thickavg | 1.975 | 0.853 | 0.953 |
| L_medialorbitofrontal_thickavg | 9.625 | 0.087 | 0.953 |
| L_middletemporal_thickavg | 0.875 | 0.972 | 0.953 |
| L_parahippocampal_thickavg | 4.837 | 0.436 | 0.953 |
| L_paracentral_thickavg | 1.975 | 0.853 | 0.953 |
| L_parsopercularis_thickavg | 1.350 | 0.930 | 0.953 |
| L_parsorbitalis_thickavg | 8.423 | 0.134 | 0.953 |
| L_parstriangularis_thickavg | 1.307 | 0.934 | 0.953 |
| L_pericalcarine_thickavg | 4.119 | 0.532 | 0.953 |
| L_postcentral_thickavg | 8.864 | 0.115 | 0.953 |
| L_posteriorcingulate_thickavg | 3.713 | 0.591 | 0.953 |
| L_precentral_thickavg | 8.168 | 0.147 | 0.953 |
| L_precuneus_thickavg | 0.900 | 0.970 | 0.998 |
| L_rostralanteriorcingulate_thickavg | 4.092 | 0.536 | 0.953 |
| L_rostralmiddlefrontal_thickavg | 7.183 | 0.207 | 0.953 |
| L_superiorfrontal_thickavg | 2.640 | 0.755 | 0.953 |
| L_superiorparietal_thickavg | 2.814 | 0.729 | 0.953 |
| L_superiortemporal_thickavg | 3.170 | 0.674 | 0.953 |
| L_supramarginal_thickavg | 4.129 | 0.531 | 0.995 |
| L_frontalpole_thickavg | 3.285 | 0.656 | 0.995 |
| L_temporalpole_thickavg | 2.911 | 0.714 | 0.953 |
| L_transversetemporal_thickavg | 5.417 | 0.367 | 0.953 |
| L_insula_thickavg | 5.981 | 0.308 | 0.953 |
| R_bankssts_thickavg | 5.013 | 0.414 | 0.953 |
| R_caudalanteriorcingulate_thickavg | 5.375 | 0.372 | 0.953 |
| R_caudalmiddlefrontal_thickavg | 8.295 | 0.141 | 0.953 |
| R_cuneus_thickavg | 4.351 | 0.500 | 0.953 |
| R_entorhinal_thickavg | 2.333 | 0.801 | 0.953 |
| R_fusiform_thickavg | 3.421 | 0.635 | 0.953 |
| R_inferiorparietal_thickavg | 2.721 | 0.743 | 0.953 |
| R_inferiortemporal_thickavg | 1.827 | 0.872 | 0.953 |
| R_isthmuscingulate_thickavg | 7.385 | 0.194 | 0.953 |
| R_lateraloccipital_thickavg | 5.431 | 0.366 | 0.953 |
| R_lateralorbitofrontal_thickavg | 4.561 | 0.472 | 0.953 |
| R_lingual_thickavg | 6.731 | 0.241 | 0.953 |
| R_medialorbitofrontal_thickavg | 14.690 | 0.012 | 0.953 |
| R_middletemporal_thickavg | 1.121 | 0.952 | 0.953 |
| R_parahippocampal_thickavg | 5.833 | 0.323 | 0.953 |
| R_paracentral_thickavg | 0.558 | 0.990 | 0.995 |
| R_parsopercularis_thickavg | 10.106 | 0.072 | 0.953 |
| R_parsorbitalis_thickavg | 4.084 | 0.537 | 0.953 |
| R_parstriangularis_thickavg | 7.357 | 0.195 | 0.953 |
| R_pericalcarine_thickavg | 9.595 | 0.088 | 0.953 |
| R_postcentral_thickavg | 6.304 | 0.278 | 0.953 |
| R_posteriorcingulate_thickavg | 2.524 | 0.773 | 0.953 |
| R_precentral_thickavg | 4.723 | 0.451 | 0.953 |
| R_precuneus_thickavg | 2.063 | 0.840 | 0.953 |
| R_rostralanteriorcingulate_thickavg | 5.578 | 0.349 | 0.953 |
| R_rostralmiddlefrontal_thickavg | 1.912 | 0.861 | 0.953 |
| R_superiorfrontal_thickavg | 7.094 | 0.214 | 0.953 |
| R_superiorparietal_thickavg | 3.296 | 0.654 | 0.953 |
| R_superiortemporal_thickavg | 3.755 | 0.585 | 0.953 |
| R_supramarginal_thickavg | 3.381 | 0.641 | 0.953 |
| R_frontalpole_thickavg | 3.846 | 0.572 | 0.953 |
| R_temporalpole_thickavg | 5.780 | 0.328 | 0.998 |
| R_transversetemporal_thickavg | 5.900 | 0.316 | 0.953 |
| R_insula_thickavg | 0.600 | 0.988 | 0.953 |

### Table S9. Field strength moderator of association with CT and positive schizotypy

| DKT regions | Q | p-value | FDR-adjusted p-value |
| --- | --- | --- | --- |
| L_bankssts_thickavg | 0.791 | 0.673 | 0.953 |
| L_caudalanteriorcingulate_thickavg | 0.115 | 0.944 | 0.995 |
| L_caudalmiddlefrontal_thickavg | 0.392 | 0.822 | 0.953 |
| L_cuneus_thickavg | 0.425 | 0.809 | 0.953 |
| L_entorhinal_thickavg | 0.911 | 0.634 | 0.953 |
| L_fusiform_thickavg | 0.184 | 0.912 | 0.995 |
| L_inferiorparietal_thickavg | 0.452 | 0.798 | 0.953 |
| L_inferiortemporal_thickavg | 1.426 | 0.490 | 0.953 |
| L_isthmuscingulate_thickavg | 1.734 | 0.420 | 0.953 |
| L_lateraloccipital_thickavg | 0.997 | 0.607 | 0.953 |
| L_lateralorbitofrontal_thickavg | 2.957 | 0.228 | 0.953 |
| L_lingual_thickavg | 0.943 | 0.624 | 0.953 |
| L_medialorbitofrontal_thickavg | 4.074 | 0.130 | 0.953 |
| L_middletemporal_thickavg | 0.337 | 0.845 | 0.953 |
| L_parahippocampal_thickavg | 3.831 | 0.147 | 0.953 |
| L_paracentral_thickavg | 1.638 | 0.441 | 0.953 |
| L_parsopercularis_thickavg | 0.640 | 0.726 | 0.953 |
| L_parsorbitalis_thickavg | 1.184 | 0.553 | 0.953 |
| L_parstriangularis_thickavg | 0.605 | 0.739 | 0.953 |
| L_pericalcarine_thickavg | 0.812 | 0.666 | 0.953 |
| L_postcentral_thickavg | 0.345 | 0.842 | 0.953 |
| L_posteriorcingulate_thickavg | 0.377 | 0.828 | 0.953 |
| L_precentral_thickavg | 1.370 | 0.504 | 0.953 |
| L_precuneus_thickavg | 0.004 | 0.998 | 0.998 |
| L_rostralanteriorcingulate_thickavg | 0.474 | 0.789 | 0.953 |
| L_rostralmiddlefrontal_thickavg | 3.532 | 0.171 | 0.953 |
| L_superiorfrontal_thickavg | 2.313 | 0.315 | 0.953 |
| L_superiorparietal_thickavg | 0.693 | 0.707 | 0.953 |
| L_superiortemporal_thickavg | 0.563 | 0.755 | 0.953 |
| L_supramarginal_thickavg | 0.071 | 0.965 | 0.995 |
| L_frontalpole_thickavg | 0.112 | 0.945 | 0.995 |
| L_temporalpole_thickavg | 0.797 | 0.671 | 0.953 |
| L_transversetemporal_thickavg | 5.257 | 0.072 | 0.953 |
| L_insula_thickavg | 3.155 | 0.206 | 0.953 |
| R_bankssts_thickavg | 4.260 | 0.119 | 0.953 |
| R_caudalanteriorcingulate_thickavg | 2.505 | 0.286 | 0.953 |
| R_caudalmiddlefrontal_thickavg | 3.453 | 0.178 | 0.953 |
| R_cuneus_thickavg | 0.836 | 0.658 | 0.953 |
| R_entorhinal_thickavg | 1.360 | 0.507 | 0.953 |
| R_fusiform_thickavg | 0.570 | 0.752 | 0.953 |
| R_inferiorparietal_thickavg | 2.466 | 0.291 | 0.953 |
| R_inferiortemporal_thickavg | 0.320 | 0.852 | 0.953 |
| R_isthmuscingulate_thickavg | 1.359 | 0.507 | 0.953 |
| R_lateraloccipital_thickavg | 0.305 | 0.859 | 0.953 |
| R_lateralorbitofrontal_thickavg | 4.524 | 0.104 | 0.953 |
| R_lingual_thickavg | 1.179 | 0.555 | 0.953 |
| R_medialorbitofrontal_thickavg | 8.611 | 0.013 | 0.953 |
| R_middletemporal_thickavg | 0.431 | 0.806 | 0.953 |
| R_parahippocampal_thickavg | 0.969 | 0.616 | 0.953 |
| R_paracentral_thickavg | 0.066 | 0.968 | 0.995 |
| R_parsopercularis_thickavg | 1.533 | 0.465 | 0.953 |
| R_parsorbitalis_thickavg | 2.394 | 0.302 | 0.953 |
| R_parstriangularis_thickavg | 2.215 | 0.330 | 0.953 |
| R_pericalcarine_thickavg | 1.530 | 0.465 | 0.953 |
| R_postcentral_thickavg | 1.467 | 0.480 | 0.953 |
| R_posteriorcingulate_thickavg | 0.610 | 0.737 | 0.953 |
| R_precentral_thickavg | 1.270 | 0.530 | 0.953 |
| R_precuneus_thickavg | 0.832 | 0.660 | 0.953 |
| R_rostralanteriorcingulate_thickavg | 1.626 | 0.444 | 0.953 |
| R_rostralmiddlefrontal_thickavg | 1.581 | 0.454 | 0.953 |
| R_superiorfrontal_thickavg | 2.459 | 0.292 | 0.953 |
| R_superiorparietal_thickavg | 1.536 | 0.464 | 0.953 |
| R_superiortemporal_thickavg | 0.664 | 0.718 | 0.953 |
| R_supramarginal_thickavg | 0.691 | 0.708 | 0.953 |
| R_frontalpole_thickavg | 2.521 | 0.283 | 0.953 |
| R_temporalpole_thickavg | 0.009 | 0.996 | 0.998 |
| R_transversetemporal_thickavg | 2.030 | 0.362 | 0.953 |
| R_insula_thickavg | 0.367 | 0.832 | 0.953 |

### Table S10. Negative schizotypy questionnaire moderator of association between CT and negative schizotypy

| DKT regions | Q | p-value | FDR p-value |
| --- | --- | --- | --- |
| L_bankssts_thickavg | 6.612 | 0.251 | 0.887 |
| L_caudalanteriorcingulate_thickavg | 1.509 | 0.912 | 0.999 |
| L_caudalmiddlefrontal_thickavg | 6.398 | 0.269 | 0.887 |
| L_cuneus_thickavg | 7.819 | 0.166 | 0.821 |
| L_entorhinal_thickavg | 4.980 | 0.418 | 0.911 |
| L_frontalpole_thickavg | 2.481 | 0.779 | 0.960 |
| L_fusiform_thickavg | 3.475 | 0.627 | 0.911 |
| L_inferiorparietal_thickavg | 2.398 | 0.792 | 0.960 |
| L_inferiortemporal_thickavg | 4.143 | 0.529 | 0.911 |
| L_insula_thickavg | 6.205 | 0.287 | 0.887 |
| L_isthmuscingulate_thickavg | 0.532 | 0.991 | 0.999 |
| L_lateraloccipital_thickavg | 2.976 | 0.704 | 0.911 |
| L_lateralorbitofrontal_thickavg | 9.222 | 0.101 | 0.531 |
| L_lingual_thickavg | 1.200 | 0.945 | 0.999 |
| L_medialorbitofrontal_thickavg | 2.910 | 0.714 | 0.911 |
| L_middletemporal_thickavg | 1.467 | 0.917 | 0.999 |
| L_paracentral_thickavg | 3.495 | 0.624 | 0.911 |
| L_parahippocampal_thickavg | 3.096 | 0.685 | 0.911 |
| L_parsopercularis_thickavg | 13.511 | 0.019 | 0.282 |
| L_parsorbitalis_thickavg | 4.165 | 0.526 | 0.911 |
| L_parstriangularis_thickavg | 1.457 | 0.918 | 0.999 |
| L_pericalcarine_thickavg | 1.055 | 0.958 | 0.999 |
| L_postcentral_thickavg | 6.006 | 0.306 | 0.887 |
| L_posteriorcingulate_thickavg | 3.423 | 0.635 | 0.911 |
| L_precentral_thickavg | 5.380 | 0.371 | 0.887 |
| L_precuneus_thickavg | 3.672 | 0.597 | 0.911 |
| L_rostralanteriorcingulate_thickavg | 4.517 | 0.478 | 0.911 |
| L_rostralmiddlefrontal_thickavg | 5.189 | 0.393 | 0.909 |
| L_superiorfrontal_thickavg | 5.542 | 0.353 | 0.887 |
| L_superiorparietal_thickavg | 1.755 | 0.882 | 0.999 |
| L_superiortemporal_thickavg | 5.570 | 0.350 | 0.887 |
| L_supramarginal_thickavg | 5.920 | 0.314 | 0.887 |
| L_temporalpole_thickavg | 4.253 | 0.514 | 0.911 |
| L_transversetemporal_thickavg | 2.506 | 0.776 | 0.960 |
| R_bankssts_thickavg | 11.634 | 0.040 | 0.495 |
| R_caudalanteriorcingulate_thickavg | 5.378 | 0.371 | 0.887 |
| R_caudalmiddlefrontal_thickavg | 23.504 | 0.000 | 0.020 |
| R_cuneus_thickavg | 0.993 | 0.963 | 0.999 |
| R_entorhinal_thickavg | 3.527 | 0.619 | 0.911 |
| R_frontalpole_thickavg | 6.075 | 0.299 | 0.887 |
| R_fusiform_thickavg | 3.224 | 0.665 | 0.911 |
| R_inferiorparietal_thickavg | 3.794 | 0.579 | 0.911 |
| R_inferiortemporal_thickavg | 9.993 | 0.075 | 0.529 |
| R_insula_thickavg | 7.015 | 0.220 | 0.887 |
| R_isthmuscingulate_thickavg | 1.847 | 0.870 | 0.999 |
| R_lateraloccipital_thickavg | 2.987 | 0.702 | 0.911 |
| R_lateralorbitofrontal_thickavg | 4.251 | 0.514 | 0.911 |
| R_lingual_thickavg | 3.273 | 0.658 | 0.911 |
| R_medialorbitofrontal_thickavg | 5.598 | 0.347 | 0.887 |
| R_middletemporal_thickavg | 6.221 | 0.285 | 0.887 |
| R_paracentral_thickavg | 18.231 | 0.003 | 0.099 |
| R_parahippocampal_thickavg | 6.295 | 0.279 | 0.887 |
| R_parsopercularis_thickavg | 9.438 | 0.093 | 0.529 |
| R_parsorbitalis_thickavg | 9.503 | 0.091 | 0.529 |
| R_parstriangularis_thickavg | 13.697 | 0.018 | 0.282 |
| R_pericalcarine_thickavg | 4.029 | 0.545 | 0.911 |
| R_postcentral_thickavg | 3.464 | 0.629 | 0.911 |
| R_posteriorcingulate_thickavg | 3.699 | 0.593 | 0.911 |
| R_precentral_thickavg | 3.274 | 0.658 | 0.911 |
| R_precuneus_thickavg | 2.163 | 0.826 | 0.986 |
| R_rostralanteriorcingulate_thickavg | 5.462 | 0.362 | 0.887 |
| R_rostralmiddlefrontal_thickavg | 10.238 | 0.069 | 0.529 |
| R_superiorfrontal_thickavg | 14.554 | 0.012 | 0.282 |
| R_superiorparietal_thickavg | 4.806 | 0.440 | 0.911 |
| R_superiortemporal_thickavg | 10.210 | 0.069 | 0.529 |
| R_supramarginal_thickavg | 3.928 | 0.560 | 0.911 |
| R_temporalpole_thickavg | 3.894 | 0.565 | 0.911 |
| R_transversetemporal_thickavg | 0.792 | 0.978 | 0.999 |

### Table S11. Freesurfer version moderator of association with CT and negative schizotypy

| DTK regions | Q | p-value | FDR p-value |
| --- | --- | --- | --- |
| L_bankssts_thickavg | 5.525 | 0.355 | 0.822 |
| L_caudalanteriorcingulate_thickavg | 3.440 | 0.632 | 0.947 |
| L_caudalmiddlefrontal_thickavg | 2.654 | 0.753 | 0.947 |
| L_cuneus_thickavg | 9.123 | 0.104 | 0.563 |
| L_entorhinal_thickavg | 4.287 | 0.509 | 0.897 |
| L_frontalpole_thickavg | 8.586 | 0.127 | 0.563 |
| L_fusiform_thickavg | 6.372 | 0.272 | 0.649 |
| L_inferiorparietal_thickavg | 2.674 | 0.750 | 0.947 |
| L_inferiortemporal_thickavg | 8.804 | 0.117 | 0.563 |
| L_insula_thickavg | 11.641 | 0.040 | 0.563 |
| L_isthmuscingulate_thickavg | 0.220 | 0.999 | 0.999 |
| L_lateraloccipital_thickavg | 3.066 | 0.690 | 0.947 |
| L_lateralorbitofrontal_thickavg | 21.071 | 0.001 | 0.058 |
| L_lingual_thickavg | 3.380 | 0.642 | 0.947 |
| L_medialorbitofrontal_thickavg | 4.928 | 0.425 | 0.873 |
| L_middletemporal_thickavg | 6.778 | 0.238 | 0.648 |
| L_paracentral_thickavg | 2.533 | 0.772 | 0.947 |
| L_parahippocampal_thickavg | 9.754 | 0.083 | 0.563 |
| L_parsopercularis_thickavg | 9.926 | 0.077 | 0.563 |
| L_parsorbitalis_thickavg | 2.967 | 0.705 | 0.947 |
| L_parstriangularis_thickavg | 3.945 | 0.557 | 0.937 |
| L_pericalcarine_thickavg | 3.158 | 0.676 | 0.947 |
| L_postcentral_thickavg | 8.676 | 0.123 | 0.563 |
| L_posteriorcingulate_thickavg | 8.043 | 0.154 | 0.592 |
| L_precentral_thickavg | 2.050 | 0.842 | 0.999 |
| L_precuneus_thickavg | 3.208 | 0.668 | 0.947 |
| L_rostralanteriorcingulate_thickavg | 7.590 | 0.180 | 0.599 |
| L_rostralmiddlefrontal_thickavg | 4.335 | 0.502 | 0.897 |
| L_superiorfrontal_thickavg | 2.834 | 0.726 | 0.947 |
| L_superiorparietal_thickavg | 2.876 | 0.719 | 0.947 |
| L_superiortemporal_thickavg | 4.432 | 0.489 | 0.897 |
| L_supramarginal_thickavg | 7.589 | 0.180 | 0.599 |
| L_temporalpole_thickavg | 6.391 | 0.270 | 0.649 |
| L_transversetemporal_thickavg | 2.725 | 0.742 | 0.947 |
| R_bankssts_thickavg | 10.139 | 0.071 | 0.563 |
| R_caudalanteriorcingulate_thickavg | 4.738 | 0.449 | 0.876 |
| R_caudalmiddlefrontal_thickavg | 7.497 | 0.186 | 0.599 |
| R_cuneus_thickavg | 8.528 | 0.129 | 0.563 |
| R_entorhinal_thickavg | 3.073 | 0.689 | 0.947 |
| R_frontalpole_thickavg | 1.542 | 0.908 | 0.999 |
| R_fusiform_thickavg | 2.472 | 0.781 | 0.947 |
| R_inferiorparietal_thickavg | 1.521 | 0.911 | 0.999 |
| R_inferiortemporal_thickavg | 7.333 | 0.197 | 0.608 |
| R_insula_thickavg | 8.553 | 0.128 | 0.563 |
| R_isthmuscingulate_thickavg | 1.862 | 0.868 | 0.999 |
| R_lateraloccipital_thickavg | 4.729 | 0.450 | 0.876 |
| R_lateralorbitofrontal_thickavg | 7.149 | 0.210 | 0.621 |
| R_lingual_thickavg | 6.736 | 0.241 | 0.648 |
| R_medialorbitofrontal_thickavg | 10.734 | 0.057 | 0.563 |
| R_middletemporal_thickavg | 8.729 | 0.120 | 0.563 |
| R_paracentral_thickavg | 6.654 | 0.248 | 0.648 |
| R_parahippocampal_thickavg | 17.773 | 0.003 | 0.120 |
| R_parsopercularis_thickavg | 8.019 | 0.155 | 0.592 |
| R_parsorbitalis_thickavg | 2.638 | 0.756 | 0.947 |
| R_parstriangularis_thickavg | 14.229 | 0.014 | 0.351 |
| R_pericalcarine_thickavg | 0.993 | 0.963 | 0.999 |
| R_postcentral_thickavg | 4.974 | 0.419 | 0.873 |
| R_posteriorcingulate_thickavg | 1.118 | 0.953 | 0.999 |
| R_precentral_thickavg | 1.869 | 0.867 | 0.999 |
| R_precuneus_thickavg | 1.433 | 0.921 | 0.999 |
| R_rostralanteriorcingulate_thickavg | 0.517 | 0.992 | 0.999 |
| R_rostralmiddlefrontal_thickavg | 4.579 | 0.469 | 0.891 |
| R_superiorfrontal_thickavg | 3.263 | 0.659 | 0.947 |
| R_superiorparietal_thickavg | 0.906 | 0.970 | 0.999 |
| R_superiortemporal_thickavg | 6.579 | 0.254 | 0.648 |
| R_supramarginal_thickavg | 7.932 | 0.160 | 0.592 |
| R_temporalpole_thickavg | 10.450 | 0.063 | 0.563 |
| R_transversetemporal_thickavg | 8.578 | 0.127 | 0.563 |

### Table S12. Field strength moderator of association with CT and negative schizotypy

| DKT regions | Q | p-value | FDR p-value |
| --- | --- | --- | --- |
| L_bankssts_thickavg | 0.802 | 0.370 | 0.977 |
| L_caudalanteriorcingulate_thickavg | 0.439 | 0.508 | 0.977 |
| L_caudalmiddlefrontal_thickavg | 0.009 | 0.927 | 0.977 |
| L_cuneus_thickavg | 2.717 | 0.099 | 0.977 |
| L_entorhinal_thickavg | 0.163 | 0.686 | 0.977 |
| L_frontalpole_thickavg | 2.652 | 0.103 | 0.977 |
| L_fusiform_thickavg | 0.036 | 0.849 | 0.977 |
| L_inferiorparietal_thickavg | 0.819 | 0.365 | 0.977 |
| L_inferiortemporal_thickavg | 0.606 | 0.436 | 0.977 |
| L_insula_thickavg | 0.783 | 0.376 | 0.977 |
| L_isthmuscingulate_thickavg | 0.004 | 0.947 | 0.977 |
| L_lateraloccipital_thickavg | 0.228 | 0.633 | 0.977 |
| L_lateralorbitofrontal_thickavg | 0.007 | 0.934 | 0.977 |
| L_lingual_thickavg | 0.115 | 0.734 | 0.977 |
| L_medialorbitofrontal_thickavg | 0.124 | 0.725 | 0.977 |
| L_middletemporal_thickavg | 0.950 | 0.330 | 0.977 |
| L_paracentral_thickavg | 0.004 | 0.950 | 0.977 |
| L_parahippocampal_thickavg | 1.740 | 0.187 | 0.977 |
| L_parsopercularis_thickavg | 0.821 | 0.365 | 0.977 |
| L_parsorbitalis_thickavg | 0.795 | 0.373 | 0.977 |
| L_parstriangularis_thickavg | 0.094 | 0.759 | 0.977 |
| L_pericalcarine_thickavg | 0.541 | 0.462 | 0.977 |
| L_postcentral_thickavg | 0.565 | 0.452 | 0.977 |
| L_posteriorcingulate_thickavg | 0.031 | 0.860 | 0.977 |
| L_precentral_thickavg | 0.246 | 0.620 | 0.977 |
| L_precuneus_thickavg | 0.690 | 0.406 | 0.977 |
| L_rostralanteriorcingulate_thickavg | 1.229 | 0.268 | 0.977 |
| L_rostralmiddlefrontal_thickavg | 0.075 | 0.784 | 0.977 |
| L_superiorfrontal_thickavg | 0.028 | 0.867 | 0.977 |
| L_superiorparietal_thickavg | 0.005 | 0.944 | 0.977 |
| L_superiortemporal_thickavg | 0.337 | 0.562 | 0.977 |
| L_supramarginal_thickavg | 0.185 | 0.667 | 0.977 |
| L_temporalpole_thickavg | 0.297 | 0.586 | 0.977 |
| L_transversetemporal_thickavg | 0.307 | 0.579 | 0.977 |
| R_bankssts_thickavg | 3.749 | 0.053 | 0.977 |
| R_caudalanteriorcingulate_thickavg | 0.019 | 0.890 | 0.977 |
| R_caudalmiddlefrontal_thickavg | 0.033 | 0.857 | 0.977 |
| R_cuneus_thickavg | 0.273 | 0.601 | 0.977 |
| R_entorhinal_thickavg | 0.581 | 0.446 | 0.977 |
| R_frontalpole_thickavg | 0.279 | 0.597 | 0.977 |
| R_fusiform_thickavg | 0.031 | 0.860 | 0.977 |
| R_inferiorparietal_thickavg | 0.205 | 0.651 | 0.977 |
| R_inferiortemporal_thickavg | 1.763 | 0.184 | 0.977 |
| R_insula_thickavg | 1.756 | 0.185 | 0.977 |
| R_isthmuscingulate_thickavg | 0.210 | 0.647 | 0.977 |
| R_lateraloccipital_thickavg | 1.042 | 0.307 | 0.977 |
| R_lateralorbitofrontal_thickavg | 0.464 | 0.496 | 0.977 |
| R_lingual_thickavg | 0.152 | 0.696 | 0.977 |
| R_medialorbitofrontal_thickavg | 0.021 | 0.884 | 0.977 |
| R_middletemporal_thickavg | 0.128 | 0.720 | 0.977 |
| R_paracentral_thickavg | 0.033 | 0.855 | 0.977 |
| R_parahippocampal_thickavg | 9.874 | 0.002 | 0.124 |
| R_parsopercularis_thickavg | 1.683 | 0.194 | 0.977 |
| R_parsorbitalis_thickavg | 0.049 | 0.825 | 0.977 |
| R_parstriangularis_thickavg | 0.129 | 0.719 | 0.977 |
| R_pericalcarine_thickavg | 0.053 | 0.817 | 0.977 |
| R_postcentral_thickavg | 0.977 | 0.323 | 0.977 |
| R_posteriorcingulate_thickavg | 0.030 | 0.863 | 0.977 |
| R_precentral_thickavg | 0.008 | 0.928 | 0.977 |
| R_precuneus_thickavg | 0.000 | 0.997 | 0.997 |
| R_rostralanteriorcingulate_thickavg | 0.002 | 0.966 | 0.979 |
| R_rostralmiddlefrontal_thickavg | 0.149 | 0.700 | 0.977 |
| R_superiorfrontal_thickavg | 0.120 | 0.729 | 0.977 |
| R_superiorparietal_thickavg | 0.141 | 0.707 | 0.977 |
| R_superiortemporal_thickavg | 0.260 | 0.610 | 0.977 |
| R_supramarginal_thickavg | 0.681 | 0.409 | 0.977 |
| R_temporalpole_thickavg | 1.363 | 0.243 | 0.977 |
| R_transversetemporal_thickavg | 2.072 | 0.150 | 0.977 |

### Table S13. Association between SA and positive schizotypy corrected for age, and sex

| DKT Regions | Partial R | Std. Err. | 95% CI, lower bound | 95% CI, upper bound | p-value | FDR p-value | Sample size per region |
| --- | --- | --- | --- | --- | --- | --- | --- |
| L_bankssts_surfavg | 0.015 | 0.020 | -0.025 | 0.055 | 0.465 | 0.934 | 2394 |
| L_caudalanteriorcingulate_surfavg | -0.011 | 0.033 | -0.075 | 0.053 | 0.731 | 0.938 | 2554 |
| L_caudalmiddlefrontal_surfavg | -0.053 | 0.022 | -0.097 | -0.009 | 0.017 | 0.394 | 2559 |
| L_cuneus_surfavg | -0.041 | 0.029 | -0.097 | 0.016 | 0.157 | 0.836 | 2420 |
| L_entorhinal_surfavg | -0.036 | 0.020 | -0.075 | 0.004 | 0.081 | 0.622 | 2395 |
| L_fusiform_surfavg | -0.002 | 0.030 | -0.060 | 0.056 | 0.953 | 0.973 | 2526 |
| L_inferiorparietal_surfavg | 0.014 | 0.034 | -0.052 | 0.080 | 0.684 | 0.934 | 2470 |
| L_inferiortemporal_surfavg | 0.014 | 0.022 | -0.028 | 0.057 | 0.509 | 0.934 | 2496 |
| L_isthmuscingulate_surfavg | 0.007 | 0.027 | -0.045 | 0.059 | 0.778 | 0.938 | 2571 |
| L_lateraloccipital_surfavg | -0.029 | 0.031 | -0.090 | 0.032 | 0.356 | 0.934 | 2529 |
| L_lateralorbitofrontal_surfavg | -0.016 | 0.025 | -0.065 | 0.033 | 0.516 | 0.934 | 2544 |
| L_lingual_surfavg | 0.004 | 0.027 | -0.049 | 0.056 | 0.891 | 0.961 | 2506 |
| L_medialorbitofrontal_surfavg | -0.049 | 0.022 | -0.091 | -0.007 | 0.024 | 0.394 | 2527 |
| L_middletemporal_surfavg | 0.033 | 0.027 | -0.021 | 0.086 | 0.234 | 0.836 | 2388 |
| L_parahippocampal_surfavg | -0.043 | 0.020 | -0.082 | -0.005 | 0.027 | 0.394 | 2555 |
| L_paracentral_surfavg | -0.008 | 0.037 | -0.081 | 0.064 | 0.822 | 0.945 | 2567 |
| L_parsopercularis_surfavg | 0.026 | 0.022 | -0.016 | 0.069 | 0.224 | 0.836 | 2544 |
| L_parsorbitalis_surfavg | 0.003 | 0.030 | -0.056 | 0.062 | 0.930 | 0.961 | 2549 |
| L_parstriangularis_surfavg | 0.003 | 0.020 | -0.035 | 0.042 | 0.874 | 0.961 | 2535 |
| L_pericalcarine_surfavg | -0.021 | 0.024 | -0.068 | 0.025 | 0.368 | 0.934 | 2473 |
| L_postcentral_surfavg | -0.011 | 0.024 | -0.058 | 0.036 | 0.653 | 0.934 | 2505 |
| L_posteriorcingulate_surfavg | 0.017 | 0.021 | -0.024 | 0.058 | 0.411 | 0.934 | 2568 |
| L_precentral_surfavg | -0.031 | 0.023 | -0.077 | 0.015 | 0.187 | 0.836 | 2523 |
| L_precuneus_surfavg | -0.019 | 0.035 | -0.087 | 0.050 | 0.597 | 0.934 | 2558 |
| L_rostralanteriorcingulate_surfavg | -0.043 | 0.023 | -0.089 | 0.002 | 0.063 | 0.622 | 2542 |
| L_rostralmiddlefrontal_surfavg | -0.025 | 0.030 | -0.085 | 0.034 | 0.409 | 0.934 | 2550 |
| L_superiorfrontal_surfavg | -0.023 | 0.026 | -0.073 | 0.027 | 0.371 | 0.934 | 2550 |
| L_superiorparietal_surfavg | 0.013 | 0.030 | -0.046 | 0.071 | 0.668 | 0.934 | 2512 |
| L_superiortemporal_surfavg | -0.013 | 0.025 | -0.062 | 0.036 | 0.613 | 0.934 | 2378 |
| L_supramarginal_surfavg | 0.008 | 0.028 | -0.046 | 0.062 | 0.775 | 0.938 | 2418 |
| L_frontalpole_surfavg | -0.015 | 0.020 | -0.054 | 0.023 | 0.434 | 0.934 | 2576 |
| L_temporalpole_surfavg | -0.030 | 0.029 | -0.087 | 0.027 | 0.301 | 0.900 | 2522 |
| L_transversetemporal_surfavg | -0.023 | 0.022 | -0.065 | 0.020 | 0.292 | 0.900 | 2576 |
| L_insula_surfavg | -0.013 | 0.032 | -0.075 | 0.050 | 0.686 | 0.934 | 2498 |
| R_bankssts_surfavg | 0.017 | 0.026 | -0.034 | 0.067 | 0.521 | 0.934 | 2505 |
| R_caudalanteriorcingulate_surfavg | 0.000 | 0.022 | -0.044 | 0.044 | 0.996 | 0.996 | 2566 |
| R_caudalmiddlefrontal_surfavg | -0.026 | 0.020 | -0.066 | 0.013 | 0.191 | 0.836 | 2550 |
| R_cuneus_surfavg | -0.053 | 0.030 | -0.112 | 0.006 | 0.076 | 0.622 | 2449 |
| R_entorhinal_surfavg | -0.056 | 0.026 | -0.107 | -0.005 | 0.032 | 0.401 | 2309 |
| R_fusiform_surfavg | 0.024 | 0.021 | -0.018 | 0.066 | 0.265 | 0.856 | 2532 |
| R_inferiorparietal_surfavg | 0.022 | 0.031 | -0.038 | 0.083 | 0.468 | 0.934 | 2468 |
| R_inferiortemporal_surfavg | -0.003 | 0.023 | -0.048 | 0.043 | 0.903 | 0.961 | 2506 |
| R_isthmuscingulate_surfavg | 0.008 | 0.030 | -0.052 | 0.068 | 0.790 | 0.940 | 2560 |
| R_lateraloccipital_surfavg | -0.001 | 0.022 | -0.044 | 0.042 | 0.965 | 0.975 | 2522 |
| R_lateralorbitofrontal_surfavg | -0.025 | 0.020 | -0.064 | 0.014 | 0.209 | 0.836 | 2520 |
| R_lingual_surfavg | -0.011 | 0.020 | -0.050 | 0.028 | 0.565 | 0.934 | 2508 |
| R_medialorbitofrontal_surfavg | -0.033 | 0.020 | -0.072 | 0.006 | 0.099 | 0.663 | 2502 |
| R_middletemporal_surfavg | 0.005 | 0.027 | -0.048 | 0.058 | 0.851 | 0.956 | 2480 |
| R_parahippocampal_surfavg | 0.007 | 0.020 | -0.032 | 0.045 | 0.739 | 0.938 | 2558 |
| R_paracentral_surfavg | -0.026 | 0.033 | -0.092 | 0.039 | 0.429 | 0.934 | 2564 |
| R_parsopercularis_surfavg | 0.007 | 0.022 | -0.036 | 0.051 | 0.745 | 0.938 | 2519 |
| R_parsorbitalis_surfavg | -0.025 | 0.020 | -0.064 | 0.013 | 0.196 | 0.836 | 2547 |
| R_parstriangularis_surfavg | 0.010 | 0.024 | -0.036 | 0.056 | 0.666 | 0.934 | 2513 |
| R_pericalcarine_surfavg | -0.031 | 0.020 | -0.070 | 0.009 | 0.126 | 0.787 | 2459 |
| R_postcentral_surfavg | -0.011 | 0.028 | -0.065 | 0.043 | 0.701 | 0.934 | 2515 |
| R_posteriorcingulate_surfavg | -0.020 | 0.029 | -0.077 | 0.036 | 0.480 | 0.934 | 2569 |
| R_precentral_surfavg | -0.014 | 0.020 | -0.053 | 0.025 | 0.495 | 0.934 | 2517 |
| R_precuneus_surfavg | 0.003 | 0.031 | -0.057 | 0.064 | 0.914 | 0.961 | 2558 |
| R_rostralanteriorcingulate_surfavg | 0.019 | 0.025 | -0.030 | 0.068 | 0.456 | 0.934 | 2511 |
| R_rostralmiddlefrontal_surfavg | -0.003 | 0.028 | -0.059 | 0.052 | 0.907 | 0.961 | 2528 |
| R_superiorfrontal_surfavg | -0.034 | 0.029 | -0.091 | 0.022 | 0.234 | 0.836 | 2549 |
| R_superiorparietal_surfavg | 0.015 | 0.020 | -0.025 | 0.055 | 0.471 | 0.934 | 2523 |
| R_superiortemporal_surfavg | -0.011 | 0.027 | -0.064 | 0.042 | 0.685 | 0.934 | 2454 |
| R_supramarginal_surfavg | 0.008 | 0.033 | -0.057 | 0.072 | 0.817 | 0.945 | 2441 |
| R_frontalpole_surfavg | 0.009 | 0.023 | -0.035 | 0.053 | 0.691 | 0.934 | 2566 |
| R_temporalpole_surfavg | -0.034 | 0.020 | -0.073 | 0.005 | 0.089 | 0.636 | 2462 |
| R_transversetemporal_surfavg | -0.038 | 0.031 | -0.099 | 0.023 | 0.221 | 0.836 | 2579 |
| R_insula_surfavg | -0.034 | 0.034 | -0.100 | 0.031 | 0.306 | 0.900 | 2458 |

### Table S14. Association between SA and negative schizotypy corrected for age, and sex

| DKT Regions | Partial R | Std. Err. | 95% CI, lower bound | 95% CI, upper bound | p-value | FDR p-value | Sample size per region |
| --- | --- | --- | --- | --- | --- | --- | --- |
| L_bankssts_surfavg | 0.009 | 0.021 | -0.032 | 0.049 | 0.676 | 0.972 | 2297 |
| L_caudalanteriorcingulate_surfavg | 0.007 | 0.036 | -0.063 | 0.077 | 0.839 | 0.973 | 2446 |
| L_caudalmiddlefrontal_surfavg | -0.008 | 0.020 | -0.048 | 0.031 | 0.687 | 0.972 | 2449 |
| L_cuneus_surfavg | -0.027 | 0.031 | -0.087 | 0.033 | 0.375 | 0.972 | 2322 |
| L_entorhinal_surfavg | -0.026 | 0.034 | -0.092 | 0.040 | 0.444 | 0.972 | 2284 |
| L_fusiform_surfavg | -0.027 | 0.031 | -0.088 | 0.033 | 0.378 | 0.972 | 2414 |
| L_inferiorparietal_surfavg | -0.033 | 0.025 | -0.083 | 0.016 | 0.186 | 0.972 | 2360 |
| L_inferiortemporal_surfavg | 0.011 | 0.026 | -0.039 | 0.061 | 0.672 | 0.972 | 2385 |
| L_isthmuscingulate_surfavg | 0.041 | 0.020 | 0.002 | 0.080 | 0.042 | 0.972 | 2460 |
| L_lateraloccipital_surfavg | -0.031 | 0.020 | -0.071 | 0.009 | 0.131 | 0.972 | 2418 |
| L_lateralorbitofrontal_surfavg | 0.002 | 0.030 | -0.058 | 0.061 | 0.961 | 0.975 | 2434 |
| L_lingual_surfavg | 0.027 | 0.023 | -0.019 | 0.073 | 0.246 | 0.972 | 2401 |
| L_medialorbitofrontal_surfavg | 0.005 | 0.034 | -0.061 | 0.072 | 0.871 | 0.973 | 2418 |
| L_middletemporal_surfavg | 0.009 | 0.021 | -0.032 | 0.050 | 0.675 | 0.972 | 2287 |
| L_parahippocampal_surfavg | -0.020 | 0.020 | -0.059 | 0.020 | 0.324 | 0.972 | 2444 |
| L_paracentral_surfavg | 0.014 | 0.020 | -0.026 | 0.053 | 0.497 | 0.972 | 2457 |
| L_parsopercularis_surfavg | 0.019 | 0.029 | -0.038 | 0.076 | 0.515 | 0.972 | 2434 |
| L_parsorbitalis_surfavg | 0.033 | 0.029 | -0.024 | 0.089 | 0.259 | 0.972 | 2438 |
| L_parstriangularis_surfavg | 0.013 | 0.023 | -0.033 | 0.059 | 0.565 | 0.972 | 2423 |
| L_pericalcarine_surfavg | -0.008 | 0.020 | -0.048 | 0.032 | 0.701 | 0.972 | 2376 |
| L_postcentral_surfavg | 0.023 | 0.023 | -0.021 | 0.067 | 0.306 | 0.972 | 2399 |
| L_posteriorcingulate_surfavg | 0.049 | 0.022 | 0.006 | 0.091 | 0.025 | 0.972 | 2457 |
| L_precentral_surfavg | 0.021 | 0.034 | -0.046 | 0.089 | 0.535 | 0.972 | 2419 |
| L_precuneus_surfavg | -0.012 | 0.021 | -0.053 | 0.029 | 0.565 | 0.972 | 2445 |
| L_rostralanteriorcingulate_surfavg | -0.023 | 0.024 | -0.070 | 0.024 | 0.341 | 0.972 | 2432 |
| L_rostralmiddlefrontal_surfavg | 0.015 | 0.020 | -0.025 | 0.054 | 0.459 | 0.972 | 2439 |
| L_superiorfrontal_surfavg | 0.046 | 0.049 | -0.051 | 0.143 | 0.352 | 0.972 | 2439 |
| L_superiorparietal_surfavg | -0.005 | 0.030 | -0.063 | 0.054 | 0.879 | 0.973 | 2398 |
| L_superiortemporal_surfavg | -0.005 | 0.036 | -0.075 | 0.064 | 0.882 | 0.973 | 2274 |
| L_supramarginal_surfavg | 0.042 | 0.032 | -0.020 | 0.104 | 0.189 | 0.972 | 2307 |
| L_frontalpole_surfavg | -0.028 | 0.030 | -0.086 | 0.030 | 0.348 | 0.972 | 2465 |
| L_temporalpole_surfavg | -0.010 | 0.020 | -0.050 | 0.029 | 0.609 | 0.972 | 2413 |
| L_transversetemporal_surfavg | -0.012 | 0.020 | -0.052 | 0.027 | 0.535 | 0.972 | 2465 |
| L_insula_surfavg | 0.003 | 0.020 | -0.037 | 0.043 | 0.892 | 0.973 | 2392 |
| R_bankssts_surfavg | 0.004 | 0.024 | -0.042 | 0.051 | 0.853 | 0.973 | 2397 |
| R_caudalanteriorcingulate_surfavg | -0.026 | 0.032 | -0.088 | 0.036 | 0.406 | 0.972 | 2456 |
| R_caudalmiddlefrontal_surfavg | -0.008 | 0.020 | -0.048 | 0.031 | 0.681 | 0.972 | 2441 |
| R_cuneus_surfavg | 0.002 | 0.031 | -0.059 | 0.062 | 0.960 | 0.975 | 2347 |
| R_entorhinal_surfavg | -0.032 | 0.034 | -0.099 | 0.035 | 0.354 | 0.972 | 2203 |
| R_fusiform_surfavg | 0.013 | 0.040 | -0.066 | 0.091 | 0.752 | 0.973 | 2421 |
| R_inferiorparietal_surfavg | -0.014 | 0.028 | -0.068 | 0.041 | 0.622 | 0.972 | 2359 |
| R_inferiortemporal_surfavg | -0.021 | 0.034 | -0.086 | 0.045 | 0.540 | 0.972 | 2395 |
| R_isthmuscingulate_surfavg | 0.027 | 0.023 | -0.019 | 0.073 | 0.246 | 0.972 | 2448 |
| R_lateraloccipital_surfavg | -0.019 | 0.020 | -0.059 | 0.021 | 0.354 | 0.972 | 2412 |
| R_lateralorbitofrontal_surfavg | 0.018 | 0.020 | -0.022 | 0.058 | 0.377 | 0.972 | 2410 |
| R_lingual_surfavg | 0.007 | 0.032 | -0.055 | 0.070 | 0.816 | 0.973 | 2405 |
| R_medialorbitofrontal_surfavg | -0.031 | 0.037 | -0.103 | 0.041 | 0.402 | 0.972 | 2397 |
| R_middletemporal_surfavg | 0.012 | 0.027 | -0.040 | 0.064 | 0.656 | 0.972 | 2369 |
| R_parahippocampal_surfavg | -0.007 | 0.021 | -0.049 | 0.035 | 0.741 | 0.973 | 2447 |
| R_paracentral_surfavg | 0.003 | 0.026 | -0.047 | 0.053 | 0.916 | 0.973 | 2454 |
| R_parsopercularis_surfavg | 0.031 | 0.028 | -0.024 | 0.085 | 0.267 | 0.972 | 2411 |
| R_parsorbitalis_surfavg | -0.021 | 0.031 | -0.081 | 0.040 | 0.504 | 0.972 | 2436 |
| R_parstriangularis_surfavg | 0.030 | 0.024 | -0.017 | 0.078 | 0.206 | 0.972 | 2402 |
| R_pericalcarine_surfavg | -0.004 | 0.030 | -0.063 | 0.056 | 0.907 | 0.973 | 2360 |
| R_postcentral_surfavg | 0.001 | 0.020 | -0.038 | 0.041 | 0.943 | 0.975 | 2415 |
| R_posteriorcingulate_surfavg | -0.012 | 0.020 | -0.051 | 0.027 | 0.553 | 0.972 | 2458 |
| R_precentral_surfavg | 0.004 | 0.027 | -0.048 | 0.056 | 0.868 | 0.973 | 2418 |
| R_precuneus_surfavg | 0.023 | 0.021 | -0.019 | 0.065 | 0.286 | 0.972 | 2444 |
| R_rostralanteriorcingulate_surfavg | -0.008 | 0.028 | -0.063 | 0.048 | 0.783 | 0.973 | 2407 |
| R_rostralmiddlefrontal_surfavg | 0.009 | 0.021 | -0.032 | 0.050 | 0.664 | 0.972 | 2417 |
| R_superiorfrontal_surfavg | 0.011 | 0.020 | -0.029 | 0.051 | 0.584 | 0.972 | 2438 |
| R_superiorparietal_surfavg | -0.011 | 0.032 | -0.073 | 0.051 | 0.731 | 0.973 | 2414 |
| R_superiortemporal_surfavg | 0.024 | 0.021 | -0.016 | 0.065 | 0.233 | 0.972 | 2346 |
| R_supramarginal_surfavg | 0.000 | 0.025 | -0.048 | 0.048 | 0.999 | 0.999 | 2333 |
| R_frontalpole_surfavg | 0.015 | 0.029 | -0.042 | 0.072 | 0.609 | 0.972 | 2455 |
| R_temporalpole_surfavg | -0.034 | 0.021 | -0.074 | 0.007 | 0.103 | 0.972 | 2351 |
| R_transversetemporal_surfavg | -0.019 | 0.020 | -0.058 | 0.021 | 0.353 | 0.972 | 2468 |
| R_insula_surfavg | 0.003 | 0.021 | -0.037 | 0.044 | 0.875 | 0.973 | 2359 |

### Table S15. Association between SV and positive schizotypy corrected for age, and sex

| DKT Regions | Partial R | Std. Err. | 95% CI, lower bound | 95% CI, upper bound | p-value | FDR p-value | Sample size per region |
| --- | --- | --- | --- | --- | --- | --- | --- |
| Laccumb | -0.028 | 0.031 | -0.089 | 0.032 | 0.355 | 0.724 | 2626 |
| Lamyg | -0.003 | 0.025 | -0.051 | 0.045 | 0.896 | 0.896 | 2635 |
| Lcaud | 0.032 | 0.019 | -0.006 | 0.070 | 0.103 | 0.724 | 2640 |
| Lhippo | 0.009 | 0.026 | -0.041 | 0.060 | 0.719 | 0.896 | 2633 |
| Lpal | -0.040 | 0.031 | -0.101 | 0.020 | 0.190 | 0.724 | 2427 |
| Lput | -0.016 | 0.020 | -0.054 | 0.022 | 0.407 | 0.724 | 2591 |
| Lthal | -0.011 | 0.030 | -0.070 | 0.048 | 0.713 | 0.896 | 2624 |
| Raccumb | -0.005 | 0.033 | -0.070 | 0.060 | 0.878 | 0.896 | 2624 |
| Ramyg | -0.013 | 0.022 | -0.055 | 0.030 | 0.562 | 0.896 | 2630 |
| Rcaud | 0.019 | 0.019 | -0.019 | 0.057 | 0.331 | 0.724 | 2641 |
| Rhippo | 0.008 | 0.025 | -0.041 | 0.057 | 0.749 | 0.896 | 2642 |
| Rpal | -0.058 | 0.034 | -0.124 | 0.008 | 0.087 | 0.724 | 2627 |
| Rput | -0.016 | 0.019 | -0.055 | 0.022 | 0.396 | 0.724 | 2625 |
| Rthal | 0.009 | 0.032 | -0.054 | 0.072 | 0.785 | 0.896 | 2639 |

### Table S16. Association between SV and negative schizotypy corrected for age, and sex

| DKT Regions | Partial R | Std. Err. | 95% CI, lower bound | 95% CI, upper bound | p-value | FDR p-value | Sample size per region |
| --- | --- | --- | --- | --- | --- | --- | --- |
| Laccumb | -0.031 | 0.025 | -0.080 | 0.017 | 0.202 | 0.429 | 2658 |
| Lamyg | 0.026 | 0.022 | -0.017 | 0.068 | 0.231 | 0.437 | 2667 |
| Lcaud | 0.030 | 0.022 | -0.012 | 0.073 | 0.164 | 0.429 | 2675 |
| Lhippo | 0.034 | 0.021 | -0.008 | 0.076 | 0.113 | 0.429 | 2669 |
| Lpal | -0.041 | 0.023 | -0.086 | 0.003 | 0.070 | 0.429 | 2458 |
| Lput | -0.017 | 0.031 | -0.078 | 0.043 | 0.573 | 0.749 | 2629 |
| Lthal | -0.009 | 0.032 | -0.072 | 0.054 | 0.782 | 0.831 | 2659 |
| Raccumb | 0.001 | 0.032 | -0.061 | 0.063 | 0.978 | 0.978 | 2657 |
| Ramyg | 0.008 | 0.025 | -0.041 | 0.057 | 0.759 | 0.831 | 2662 |
| Rcaud | 0.013 | 0.022 | -0.030 | 0.056 | 0.542 | 0.749 | 2677 |
| Rhippo | 0.029 | 0.022 | -0.014 | 0.071 | 0.189 | 0.429 | 2676 |
| Rpal | -0.034 | 0.019 | -0.072 | 0.004 | 0.081 | 0.429 | 2663 |
| Rput | -0.010 | 0.027 | -0.064 | 0.044 | 0.714 | 0.831 | 2662 |
| Rthal | -0.028 | 0.039 | -0.106 | 0.049 | 0.472 | 0.729 | 2678 |

### Fig S1. Associations between CT profiles of positive and negative schizotypy with those from CHR stages and schizophrenia

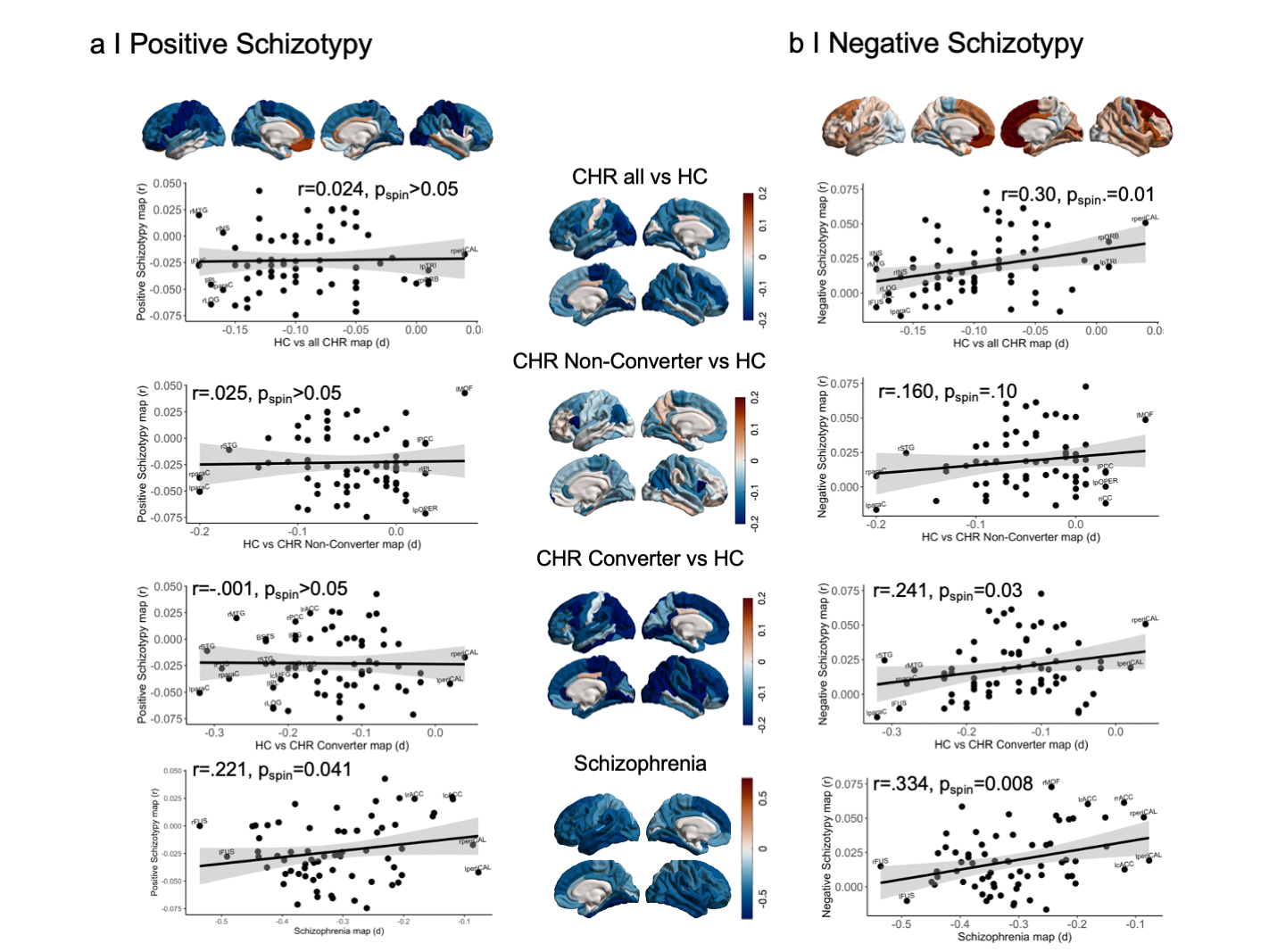

### Fig S2. Normative functional degree centrality derived from the HCP sample (n=207)

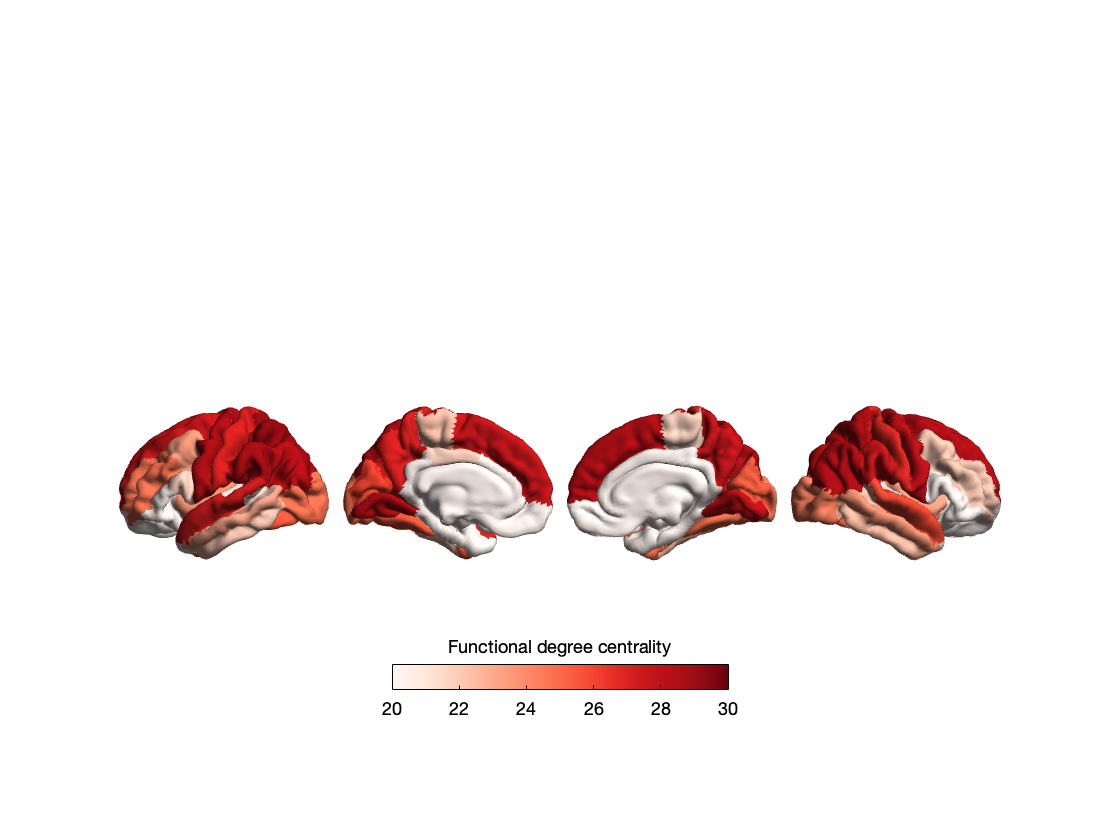

### Fig S3. Normative structural degree centrality derived from the HCP sample (n=207)

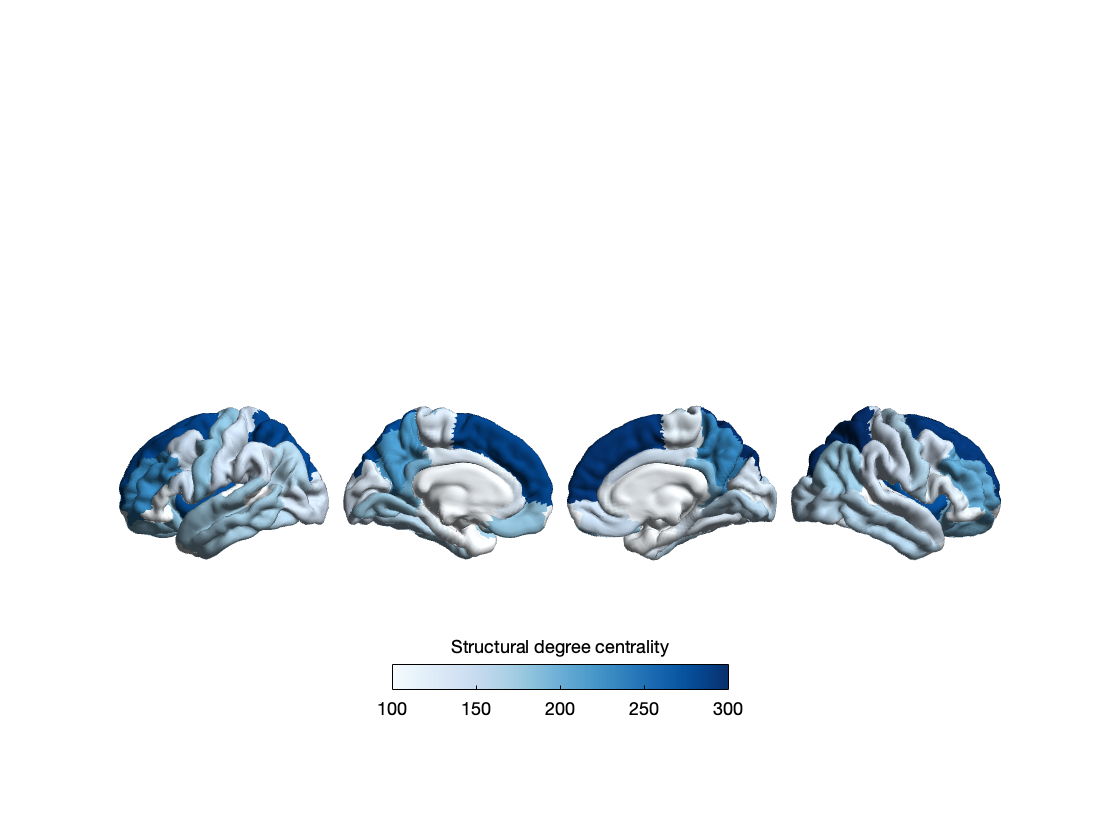

### Table S17. Functional epicenters of associations between CT and positive schizotypy

| DKT regions | Functional epicenter r-values | pspin |
| --- | --- | --- |
| L_inferiortemporal_thickavg | -0.541680148 | 0.001 |
| R_fusiform_thickavg | -0.512515271 | 0.0015 |
| L_precentral_thickavg | -0.502460361 | 0.0015 |
| R_paracentral_thickavg | -0.48829117 | 0.002 |
| L_superiorparietal_thickavg | -0.468902802 | 0.0015 |
| L_supramarginal_thickavg | -0.466722129 | 0.003 |
| L_paracentral_thickavg | -0.465377588 | 0.0025 |
| R_supramarginal_thickavg | -0.465268837 | 0.0025 |
| L_transversetemporal_thickavg | -0.463268157 | 0.001 |
| R_precentral_thickavg | -0.458660906 | 0.003 |
| R_insula_thickavg | -0.453906665 | 0.0025 |
| L_lateraloccipital_thickavg | -0.450966614 | 0 |
| L_parstriangularis_thickavg | -0.449613694 | 0.001 |
| L_fusiform_thickavg | -0.447597364 | 0.0025 |
| R_superiorparietal_thickavg | -0.444430332 | 0.0035 |
| R_postcentral_thickavg | -0.442507552 | 0.0025 |
| R_bankssts_thickavg | -0.439967713 | 0.0025 |
| L_insula_thickavg | -0.43082653 | 0.0055 |
| R_parsopercularis_thickavg | -0.43029572 | 0.0025 |
| L_postcentral_thickavg | -0.424835738 | 0.005 |
| R_pericalcarine_thickavg | -0.416999046 | 0.0065 |
| R_cuneus_thickavg | -0.413254388 | 0.0095 |
| L_caudalanteriorcingulate_thickavg | -0.410612027 | 0.0095 |
| L_bankssts_thickavg | -0.401772067 | 0.0075 |
| R_lingual_thickavg | -0.400782585 | 0.0185 |
| L_superiortemporal_thickavg | -0.39896963 | 0.0085 |
| R_inferiortemporal_thickavg | -0.391763266 | 0.0155 |
| L_parsopercularis_thickavg | -0.388610204 | 0.01 |
| L_rostralmiddlefrontal_thickavg | -0.382626214 | 0.0065 |
| R_superiortemporal_thickavg | -0.380983749 | 0.016 |
| L_cuneus_thickavg | -0.379974987 | 0.01 |
| L_lingual_thickavg | -0.379200675 | 0.0185 |
| R_transversetemporal_thickavg | -0.372581011 | 0.025 |
| L_pericalcarine_thickavg | -0.352034631 | 0.0215 |
| R_caudalanteriorcingulate_thickavg | -0.337108828 | 0.0265 |
| L_posteriorcingulate_thickavg | -0.329816534 | 0.0255 |
| R_posteriorcingulate_thickavg | -0.32437978 | 0.0285 |
| L_superiorfrontal_thickavg | -0.322737804 | 0.0305 |
| L_caudalmiddlefrontal_thickavg | -0.315727326 | 0.0155 |
| L_entorhinal_thickavg | -0.31329959 | 0.036 |
| R_lateraloccipital_thickavg | -0.306338903 | 0.0495 |
| R_parstriangularis_thickavg | -0.287850241 | 0.041 |
| L_lateralorbitofrontal_thickavg | -0.277072976 | 0.074 |
| R_lateralorbitofrontal_thickavg | -0.272083636 | 0.029 |
| R_rostralmiddlefrontal_thickavg | -0.256911327 | 0.0405 |
| R_entorhinal_thickavg | -0.255044799 | 0.05 |
| R_superiorfrontal_thickavg | -0.239138313 | 0.0735 |
| R_caudalmiddlefrontal_thickavg | -0.224098748 | 0.046 |
| L_middletemporal_thickavg | -0.222989625 | 0.056 |
| L_parahippocampal_thickavg | -0.216029004 | 0.121 |
| R_inferiorparietal_thickavg | -0.199302834 | 0.084 |
| R_middletemporal_thickavg | -0.190080766 | 0.07 |
| L_inferiorparietal_thickavg | -0.186544165 | 0.103 |
| R_parahippocampal_thickavg | -0.177121865 | 0.219 |
| L_parsorbitalis_thickavg | -0.160661335 | 0.212 |
| R_precuneus_thickavg | -0.157037594 | 0.2175 |
| L_precuneus_thickavg | -0.140488629 | 0.228 |
| L_temporalpole_thickavg | -0.105385341 | 0.305 |
| R_parsorbitalis_thickavg | -0.091513058 | 0.2655 |
| R_isthmuscingulate_thickavg | -0.046090321 | 0.4165 |
| L_isthmuscingulate_thickavg | 0.011160005 | 0.4335 |
| L_medialorbitofrontal_thickavg | 0.021895841 | 0.3865 |
| R_frontalpole_thickavg | 0.086776598 | 0.244 |
| L_frontalpole_thickavg | 0.096296975 | 0.1715 |
| R_temporalpole_thickavg | 0.129917582 | 0.209 |
| L_rostralanteriorcingulate_thickavg | 0.175427439 | 0.108 |
| R_rostralanteriorcingulate_thickavg | 0.23721669 | 0.0615 |
| R_medialorbitofrontal_thickavg | 0.258349688 | 0.032 |

### Table S18. Structural epicenters of associations between CT and positive schizotypy

| DKT regions | Structural epicenter r-values | pspin |
| --- | --- | --- |
| L_precentral_thickavg | -0.450527433 | 0.01 |
| L_superiorparietal_thickavg | -0.387073376 | 0.019 |
| L_postcentral_thickavg | -0.346522677 | 0.0525 |
| L_supramarginal_thickavg | -0.332381842 | 0.0385 |
| R_postcentral_thickavg | -0.296875232 | 0.0515 |
| R_paracentral_thickavg | -0.28088864 | 0.087 |
| R_caudalmiddlefrontal_thickavg | -0.280172167 | 0.032 |
| R_superiorparietal_thickavg | -0.270674693 | 0.0315 |
| L_caudalmiddlefrontal_thickavg | -0.26417868 | 0.107 |
| L_paracentral_thickavg | -0.25620386 | 0.1445 |
| R_precentral_thickavg | -0.245620138 | 0.0835 |
| L_inferiorparietal_thickavg | -0.242205114 | 0.08 |
| L_cuneus_thickavg | -0.234810227 | 0.1475 |
| L_middletemporal_thickavg | -0.232945211 | 0.0865 |
| L_parsopercularis_thickavg | -0.228817022 | 0.104 |
| R_parsopercularis_thickavg | -0.219468843 | 0.026 |
| R_pericalcarine_thickavg | -0.213011237 | 0.126 |
| L_superiortemporal_thickavg | -0.200442324 | 0.1595 |
| L_parstriangularis_thickavg | -0.183534435 | 0.17 |
| R_cuneus_thickavg | -0.182259429 | 0.1835 |
| L_fusiform_thickavg | -0.174608545 | 0.2235 |
| R_supramarginal_thickavg | -0.161567074 | 0.085 |
| L_transversetemporal_thickavg | -0.156825374 | 0.197 |
| L_pericalcarine_thickavg | -0.153830126 | 0.268 |
| L_rostralmiddlefrontal_thickavg | -0.142202204 | 0.2555 |
| R_inferiorparietal_thickavg | -0.140757458 | 0.0385 |
| L_inferiortemporal_thickavg | -0.139842846 | 0.233 |
| L_parahippocampal_thickavg | -0.136357659 | 0.308 |
| L_precuneus_thickavg | -0.134178638 | 0.308 |
| L_lateraloccipital_thickavg | -0.129769313 | 0.3235 |
| L_insula_thickavg | -0.113380386 | 0.3345 |
| L_isthmuscingulate_thickavg | -0.079776739 | 0.396 |
| L_parsorbitalis_thickavg | -0.058150885 | 0.455 |
| R_middletemporal_thickavg | -0.056731328 | 0.117 |
| L_bankssts_thickavg | -0.054444487 | 0.448 |
| R_posteriorcingulate_thickavg | -0.053780861 | 0.413 |
| R_fusiform_thickavg | -0.051653246 | 0.2025 |
| R_parstriangularis_thickavg | -0.043190004 | 0.255 |
| L_lingual_thickavg | -0.038636232 | 0.4695 |
| R_lingual_thickavg | -0.026244175 | 0.3805 |
| R_lateraloccipital_thickavg | -0.023249738 | 0.3095 |
| R_precuneus_thickavg | -0.016512143 | 0.3905 |
| R_superiortemporal_thickavg | -0.013499584 | 0.323 |
| R_isthmuscingulate_thickavg | -0.007934888 | 0.4175 |
| R_insula_thickavg | 0.021073885 | 0.8015 |
| R_rostralmiddlefrontal_thickavg | 0.030814987 | 0.5095 |
| R_transversetemporal_thickavg | 0.032547412 | 0.4865 |
| R_frontalpole_thickavg | 0.058154123 | 0.434 |
| L_superiorfrontal_thickavg | 0.064981736 | 0.383 |
| R_inferiortemporal_thickavg | 0.065164281 | 0.4465 |
| L_temporalpole_thickavg | 0.099028243 | 0.205 |
| R_bankssts_thickavg | 0.101564115 | 0.3085 |
| R_parsorbitalis_thickavg | 0.102204491 | 0.336 |
| R_entorhinal_thickavg | 0.102875849 | 0.354 |
| L_entorhinal_thickavg | 0.116996487 | 0.1665 |
| R_parahippocampal_thickavg | 0.121845215 | 0.3505 |
| R_superiorfrontal_thickavg | 0.126388892 | 0.321 |
| L_posteriorcingulate_thickavg | 0.153332731 | 0.2245 |
| L_frontalpole_thickavg | 0.185829054 | 0.1745 |
| L_caudalanteriorcingulate_thickavg | 0.199543337 | 0.096 |
| L_lateralorbitofrontal_thickavg | 0.210287497 | 0.078 |
| R_temporalpole_thickavg | 0.260177451 | 0.066 |
| R_rostralanteriorcingulate_thickavg | 0.274163057 | 0.072 |
| L_rostralanteriorcingulate_thickavg | 0.284256954 | 0.0455 |
| R_caudalanteriorcingulate_thickavg | 0.290121173 | 0.0655 |
| L_medialorbitofrontal_thickavg | 0.315712575 | 0.0295 |
| R_lateralorbitofrontal_thickavg | 0.324852123 | 0.0425 |
| R_medialorbitofrontal_thickavg | 0.352655488 | 0.025 |

### Table S19. Functional epicenters of associations between CT and negative schizotypy

| DKT regions | Functional epicenter r-values | pspin |
| --- | --- | --- |
| R_fusiform_thickavg | -0.486010279 | 0.003 |
| R_superiortemporal_thickavg | -0.441189947 | 0.0095 |
| L_bankssts_thickavg | -0.439478655 | 0.0095 |
| L_superiortemporal_thickavg | -0.428526131 | 0.018 |
| L_inferiortemporal_thickavg | -0.4282116 | 0.0095 |
| R_superiorparietal_thickavg | -0.424477627 | 0.0085 |
| L_fusiform_thickavg | -0.422526419 | 0.015 |
| L_entorhinal_thickavg | -0.416708471 | 0.0225 |
| L_postcentral_thickavg | -0.415400515 | 0.004 |
| R_paracentral_thickavg | -0.414600402 | 0.015 |
| L_parahippocampal_thickavg | -0.404285021 | 0.008 |
| R_precentral_thickavg | -0.400258903 | 0.0105 |
| L_transversetemporal_thickavg | -0.398740616 | 0.0175 |
| L_superiorparietal_thickavg | -0.39756996 | 0.0085 |
| L_insula_thickavg | -0.387598935 | 0.025 |
| R_inferiortemporal_thickavg | -0.373901367 | 0.0245 |
| R_lateraloccipital_thickavg | -0.371486939 | 0.0105 |
| R_pericalcarine_thickavg | -0.369244987 | 0.0125 |
| L_precentral_thickavg | -0.367821631 | 0.0305 |
| R_lingual_thickavg | -0.361181989 | 0.049 |
| L_paracentral_thickavg | -0.356513044 | 0.041 |
| R_parahippocampal_thickavg | -0.351400486 | 0.036 |
| R_supramarginal_thickavg | -0.348486851 | 0.0265 |
| L_supramarginal_thickavg | -0.347902378 | 0.039 |
| L_lateraloccipital_thickavg | -0.342756874 | 0.0145 |
| R_transversetemporal_thickavg | -0.335401738 | 0.0565 |
| R_posteriorcingulate_thickavg | -0.333242032 | 0.027 |
| L_cuneus_thickavg | -0.328429022 | 0.036 |
| R_postcentral_thickavg | -0.32662095 | 0.0545 |
| L_posteriorcingulate_thickavg | -0.326448996 | 0.0545 |
| R_entorhinal_thickavg | -0.324518811 | 0.013 |
| R_cuneus_thickavg | -0.316737326 | 0.057 |
| L_superiorfrontal_thickavg | -0.316328742 | 0.0785 |
| R_bankssts_thickavg | -0.31575878 | 0.0725 |
| L_caudalmiddlefrontal_thickavg | -0.303260063 | 0.0525 |
| R_insula_thickavg | -0.295932794 | 0.0765 |
| R_superiorfrontal_thickavg | -0.292868364 | 0.048 |
| R_caudalanteriorcingulate_thickavg | -0.288039176 | 0.0545 |
| L_parstriangularis_thickavg | -0.275081808 | 0.1445 |
| L_precuneus_thickavg | -0.269198416 | 0.0705 |
| L_lingual_thickavg | -0.258816837 | 0.104 |
| L_pericalcarine_thickavg | -0.258170425 | 0.079 |
| L_caudalanteriorcingulate_thickavg | -0.252185783 | 0.105 |
| L_temporalpole_thickavg | -0.250454461 | 0.1015 |
| R_caudalmiddlefrontal_thickavg | -0.247540655 | 0.032 |
| L_rostralmiddlefrontal_thickavg | -0.246358468 | 0.1595 |
| L_parsopercularis_thickavg | -0.244155873 | 0.19 |
| L_inferiorparietal_thickavg | -0.243111634 | 0.0795 |
| R_precuneus_thickavg | -0.242577544 | 0.12 |
| R_inferiorparietal_thickavg | -0.219461536 | 0.048 |
| R_parsopercularis_thickavg | -0.216106526 | 0.1065 |
| L_lateralorbitofrontal_thickavg | -0.209305933 | 0.2295 |
| R_parstriangularis_thickavg | -0.207308696 | 0.123 |
| L_middletemporal_thickavg | -0.180481979 | 0.181 |
| R_middletemporal_thickavg | -0.134239674 | 0.1565 |
| L_parsorbitalis_thickavg | -0.129350392 | 0.4135 |
| R_isthmuscingulate_thickavg | -0.122828013 | 0.3025 |
| L_isthmuscingulate_thickavg | -0.113145869 | 0.341 |
| R_temporalpole_thickavg | -0.093699235 | 0.2425 |
| R_rostralmiddlefrontal_thickavg | -0.06805229 | 0.281 |
| L_medialorbitofrontal_thickavg | -0.059581721 | 0.457 |
| R_lateralorbitofrontal_thickavg | -0.052411493 | 0.328 |
| L_frontalpole_thickavg | -0.040604691 | 0.5905 |
| R_medialorbitofrontal_thickavg | -0.018746628 | 0.4835 |
| R_parsorbitalis_thickavg | -0.005409198 | 0.4565 |
| L_rostralanteriorcingulate_thickavg | 0.015941545 | 0.4 |
| R_frontalpole_thickavg | 0.044061744 | 0.307 |
| R_rostralanteriorcingulate_thickavg | 0.121252941 | 0.239 |

### Table S20. Structural epicenters of associations between CT and negative schizotypy

| DKT regions | Structural epicenter r-values | pspin |
| --- | --- | --- |
| L_parahippocampal_thickavg | -0.456381671 | 0.0185 |
| R_temporalpole_thickavg | -0.362083303 | 0.0015 |
| R_caudalmiddlefrontal_thickavg | -0.353785336 | 0.0255 |
| R_paracentral_thickavg | -0.342905438 | 0.0435 |
| L_parsopercularis_thickavg | -0.3276488 | 0.067 |
| L_precuneus_thickavg | -0.30776082 | 0.1145 |
| R_cuneus_thickavg | -0.299329052 | 0.0945 |
| R_superiorparietal_thickavg | -0.2860747 | 0.0775 |
| L_middletemporal_thickavg | -0.272028989 | 0.178 |
| R_entorhinal_thickavg | -0.266706404 | 0.02 |
| L_fusiform_thickavg | -0.263043115 | 0.1235 |
| L_supramarginal_thickavg | -0.260733469 | 0.1235 |
| L_inferiorparietal_thickavg | -0.25432104 | 0.191 |
| L_lateraloccipital_thickavg | -0.235610147 | 0.2695 |
| L_precentral_thickavg | -0.235353145 | 0.107 |
| R_parstriangularis_thickavg | -0.23356289 | 0.0735 |
| L_pericalcarine_thickavg | -0.226560308 | 0.2475 |
| L_paracentral_thickavg | -0.202791913 | 0.286 |
| L_caudalmiddlefrontal_thickavg | -0.198459146 | 0.293 |
| R_postcentral_thickavg | -0.197517862 | 0.284 |
| R_parsopercularis_thickavg | -0.159553304 | 0.2885 |
| R_rostralmiddlefrontal_thickavg | -0.157918027 | 0.085 |
| L_rostralanteriorcingulate_thickavg | -0.142391894 | 0.08 |
| R_lingual_thickavg | -0.138259598 | 0.023 |
| L_transversetemporal_thickavg | -0.13786686 | 0.445 |
| L_isthmuscingulate_thickavg | -0.136628108 | 0.448 |
| R_middletemporal_thickavg | -0.132741897 | 0.8585 |
| R_inferiortemporal_thickavg | -0.132491884 | 0.2995 |
| L_entorhinal_thickavg | -0.121645732 | 0.24 |
| L_postcentral_thickavg | -0.092612607 | 0.5015 |
| L_parstriangularis_thickavg | -0.084343369 | 0.567 |
| R_pericalcarine_thickavg | -0.083138389 | 0.637 |
| L_parsorbitalis_thickavg | -0.081260884 | 0.5515 |
| R_rostralanteriorcingulate_thickavg | -0.07625479 | 0.0855 |
| R_isthmuscingulate_thickavg | -0.055094515 | 0.143 |
| R_parahippocampal_thickavg | -0.051823428 | 0.4055 |
| L_superiortemporal_thickavg | -0.030600467 | 0.772 |
| R_precuneus_thickavg | -0.018873909 | 0.205 |
| L_posteriorcingulate_thickavg | -0.01315417 | 0.318 |
| R_lateraloccipital_thickavg | -0.010780333 | 0.1185 |
| L_bankssts_thickavg | -0.005790831 | 0.3185 |
| L_medialorbitofrontal_thickavg | -0.001652915 | 0.2665 |
| L_lingual_thickavg | -0.00095885 | 0.1965 |
| R_lateralorbitofrontal_thickavg | 0.003630264 | 0.6545 |
| R_superiorfrontal_thickavg | 0.013252736 | 0.5465 |
| R_frontalpole_thickavg | 0.016668482 | 0.8665 |
| R_supramarginal_thickavg | 0.033910472 | 0.3815 |
| R_fusiform_thickavg | 0.03477334 | 0.499 |
| L_superiorfrontal_thickavg | 0.040156936 | 0.71 |
| R_superiortemporal_thickavg | 0.053382429 | 0.531 |
| R_parsorbitalis_thickavg | 0.087852296 | 0.58 |
| R_bankssts_thickavg | 0.143779998 | 0.4665 |
| R_inferiorparietal_thickavg | 0.169802347 | 0.0705 |
| L_superiorparietal_thickavg | 0.204117461 | 0.0685 |
| L_temporalpole_thickavg | 0.208877256 | 0.3145 |
| R_precentral_thickavg | 0.22632278 | 0.034 |
| L_inferiortemporal_thickavg | 0.274993147 | 0.0715 |
| L_lateralorbitofrontal_thickavg | 0.27499686 | 0.19 |
| R_posteriorcingulate_thickavg | 0.286734894 | 0.132 |
| L_caudalanteriorcingulate_thickavg | 0.306496755 | 0.0705 |
| L_rostralmiddlefrontal_thickavg | 0.321708508 | 0.0175 |
| R_caudalanteriorcingulate_thickavg | 0.340200895 | 0.046 |
| L_cuneus_thickavg | 0.346424602 | 0.017 |
| R_medialorbitofrontal_thickavg | 0.365467806 | 0.029 |
| L_frontalpole_thickavg | 0.376647635 | 0.0365 |
| L_insula_thickavg | 0.39228198 | 0.0175 |
| R_transversetemporal_thickavg | 0.416714908 | 0.027 |
| R_insula_thickavg | 0.50482831 | 0.006 |

### Funding and Acknowledgements

| FOR 2107-MR | This work was funded by the German Research Foundation (DFG grants FOR2107 KI588/14-1, and KI588/14-2, and KI588/20-1, KI588/22-1 to Tilo Kircher, Marburg, Germany). Biosamples and corresponding data were sampled, processed and stored in the Marburg Biobank CBBMR. |
| --- | --- |
| FOR2107-MS | This work was funded by the German Research Foundation (DFG, grant FOR2107 DA1151/5-1, DA1151/5-2, DA1151/9-1, DA1151/10-1, DA1151/11-1 to UD) and the Interdisciplinary Center for Clinical Research (IZKF) of the medical faculty of M√ºnster (grant Dan3/022/22 to UD). |
| Bonn1, Bonn2, Bonn3 | DFG Et 31/2-1 |
| FOR2107-MR, Jena | DFG (NE2254/1-2, NE2254/2-1, NE2254/3-1, NE2254/4-1) |
| IGP and ASRB | NHMRC APP630471; APP1051672; APP1081603 |
| FIDMAG | AGAUR, Generalitat de Catalunya. CIBERSAM ISCIII |
| Zurich 2 | Collegium Heveticum |
| ENIGMA SZ WG | This work was in part supported by the National Institute of Mental Health of the National Institutes of Health under award numbers R01MH121246 and R01MH1345261. |
| Paris | ANR MNP VIP, ANR Labex BioPsy |
| Geneva | Swiss National Science Foundation grant number 159440 |
